# Discovery of 42 Genome-Wide Significant Loci Associated with Dyslexia

**DOI:** 10.1101/2021.08.20.21262334

**Authors:** Catherine Doust, Pierre Fontanillas, Else Eising, Scott D Gordon, Zhengjun Wang, Gökberk Alagöz, Barbara Molz, 23andMe Research Team, Quantitative Trait Working Group of the GenLang Consortium, Beate St Pourcain, Clyde Francks, Riccardo E Marioni, Jingjing Zhao, Silvia Paracchini, Joel B Talcott, Anthony P Monaco, John F Stein, Jeffrey R Gruen, Richard K Olson, Erik G Willcutt, John C DeFries, Bruce F Pennington, Shelley D Smith, Margaret J Wright, Nicholas G Martin, Adam Auton, Timothy C Bates, Simon E Fisher, Michelle Luciano

## Abstract

Reading and writing are crucial for many aspects of modern life but up to 1 in 10 children are affected by dyslexia [1, 2], which can persist into adulthood. Family studies of dyslexia suggest heritability up to 70% [3, 4], yet no convincing genetic markers have been found due to limited study power [5]. Here, we present a genome-wide association study representing a 20-fold increase in sample size from prior work, with 51,800 adults self-reporting a dyslexia diagnosis and 1,087,070 controls. We identified 42 independent genome-wide significant loci: 17 are in genes linked to or pleiotropic with cognitive ability/educational attainment; 25 are novel and may be more specifically associated with dyslexia. Twenty-three loci (12 novel) were validated in independent cohorts of Chinese and European ancestry. We confirmed a similar genetic aetiology of dyslexia between sexes, and found genetic covariance with many traits, including ambidexterity, but not neuroanatomical measures of language-related circuitry. Causal analyses revealed a directional effect of dyslexia on attention deficit hyperactivity disorder and bidirectional effects on socio-educational traits but these relationships require further investigation. Dyslexia polygenic scores explained up to 6% of variance in reading traits in independent cohorts, and might in future enable earlier identification and remediation of dyslexia.

## INTRODUCTION

The ability to read is crucial for success at school and access to employment, information, and health/social services, and is related to attained socio-economic status [6]. Dyslexia is a neurodevelopmental disorder characterised by severe reading difficulties, present in between 5 and 17.5% of the population, depending on diagnostic criteria [1, 2]. It often involves impaired phonological processing (the decoding of sound units, or phonemes, within words) and frequently co-occurs with psychiatric and other developmental disorders [7], especially attention-deficit hyperactivity disorder (ADHD) [8, 9] and speech and language disorders [10, 11]. Dyslexia may represent the low extreme of a continuum of reading ability, a complex multifactorial trait with heritability estimates ranging from 40 to 80% [12, 13]. Identifying genetic risk factors not only aids increased understanding of the biological mechanisms, but may also expand diagnostic capabilities, facilitating earlier identification of individuals prone to dyslexia and comorbid disorders for specific support.

Prior genome-wide investigations of dyslexia have been limited to linkage analyses of affected families [14] or modest (N < 2,300 cases) association studies of diagnosed children and adolescents [5]. Candidate genes from linkage studies show inconsistent replication and genome-wide association studies (GWAS) have not found significant associations, although LOC388780 and VEPH1 were supported in gene-based tests [5]. Larger cohorts are vital for increasing sensitivity to detect novel genetic associations of small effect. We present the largest dyslexia GWAS to date, with 51,800 adults self-reporting a dyslexia diagnosis and 1,087,070 controls, all of whom are research participants with the personal genetics company 23andMe, Inc. We validate our association discoveries in independent cohorts, provide functional annotations of significant variants (mainly SNPs, single-nucleotide polymorphisms) and potential causal genes, and estimates of SNP-based heritability. Lastly, we investigate genetic correlations with reading and related skills, health, socio-economic, and psychiatric measures, and test for causal effects of dyslexia on ADHD, cognitive-educational abilities, and economic outcomes.

## RESULTS

### Genome-wide associations

The full dataset included 51,800 (21,513 males, 30,287 females) participants responding “yes” to the question “Have you been diagnosed with dyslexia?” (cases) and 1,087,070 (446,054 males, 641,016 females) participants responding “no” (controls). Participants were ≥18 years (mean ages of cases and controls were 49.9 years (SE 16.3) and 51.8 years (SE 16.6), respectively). We identified 42 independent genome-wide significant associated loci (*p* < 5 × 10^−8^) and 64 loci with suggestive significance (*p* < 1 × 10^−6^) (Figure 1; Supplementary Table 1). Genomic inflation was moderate (*λ*_*GC*_ = 1.18) and consistent with polygenicity (see Q-Q plot, Supplementary Figure 1). All significant variants were autosomal, except Xq27.3:rs5904158; their regional association plots are shown in Supplementary Figures 4.i.-xl. Seventeen index variants were in high linkage with published (genome-wide significant) associated SNPs in the NHGRI GWAS Catalog [15] (15 were associated with cognitive/educational traits; Supplementary Tables 1 and 2). Of these, four (rs4696277, rs34349354, rs9696811, rs72841395) were pleiotropic with general cognitive ability [16] based on HEIDI-outlier analysis [17]. The remaining 25 associated loci with no evidence of published genome-wide associations with traits expected to overlap with dyslexia or were not pleiotropic with cognitive ability were considered as novel (Table 1).

**Figure 1.**
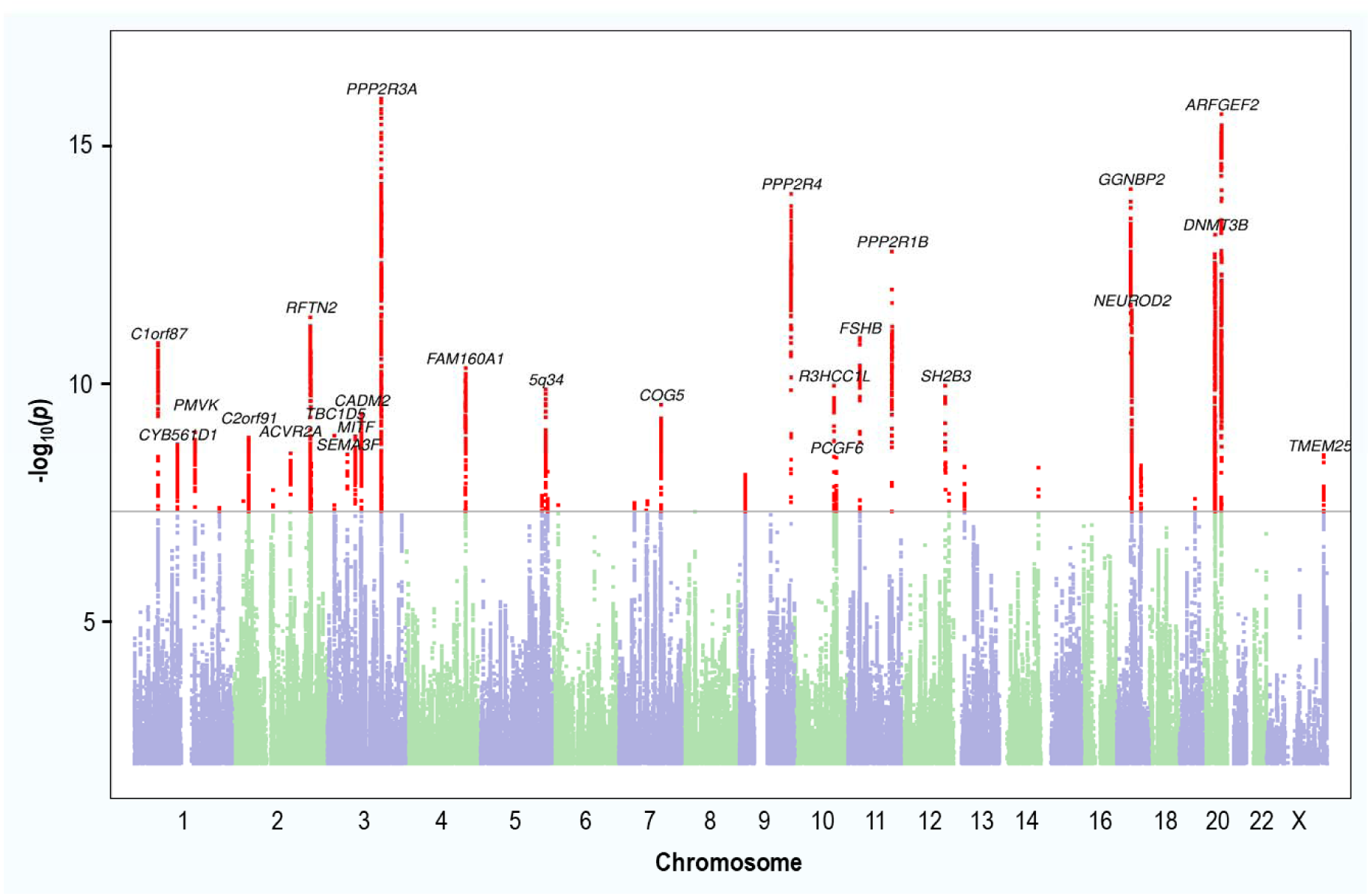
Manhattan plot of the genome-wide association analysis of dyslexia. The *y* axis represents the negative log_10_ *p* value for association of SNPs with self-reported dyslexia diagnosis from 51,800 individuals and 1,087,070 controls. The threshold for genome-wide significance (*p* < 5 × 10^−8^) is represented by a horizontal purple line. Genome-wide significant variants in the 42 genome-wide significant loci are red. Variants located within a distance of < 250 kb of each other are considered as one locus.

**Table 1.**
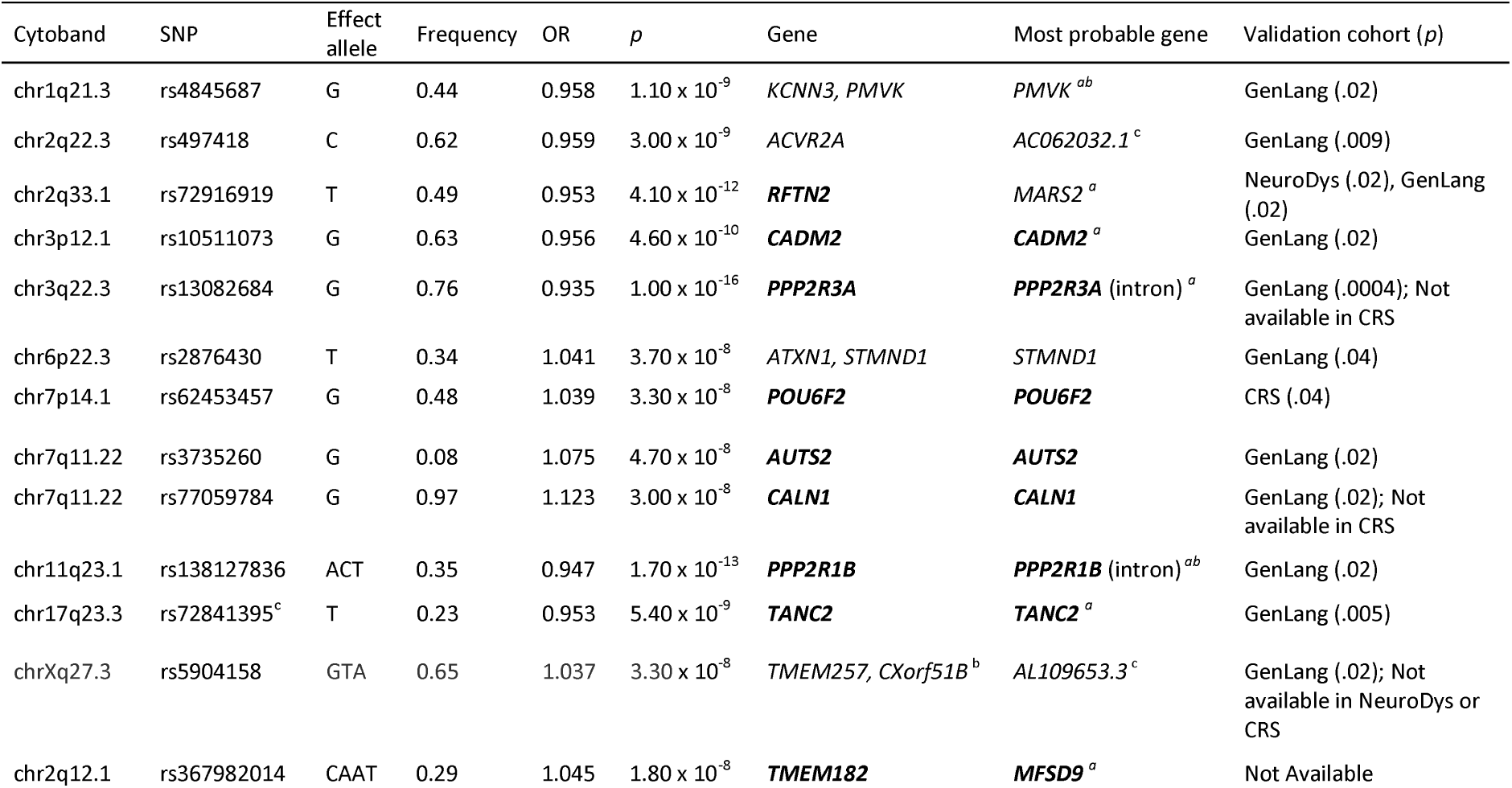
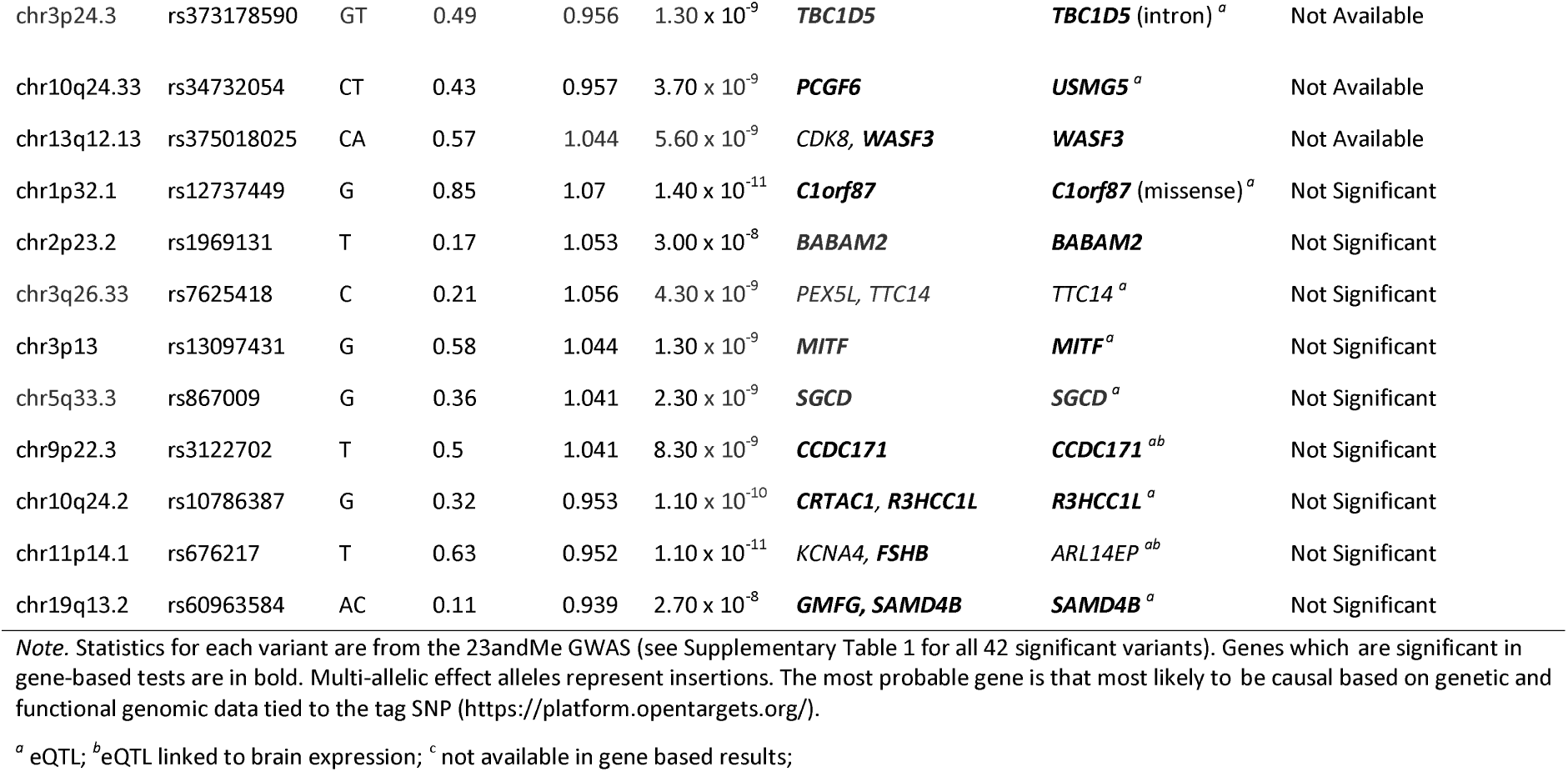
25 Novel Variants Associated with Dyslexia Including Gene-based Results, eQTL Status, Expression in Brain, and Validation in Three Independent Cohorts (GenLang Consortium, Chinese Reading Study (CRS), NeuroDys).

| Cytoband | SNP | Effect allele | Frequency | OR | *p* | Gene | Most probable gene | Validation cohort (*p*) |
| --- | --- | --- | --- | --- | --- | --- | --- | --- |
| chr1q21.3 | rs4845687 | G | 0.44 | 0.958 | $1.10 \times 10^{-9}$ | *KCNN3*, *PMVK* | *PMVK*<sup>ab</sup> | GenLang (.02) |
| chr2q22.3 | rs497418 | C | 0.62 | 0.959 | $3.00 \times 10^{-9}$ | *ACVR2A* | *AC062032.1*<sup>c</sup> | GenLang (.009) |
| chr2q33.1 | rs72916919 | T | 0.49 | 0.953 | $4.10 \times 10^{-12}$ | ***RFTN2*** | *MARS2*<sup>a</sup> | NeuroDys (.02), GenLang (.02) |
| chr3p12.1 | rs10511073 | G | 0.63 | 0.956 | $4.60 \times 10^{-10}$ | ***CADM2*** | ***CADM2***<sup>a</sup> | GenLang (.02) |
| chr3q22.3 | rs13082684 | G | 0.76 | 0.935 | $1.00 \times 10^{-16}$ | ***PPP2R3A*** | ***PPP2R3A*** (intron)<sup>a</sup> | GenLang (.0004); Not available in CRS |
| chr6p22.3 | rs2876430 | T | 0.34 | 1.041 | $3.70 \times 10^{-8}$ | *ATXN1*, *STMND1* | *STMND1* | GenLang (.04) |
| chr7p14.1 | rs62453457 | G | 0.48 | 1.039 | $3.30 \times 10^{-8}$ | ***POU6F2*** | ***POU6F2*** | CRS (.04) |
| chr7q11.22 | rs3735260 | G | 0.08 | 1.075 | $4.70 \times 10^{-8}$ | ***AUTS2*** | ***AUTS2*** | GenLang (.02) |
| chr7q11.22 | rs77059784 | G | 0.97 | 1.123 | $3.00 \times 10^{-8}$ | ***CALN1*** | ***CALN1*** | GenLang (.02); Not available in CRS |
| chr11q23.1 | rs138127836 | ACT | 0.35 | 0.947 | $1.70 \times 10^{-13}$ | ***PPP2R1B*** | ***PPP2R1B*** (intron)<sup>ab</sup> | GenLang (.02) |
| chr17q23.3 | rs72841395<sup>c</sup> | T | 0.23 | 0.953 | $5.40 \times 10^{-9}$ | ***TANC2*** | ***TANC2***<sup>a</sup> | GenLang (.005) |
| chrXq27.3 | rs5904158 | GTA | 0.65 | 1.037 | $3.30 \times 10^{-8}$ | *TMEM257*, *CXorf51B*<sup>b</sup> | *AL109653.3*<sup>c</sup> | GenLang (.02); Not available in NeuroDys or CRS |
| chr2q12.1 | rs367982014 | CAAT | 0.29 | 1.045 | $1.80 \times 10^{-8}$ | ***TMEM182*** | ***MFSD9***<sup>a</sup> | Not Available |
| chr3p24.3 | rs373178590 | GT | 0.49 | 0.956 | 1.30 x 10<sup>-9</sup> | **TBC1D5** | **TBC1D5** (intron)<sup>a</sup> | Not Available |
| chr10q24.33 | rs34732054 | CT | 0.43 | 0.957 | 3.70 x 10<sup>-9</sup> | **PCGF6** | **USMG5**<sup>a</sup> | Not Available |
| chr13q12.13 | rs375018025 | CA | 0.57 | 1.044 | 5.60 x 10<sup>-9</sup> | **CDK8, WASF3** | **WASF3** | Not Available |
| chr1p32.1 | rs12737449 | G | 0.85 | 1.07 | 1.40 x 10<sup>-11</sup> | **C1orf87** | **C1orf87** (missense)<sup>a</sup> | Not Significant |
| chr2p23.2 | rs1969131 | T | 0.17 | 1.053 | 3.00 x 10<sup>-8</sup> | **BABAM2** | **BABAM2** | Not Significant |
| chr3q26.33 | rs7625418 | C | 0.21 | 1.056 | 4.30 x 10<sup>-9</sup> | *PEX5L, TTC14* | *TTC14*<sup>a</sup> | Not Significant |
| chr3p13 | rs13097431 | G | 0.58 | 1.044 | 1.30 x 10<sup>-9</sup> | **MITF** | **MITF**<sup>a</sup> | Not Significant |
| chr5q33.3 | rs867009 | G | 0.36 | 1.041 | 2.30 x 10<sup>-9</sup> | **SGCD** | **SGCD**<sup>a</sup> | Not Significant |
| chr9p22.3 | rs3122702 | T | 0.5 | 1.041 | 8.30 x 10<sup>-9</sup> | **CCDC171** | **CCDC171**<sup>ab</sup> | Not Significant |
| chr10q24.2 | rs10786387 | G | 0.32 | 0.953 | 1.10 x 10<sup>-10</sup> | **CRTAC1, R3HCC1L** | **R3HCC1L**<sup>a</sup> | Not Significant |
| chr11p14.1 | rs676217 | T | 0.63 | 0.952 | 1.10 x 10<sup>-11</sup> | *KCNA4, FSHB* | *ARL14EP*<sup>ab</sup> | Not Significant |
| chr19q13.2 | rs60963584 | AC | 0.11 | 0.939 | 2.70 x 10<sup>-8</sup> | **GMFG, SAMD4B** | **SAMD4B**<sup>a</sup> | Not Significant |

*Note.* Statistics for each variant are from the 23andMe GWAS (see Supplementary Table 1 for all 42 significant variants). Genes which are significant in gene-based tests are in bold. Multi-allelic effect alleles represent insertions. The most probable gene is that most likely to be causal based on genetic and functional genomic data tied to the tag SNP (https://platform.opentargets.org/).
<sup>a</sup> eQTL; <sup>b</sup> eQTL linked to brain expression; <sup>c</sup> not available in gene based results;

Of 38 associated loci (the 4 remaining were tagged by indels unavailable in validation cohorts), three (rs13082684; rs34349354; rs11393101) were significant at a Bonferroni-corrected level (P<.05/38) in the GenLang consortium GWAS meta-analysis (Eising et al., in preparation) of reading (*N* = 33,959) and spelling (*N* = 18,514) ability. At *p* < 0.05, 18 were observed in GenLang, three in the NeuroDys case-control GWAS [5] (*N* = 2,274 cases), and five in the Chinese Reading Study of reading accuracy and fluency (*N* = 2,270; Supplementary Method) (Table 1; Supplementary Tables 3-6).

Gene-based tests identified 173 significantly associated genes (Supplementary Table 7) but no significantly enriched biological pathways (Supplementary Table 8). Using LDSC, we estimated the liability-scale SNP-based heritability of dyslexia to be h^2^_SNP_ = 0.152 (SE = 0.006), assuming a population prevalence of 5% (from 23andMe), and h^2^_SNP_ = 0.189 (SE = 0.008) for a 10% prevalence [1, 2].

We also performed sex-specific GWAS and age-specific GWAS (younger or older than 55 years) because dyslexia prevalence was higher in our younger (5.34% in 20 to 30-year olds) than older (3.23% in 80 to 90-year olds) participants. These showed high consistency with the main GWAS (Supplementary Figures 2 and 3). Genetic correlation estimated by LDSC was 0.91 (*p* = 8.26 × 10^−253^) in males and females, and 0.97 (*p* = 2.32 × 10^−268^) between younger and older adults (Supplementary Table 16).

### Fine-mapping and functional annotations

Within the credible variant set (Supplementary Table 1), missense variants were the most common (55%) of the coding variants; Supplementary Figure 5 summarises all predicted variant effects. Predicted deleterious variants (by SIFT score) were identified in *R3HCC1L, SH2B3, CCDC171, C1orf87, LOXL4, DLAT, ALG9*, and *SORT1*. Within the credible variant set no genes were especially intolerant to functional variation (smallest LoFtool percentile was .39). For the 42 associated loci, the most probable gene targets of each was estimated by the Overall V2G score from OpenTargets (Supplementary Table 9). Two of the index variants could be causal: the missense variant, rs12737449 (*C1orf87*), and variant rs3735260 (*AUTS2*) had Combined Annotation Dependent Depletion (CADD) scores suggestive of deleteriousness to gene function [18] (Supplementary Table 10). The *AUTS2* variant RegulomeDB rank of 2b indicated a regulatory role; its chromatin state supported location at an active transcription start site [19, 20]. The function of genes located at (or nearby) significant index variants appears in Supplementary Table 11.

Of the 173 significant genes from genome-wide gene-based tests in MAGMA, 129 could be functionally annotated (Supplementary Table 12). Protein-coding and non-coding sequences are actively conserved in approximately three quarters of these genes, 63% are more intolerant to variation than average, and 33% are intolerant to loss-of-function mutations. Gene property analysis for general tissues confirmed the importance of brain and specific regions (Supplementary Tables 13 and 14). Levels of brain expression for the 136 genes proximal to the top single variants are shown in Supplementary Table 15. Twenty-one showed high general brain expression levels (of these, *PPP1R1B, NPM1, PMVK*, and *WASF3* were significant in gene-based tests). Of the 12 brain regions assessed, gene expression was generally highest in the cerebellar hemisphere, cerebellum, and cerebral cortex (consistent with gene property analysis results). For the 173 significant genes from gene-based tests, there was enrichment of differentially expressed genes in brain subcortical and cortical areas (Supplementary Figure 6).

### Partitioned heritability

SNP-based heritability of dyslexia partitioned by functional annotation showed significant enrichment for conserved regions and H3K4me1 clusters (Supplementary Table 16 and Supplementary Figure 7). There was enrichment in genes expressed in the frontal cortex, cortex, and anterior cingulate cortex (*p* < 4.17 × 10^−3^) (Supplementary Table 17 and Supplementary Figure 8), but not for brain cell type (Supplementary Table 18 and Supplementary Figure 9). Enrichment was seen in enhancer and promoter regions with the chromatin marks H3M4me1 and H3K4me3 chromatin marks in multiple central nervous system (CNS) tissues (Supplementary Tables 19 and 20; Supplementary Figures 10 and 11). Reading, an offshoot of spoken language, is a uniquely human trait, but there was no enrichment for a range of annotations related to human evolution spanning the last 30 million to 50,000 years [21] (Supplementary Table 21).

### Genetic correlations (LDSC)

Genetic correlations were estimated for 98 traits (Supplementary Table 22 and Figure 2) including reading and spelling measures from GenLang (Supplementary Figure 12) and brain subcortical structure volumes, total cortical surface area and thickness from the Enhancing Neuro Imaging Genetic through Meta-Analysis (ENIGMA) consortium. Sixty-three traits showed genetic correlations with dyslexia at the Bonferroni-corrected significance threshold (P<0.05/98; Figure 2). Genetic correlations (*r*_*g*_) with quantitative reading and spelling measures ranged from -.70 to -.75, with phoneme awareness -0.62, and nonword repetition -.45. Childhood/adolescent performance IQ *r*_*g*_ was lower (-.19) than adult verbal-numerical reasoning [22] (-.50) and work-related/vocational qualifications (.50) but similar to childhood IQ and educational attainment [23] (-.32 and -.22, respectively). Positive *r*_*g*_ included: jobs involving heavy manual work [23] (.40), ADHD [24] (.53), equal use of right and left hands [23] (.38), and pain measures [23] (average = .31). Of the 11 ENIGMA measures tested, only intracranial volume was significantly correlated with dyslexia (*r*_*g*_ = - .14). Targeted investigation of 80 UK Biobank structural neuroimaging measures including surface-based morphometry and diffusion-weighted imaging for brain circuitry linked to language were non-significant at a corrected significance level using spectral decomposition of matrices (Supplementary Table 23).

**Figure 2.**
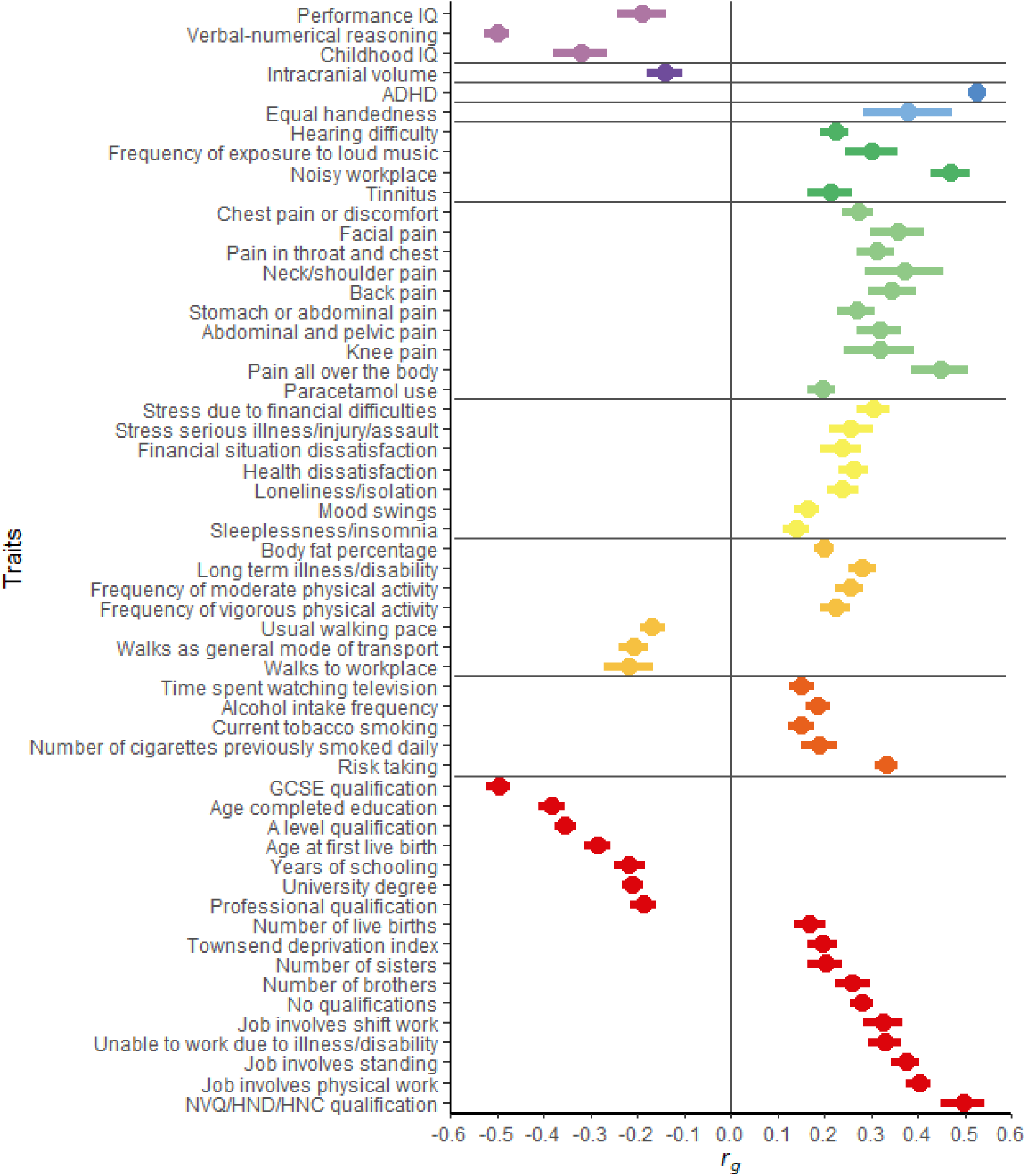
Genetic correlations of dyslexia with other phenotypes. Significant (*p* < 5 × 10^−4^) genetic correlations (*r*_g_) between self-reported dyslexia diagnosis from 23andMe and other phenotypes from the LD Hub database and Enhancing Neuro Imaging Genetic Through Meta-Analysis (ENIGMA). Ninety-eight traits were tested but only those that were significant after Bonferroni correction are presented. Points represent genetic correlation and error bars represent 95% confidence limits. The vertical dotted line indicates a genetic correlation of zero and the horizontal dotted lines divide groups of related traits.

### Mendelian randomisation

We investigated potential causal relationships between dyslexia and a selection of other relevant traits. Two-sample inverse variance weighted Mendelian Randomisation (MR) analyses were compatible with a causal effect of dyslexia on ADHD (OR = 1.43, 95% CI: 1.28-1.61) and adult general cognitive ability (β = -0.21, *SE* = 0.02) (Supplementary Figure 13 and Supplementary Table 24 for complete results). We detected a small causal effect of dyslexia on secondary school achievement (β = -0.03, *SE* = 0.01), but inconsistent support across inverse variance weighted MR, MR Egger, weighted median MR and weighted mode MR for educational attainment and income. Because ADHD and general cognitive ability can precede dyslexia, this directional path was tested, but was only significant for cognitive ability. The few available SNPs forming an instrument for ADHD likely biased that test towards the null, so we cannot rule out a bidirectional link. In tests for directional pleiotropy, Egger intercepts did not significantly differ from zero, supporting the assumption of no direct effect from the instrument to outcome. However, there may be confounding from dynastic, assortative mating and population stratification effects that could not be tested in this study.

### Polygenic score analyses

Dyslexia polygenic scores (PGS) based on the 23andMe dyslexia GWAS were computed in four independent cohorts, and overall, higher PGS were associated with lower reading and spelling accuracy (Supplementary Table 25). In two Australian population-based samples (1,647 adolescents, 1,163 adults), the dyslexia PGS explained up to 3.6% of variance in nonword reading (phonological decoding). Dyslexia PGS did not correlate with nonword repetition (phonological short-term memory). In developmental cohorts enriched for reading difficulties, the dyslexia PGS explained 3.7% (UKdys; N=930) and 5.6% (CLDRC; N=717) of variance in word recognition tests.

### Analyses of dyslexia associations from the literature

Of 75 previously reported dyslexia associations, none showed genome-wide significance in our analyses (Supplementary Table 26). Nineteen of these targeted variants—in *ATP2C2, CMIP, CNTNAP2, DCDC2, DIP2A, DYX1C1, FOXP2, KIAA0319L* and *PCNT*—showed association surviving Bonferroni-correction which accounted for LD (*p* < .05/68.7). In gene-based tests of 14 candidate genes from literature [25, 26] association at a Bonferroni level (p < .05/14) was seen for *KIAA0319L* (*p* = 1.84 × 10^−4^) and *ROBO1* (*p* = 1.53 × 10^−3^) (Supplementary Table 27). *CNTNAP2* was marginal (*p* = .004). Targeted gene-set analysis of three pathways previously implicated in dyslexia (Supplementary Table 28) showed replication-level support (*p* = 2.00 × 10^−3^) for the axon guidance pathway (comprising 216 genes).

## DISCUSSION

In the largest GWAS of dyslexia to date (>50,000 self-reported diagnoses), we identified 42 significant independent loci. Twenty-five of these represent novel associations that have not been uncovered in GWAS of related cognitive traits; 11 of these were validated in the GenLang consortium GWAS meta-analysis of reading/spelling in English and other European languages, and another in a Chinese language cohort. 38% of the significant SNPs overlap with variants from general cognitive ability GWAS, consistent with twin studies that find genetic variation in reading disability is explained by general and reading-specific cognitive ability [13]. Similar to other complex traits, each significant locus showed small effects (ORs ranging 1.04 to 1.12), consistent with high polygenicity. Our estimated SNP-based heritability of 15% (5% dyslexia prevalence) was similar to the 19% reported in a smaller GWAS [5], but lower than heritability estimates from twin studies (40-80%) [27, 28]. This difference may be partly due to effects of rare and structural variants [29], which have been implicated in reading and related traits [30, 31].

Whereas *AUTS2* has been implicated in autism [32], intellectual disability [33], and dyslexia [34], the variant we uncover (rs3735260) represents the strongest *AUTS2* SNP association with a neurodevelopmental trait to date. Amongst our findings were other known neurodevelopmental genes, like *TANC2* (implicated in language delay and intellectual disability [35, 36]), and especially *GGNBP2* (linked to neurodevelopmental delay [37] and autism [38]) with its variant rs34349354 supported in all our validation cohorts. However, rs34349354 is pleiotropic with general cognitive ability, and based on eQTL evidence is more likely linked to *ZNHIT3*, associated with cognitive performance and co-localising with molecular QTLs [16]. Notably, none of the more established candidate genes for dyslexia approached genome-wide significance in our results.

Consistent with studies of other neurodevelopmental disorders [39, 40], partitioning of SNP-based heritability revealed enrichment in conserved regions, and at H3K4me1 and H3K4me3 clusters in the CNS (marking enhancers and promoters respectively). Since reading/writing systems are built on our capacities for spoken language, it is plausible that evolutionary changes on the human lineage helped shape the underlying genetic architecture [41]. However, we did not find enrichment of significant associations for curated annotations spanning different periods of hominin prehistory.

Our self-reported dyslexia diagnosis binary trait showed strong negative genetic correlations with quantitative reading and spelling measures, supporting the validity of this measure in the 23andMe cohort, and suggesting that reading skills and disorder are not qualitatively distinct. The positive genetic correlation between hearing difficulties and dyslexia is consistent with genetic correlations reported for childhood reading skill [42], suggesting that hearing problems at an early age could affect acquisition of phonological processing skills.

Dyslexia showed moderately negative genetic correlations with adult verbal-numerical reasoning. MR demonstrated a moderate causal effect of dyslexia on general cognitive ability and a larger reverse causal effect, highlighting the importance of general cognitive processes in reading acquisition. The lack of a strong genetic correlation of dyslexia with (nonverbal) performance IQ suggests that the causal effect is on verbal ability and that individuals with dyslexia are disadvantaged on verbal IQ tests [43]. This putative causal effect was less pronounced for secondary school achievement (perhaps due to school adjustments/support) and not robustly supported for college/university education.

There was little support for common genetic variation in dyslexia being related to inter-individual differences in subcortical volumes, or structural connectivity and morphometry for brain regions implicated in language processing in adults. Thus, the phenotypic correlations previously reported between dyslexia and aspects of neuroanatomy may in large part reflect environmental shaping of the brain, perhaps through the process of reading itself [44]. Left-handedness and ambidexterity show little genetic overlap with each other [45] yet are both phenotypically linked to neurodevelopmental disorders/cognitive abilities [46, 47]. We report a significant genetic correlation between dyslexia and self-reported equal hand use, but not left-handedness, supporting the ambidexterity theory [48].

Dyslexia and ADHD [8, 9] are comorbid (24% reporting ADHD in our cases versus 9% in controls), and we show a moderate genetic correlation, potentially reflecting shared endophenotypes like deficits in working memory and attention [49]. Our MR finding of a causal effect of dyslexia on ADHD implies that a reduction in dyslexia prevalence (achievable through remediation) could reduce ADHD prevalence. However, this result is tentative given the potential for unmeasured confounding with the genetic instrument. Although dyslexia and ASD were not genetically correlated, the latter encompasses diverse neurodevelopmental phenotypes including sub-groups with varying educational attainment and IQ [40]. Unexpected genetic correlations with pain-related traits suggest that individuals with dyslexia may have a lower threshold for pain perception. Links between pain and other neurodevelopmental disorders have been reported [50].

Dyslexia polygenic scores were correlated with lower achievement on reading and spelling tests in population-based and reading-disorder enriched samples, especially for nonword reading, a measure of phonological decoding typically impaired in dyslexia. Polygenic scores could become a valuable tool to help identify children with a propensity for dyslexia, enabling learning support prior to development of reading skills.

In summary, we report 42 novel independent significant loci associated with dyslexia, 25 of which have not been associated with cognitive-educational traits and should be prioritised for follow up as dyslexia candidates. Functional annotation highlights conserved and enhancer regions of the genome. Dyslexia shows positive genetic correlations with ADHD, vocational qualifications, physical occupations, ambidexterity, and pain perception, and negative correlations with academic qualifications and cognitive ability; family-based methods are needed to dissociate pleiotropic and causal effects.

## Supporting information

GenLang Consortium Author List and Acknowledgements

## Data Availability

The full summary statistics for each dyslexia GWAS presented in this paper will be made available through 23andMe to qualified researchers under an agreement with 23andMe that protects the privacy of the 23andMe participants. Interested investigators should and reference this paper for more information.

## ACKNOWLEDGEMENTS

We thank the research participants and employees of 23andMe Inc, the GenLang Consortium, the Brisbane Adults Reading Study, and the Chinese Reading Study.

The following members of the 23andMe Research Team contributed to this study: Stella Aslibekyan, Adam Auton, Elizabeth Babalola, Robert K. Bell, Jessica Bielenberg, Katarzyna Bryc, Emily Bullis, Daniella Coker, Gabriel Cuellar Partida, Devika Dhamija, Sayantan Das, Sarah L. Elson, Teresa Filshtein, Kipper Fletez-Brant, Pierre Fontanillas, Will Freyman, Pooja M. Gandhi, Karl Heilbron, Barry Hicks, David A. Hinds, Ethan M. Jewett, Yunxuan Jiang, Katelyn Kukar, Keng-Han Lin, Maya Lowe, Jey McCreight, Matthew H. McIntyre, Steven J. Micheletti, Meghan E. Moreno, Joanna L. Mountain, Priyanka Nandakumar, Elizabeth S. Noblin, Jared O’Connell, Aaron A. Petrakovitz, G. David Poznik, Morgan Schumacher, Anjali J. Shastri, Janie F. Shelton, Jingchunzi Shi, Suyash Shringarpure, Vinh Tran, Joyce Y. Tung, Xin Wang, Wei Wang, Catherine H. Weldon, Peter Wilton, Alejandro Hernandez, Corinna Wong, Christophe Toukam Tchakouté

## ETHICAL STANDARDS

Participants provided informed consent and participated in the research online, under a protocol approved by the external AAHRPP-accredited IRB, Ethical & Independent Review Services (E&I Review). Participants were included in the analysis on the basis of consent status as checked at the time data analyses were initiated.

## REFERENCES

1. Katusic, S.K., et al., Incidence of reading disability in a population-based birth cohort, 1976-1982, Rochester, Minn. Mayo Clin Proc, 2001. 76(11): p. 1081–92.

2. Shaywitz, S.E., et al., Prevalence of Reading Disability in Boys and Girls: Results of the Connecticut Longitudinal Study. JAMA, 1990. 264(8): p. 998–1002.

3. Hensler, B.S., et al., Behavioral genetic approach to the study of dyslexia. Journal of developmental and behavioral pediatrics : JDBP, 2010. 31(7): p. 525–532.

4. Hawke, J.L., S.J. Wadsworth, and J.C. DeFries, Genetic influences on reading difficulties in boys and girls: the Colorado twin study. Dyslexia, 2006. 12(1): p. 21–9.

5. Gialluisi, A., et al., Genome-wide association study reveals new insights into the heritability and genetic correlates of developmental dyslexia. Molecular Psychiatry, 2020.

6. Ritchie, S.J. and T.C. Bates, Enduring links from childhood mathematics and reading achievement to adult socioeconomic status. Psychol Sci, 2013. 24(7): p. 1301–8.

7. Carroll, J.M., et al., Literacy difficulties and psychiatric disorders: evidence for comorbidity. J Child Psychol Psychiatry, 2005. 46(5): p. 524–32.

8. Margari, L., et al., Neuropsychopathological comorbidities in learning disorders. BMC Neurology, 2013. 13(1): p. 198.

9. Willcutt, E.G., B.F. Pennington, and J.C. DeFries, Twin study of the etiology of comorbidity between reading disability and attention-deficit/hyperactivity disorder. Am J Med Genet, 2000. 96(3): p. 293–301.

10. McArthur, G.M., et al., On the “specifics” of specific reading disability and specific language impairment. J Child Psychol Psychiatry, 2000. 41(7): p. 869–74.

11. Catts, H.W., et al., A longitudinal investigation of reading outcomes in children with language impairments. J Speech Lang Hear Res, 2002. 45(6): p. 1142–57.

12. Bates, T.C., et al., Genetic and environmental bases of reading and spelling: A unified genetic dual route model. Reading and Writing, 2007. 20(20): p. 147–171.

13. Haworth, C.M.A., et al., Generalist genes and learning disabilities: a multivariate genetic analysis of low performance in reading, mathematics, language and general cognitive ability in a sample of 8000 12-year-old twins. Journal of Child Psychology and Psychiatry, 2009. 50(10): p. 1318–1325.

14. Fisher, S.E. and J.C. DeFries, Developmental dyslexia: genetic dissection of a complex cognitive trait. Nat Rev Neurosci, 2002. 3(10): p. 767–80.

15. Buniello, A., et al., The NHGRI-EBI GWAS Catalog of published genome-wide association studies, targeted arrays and summary statistics 2019. Nucleic Acids Research, 2018. 47(D1): p. D1005–D1012.

16. Lee, J.J., et al., Gene discovery and polygenic prediction from a genome-wide association study of educational attainment in 1.1 million individuals. Nat Genet, 2018. 50(8): p. 1112–1121.

17. Zhu, Z., et al., Causal associations between risk factors and common diseases inferred from GWAS summary data. Nature Communications, 2018. 9(1): p. 224.

18. Kircher, M., et al., A general framework for estimating the relative pathogenicity of human genetic variants. Nature genetics, 2014. 46(3): p. 310–315.

19. Ernst, J. and M. Kellis, ChromHMM: automating chromatin-state discovery and characterization. Nature Methods, 2012. 9(3): p. 215–216.

20. Kundaje, A., et al., Integrative analysis of 111 reference human epigenomes. Nature, 2015. 518(7539): p. 317–30.

21. Tilot, A.K., et al., The Evolutionary History of Common Genetic Variants Influencing Human Cortical Surface Area. Cerebral Cortex, 2020. 31(4): p. 1873–1887.

22. Sniekers, S., et al., Genome-wide association meta-analysis of 78,308 individuals identifies new loci and genes influencing human intelligence. Nat Genet, 2017. 49(7): p. 1107–1112.

23. Bycroft, C., et al., The UK Biobank resource with deep phenotyping and genomic data. Nature, 2018. 562(7726): p. 203–209.

24. Middeldorp, C.M., et al., A Genome-Wide Association Meta-Analysis of Attention-Deficit/Hyperactivity Disorder Symptoms in Population-Based Pediatric Cohorts. J Am Acad Child Adolesc Psychiatry, 2016. 55(10): p. 896-905.e6.

25. Luciano, M., et al., The Influence of Dyslexia Candidate Genes on Reading Skill in Old Age. Behavior Genetics, 2018. 48(5): p. 351–360.

26. Doust, C., et al., The Association of Dyslexia and Developmental Speech and Language Disorder Candidate Genes with Reading and Language Abilities in Adults. Twin Research and Human Genetics, 2020. 23(1): p. 23–32.

27. Davis, C.J., et al., Genetics and environmental influences on rapid naming and reading ability. Annals of Dyslexia, 2001. 51: p. 231–247.

28. Gayán, J. and R.K. Olson, Genetic and environmental influences on orthographic and phonological skills in children with reading disabilities. Dev Neuropsychol, 2001. 20(2): p. 483–507.

29. Hannula-Jouppi, K., et al., The axon guidance receptor gene ROBO1 is a candidate gene for developmental dyslexia. PLoS Genet, 2005. 1(4): p. e50.

30. Ganna, A., et al., Ultra-rare disruptive and damaging mutations influence educational attainment in the general population. Nature neuroscience, 2016. 19(12): p. 1563–1565.

31. Gialluisi, A., et al., Investigating the effects of copy number variants on reading and language performance. Journal of neurodevelopmental disorders, 2016. 8: p. 17–17.

32. Oksenberg, N., et al., Function and Regulation of AUTS2, a Gene Implicated in Autism and Human Evolution. PLOS Genetics, 2013. 9(1): p. e1003221.

33. Beunders, G., et al., Two male adults with pathogenic AUTS2 variants, including a two-base pair deletion, further delineate the AUTS2 syndrome. European Journal of Human Genetics, 2015. 23(6): p. 803–807.

34. Girirajan, S., et al., Relative Burden of Large CNVs on a Range of Neurodevelopmental Phenotypes. PLOS Genetics, 2011. 7(11): p. e1002334.

35. Wessel, K., et al., 17q23.2q23.3 de novo duplication in association with speech and language disorder, learning difficulties, incoordination, motor skill impairment, and behavioral disturbances: a case report. BMC Med Genet, 2017. 18(1): p. 119.

36. Guo, H., et al., Disruptive mutations in TANC2 define a neurodevelopmental syndrome associated with psychiatric disorders. Nature Communications, 2019. 10(1): p. 4679.

37. Pasmant, E., et al., Characterization of a 7.6-Mb germline deletion encompassing the NF1 locus and about a hundred genes in an NF1 contiguous gene syndrome patient. Eur J Hum Genet, 2008. 16(12): p. 1459–66.

38. Takata, A., et al., Integrative Analyses of De Novo Mutations Provide Deeper Biological Insights into Autism Spectrum Disorder. Cell Reports, 2018. 22(3): p. 734–747.

39. Demontis, D., et al., Discovery of the first genome-wide significant risk loci for attention deficit/hyperactivity disorder. Nature Genetics, 2019. 51(1): p. 63–75.

40. Grove, J., et al., Identification of common genetic risk variants for autism spectrum disorder. Nature Genetics, 2019. 51(3): p. 431–444.

41. Mozzi, A., et al., The evolutionary history of genes involved in spoken and written language: beyond FOXP2. Scientific Reports, 2016. 6(1): p. 22157.

42. Schmitz, J., F. Abbondanza, and S. Paracchini, Genome-wide association study and polygenic risk score analysis for hearing measures in children. bioRxiv, 2021: p. 2020.07.22.215376.

43. Vellutino, F., Alternative Conceptualizations of Dyslexia: Evidence in Support of a Verbal-Deficit Hypothesis. Harvard Educational Review, 2012. 47(3): p. 334–354.

44. Dehaene, S., et al., Illiterate to literate: behavioural and cerebral changes induced by reading acquisition. Nat Rev Neurosci, 2015. 16(4): p. 234–44.

45. Cuellar-Partida, G., et al., Genome-wide association study identifies 48 common genetic variants associated with handedness. Nature Human Behaviour, 2021. 5(1): p. 59–70.

46. Papadatou-Pastou, M., et al., Human handedness: A meta-analysis. Psychological Bulletin, 2020. 146(6): p. 481–524.

47. Peters, M., S. Reimers, and J.T. Manning, Hand preference for writing and associations with selected demographic and behavioral variables in 255,100 subjects: The BBC internet study. Brain and Cognition, 2006. 62(2): p. 177–189.

48. Brandler, W.M. and S. Paracchini, The genetic relationship between handedness and neurodevelopmental disorders. Trends in molecular medicine, 2014. 20(2): p. 83–90.

49. Willcutt, E.G., et al., Neuropsychological analyses of comorbidity between reading disability and attention deficit hyperactivity disorder: in search of the common deficit. Dev Neuropsychol, 2005. 27(1): p. 35–78.

50. Whitney, D.G. and D.N. Shapiro, National Prevalence of Pain Among Children and Adolescents With Autism Spectrum Disorders. JAMA Pediatrics, 2019. 173(12): p. 1203–1205.

