## Supplementary material for "Discovery of 42 Genome-Wide Significant Loci Associated with Dyslexia": Methods

Catherine Doust^1^, Pierre Fontanillas^2^, Else Eising^3^, Scott D Gordon^4^, Zhengjun Wang^5^, Gökberk Alagöz^3^, Barbara Molz^3^, 23andMe Research Team^2^, Quantitative Trait Working Group of the GenLang Consortium, Beate St Pourcain^3,6^, Clyde Francks^3,6^, Riccardo E Marioni^7^, Jingjing Zhao^5^, Silvia Paracchini^8^, Joel B Talcott^9^, Anthony P Monaco^10^, John F Stein^11^, Jeffrey R Gruen^12^, Richard K Olson^13^, Erik G Willcutt^13^, John C DeFries^13^, Bruce F Pennington^14^, Shelley D Smith^15^, Margaret J Wright^16^, Nicholas G Martin^4^, Adam Auton^2^, Timothy C Bates^1^, Simon E Fisher^3,6^ & Michelle Luciano^1^

^1^ Department of Psychology, The University of Edinburgh, Edinburgh, EH8 9JZ, UK

^2^ 23andMe, Inc., Sunnyvale, CA, USA

^3^ Language and Genetics Department, Max Planck Institute for Psycholinguistics, 6500 AH, Nijmegen, The Netherlands

^4^ Genetic Epidemiology Laboratory, QIMR Berghofer Medical Research Institute, Brisbane, 4029, Queensland, Australia

^5^ School of Psychology, Shaanxi Normal University and Shaanxi Key Research Center of Child Mental and Behavioral Health, Xi’an, 710062, China

^6^ Donders Institute for Brain, Cognition and Behaviour, 6500 EH, Nijmegen, The Netherlands

^7^ Centre for Genomic & Experimental Medicine, Institute of Genetics & Cancer, University of Edinburgh, Edinburgh, EH4 2XU, UK

^8^ School of Medicine, Medical & Biological Sciences, University of St Andrews, St Andrews, KY16 9TF, UK

^9^ Developmental Cognitive Neuroscience, Aston Brain Centre, Birmingham, B4 7ET, UK

^10^ Office of the President, Tufts University, Medford, Massachusetts, 02155, USA

^11^ Department of Physiology, Anatomy and Genetics, Oxford University, **Oxford, OX1 3PT, UK**

^12^ Departments of Pediatrics and Genetics, Yale Medical School, New Haven, Connecticut, 06520-8081, USA

^13^ Department of Psychology and Neuroscience, University of Colorado, Boulder, Colorado, 80309-0345, USA

^14^ Department of Psychology at the University of Denver, Denver, Colorado, 80208, USA

^15^ Department of Neurological Sciences, College of Medicine, University of Nebraska Medical Center, Omaha, Nebraska, 68198-8440, USA

^16^ Queensland Brain Institute, University of Queensland, Brisbane, 4072, Queensland, Australia

### **GWAS Participants**

Participants were drawn from the customer base of 23andMe, Inc., a consumer genetics company. Participants provided informed consent and participated in the research online, under a protocol approved by the external AAHRPP-accredited IRB, Ethical & Independent Review Services ([www.eandireview.com](http://www.eandireview.com)). They included 51,800 (21,513 male, 30,287 female) participants who responded “yes” to the question “Have you been diagnosed with dyslexia?” (cases) and 1,087,070 (446,054 male, 641,016 female) participants who responded “no” (controls). Age ranged from 18 to 110 years, with the prevalence of dyslexia highest for younger participants (5.34% in those aged 20 to 30 years) than older participants (3.23% in those aged 80 to 90 years). The negative linear relationship between dyslexia prevalence and participant age was expected given that screening for specific learning difficulties has only become commonplace in more recent decades. Moreover, this aligns with findings from the subsample (4.3%) of participants who reported age of diagnosis: younger participants were diagnosed at an earlier age (e.g., 9.7 years (±4.7) for 20- to 30-year-olds) than older participants (e.g., 22.4 years (±17.8) for 80- to 90-year-olds). The prevalence of dyslexia in our sample was similar for women (4.51%) and men (4.6%), although the slightly higher prevalence in males in this very large sample was statistically significant (*p* < 8.7 x 10^-6^). Such a prevalence lies at the lower end of the range typically reported in the US population [1] and might represent the more severe cases of dyslexia given that a formal diagnosis was required; additionally, people with dyslexia might opt out of survey research which requires reading, further restricting the sample range.

### **Genotyping and imputation**

DNA was extracted from saliva samples and genotyped on one of five genotyping platforms by National Genetics Institute (NGI). In the present analysis, only those with European ancestry were included. Details about the genotyping arrays, quality control of samples, and ancestry derivation can be found in Fontanillas et al., [2] and the Supplementary Methods. Phased genotypes were imputed to a combined reference panel of the 1000 Genomes Phase 3 haplotypes (May 2015) and the UK10K imputation reference panel using Minimac3 (see Das et al., [3]).

### **Statistical analyses**

#### **Genome-wide associations and downstream analyses**

##### **Association analysis**

Association analysis was performed on genotyped and imputed SNP dosage data using logistic regression and assuming an additive model of allelic effects. For X-chromosome analysis, male genotypes were treated as homozygous diploid. Covariates included age, age squared, gender, the first five ancestry principal components, and genotype platform. SNP significance was evaluated by a likelihood ratio test and genome-wide significance was determined as *p* < 5 × 10^-8^ (suggestive significance level as *p* < 1 × 10^-6^). Only reliably imputed SNPs (*r*^2^ > .80) and those with minor allele frequency (MAF) > .01 are presented (*N* = 7,995,923). Subsidiary GWA analysis of separate male (*n* = 21,513 cases, 446,054 controls) and female (*n* = 30,287 cases, 641,016 controls) groups, and younger (below 55 years; *n* = 30,763 cases, 582,276 controls) and older (55 and above; *n* = 21,037 cases, 504,794 controls) groups was performed. The latter was to check whether reliability of diagnosis (assumed to be higher in the younger sample whose recall of diagnosis should be better and who would have been exposed to greater levels of dyslexia screening) affected the GWAS signal.

We also looked to independently validate our genome-wide significant variants within 1) a published GWAS meta-analysis of 2,274 dyslexia cases from nine European countries representing six different languages (NeuroDys) by Gialluisi et al., [4], 2) a population sample (Chinese Reading Study, CRS) of children measured on quantitative traits of reading accuracy and reading fluency (*N* = 2,270; described in Supplementary Material), and 3) within the GenLang quantitative trait GWAS meta-analysis of word reading (up to *N* = 33,959) and spelling (up to *N* = 18,514) skills measured in cohorts of children and adolescents from Europe, USA, and Australia, and representing eight European languages of which English was the most common (Eising *et al*., in preparation).

##### **Gene-based analyses**

The GWAS results were used to calculate gene-based p-values for association with dyslexia by performing the gene analysis in MAGMA v1.08 [5] through the FUMA interface [6] using standard settings. In total, 19,039 genes were tested and *p* values were judged based on a Bonferroni-corrected significance threshold of *p* < 2.63 x 10^-6^. We also performed gene-set analyses for association of biological pathways (all available gene ontology (GO) terms and curated gene sets from the Molecular Signatures Database (MsigDB) [7, 8]) with dyslexia in MAGMA through the FUMA interface. The total number of pathways tested was 15,486 and *p* values were judged based on a Bonferroni-corrected significance threshold of *p* < 3.23 x 10^-6^.

##### **Biological annotations**

Genome-wide significant variants and nearby gene(s) were annotated using external reference data and evaluated for functional or regulatory impact. A 99% credible set of potentially causal variants for SNPs in significant regions was based on Approximate Bayes Factor (ABFs) [9] assuming a prior variance of 0.1, and using Maller et al.’s [10] method to define these sets. Variant effect prediction of these was done in ENSEMBL (release 104; [11]). For genome-wide significant variants we considered: gene context (whether a variant is intergenic or located within a specific functional region within a gene locus); deleteriousness (Combined Annotation Dependent Depletion (CADD) score), functionality (RegulomeDB (RDB) category); chromatin state (minimum and common 15-core chromatin state); and SNP-trait associations reported in the GWAS Catalog. Pleiotropic SNPs for dyslexia and general cognitive ability (which can affect the presentation of dyslexia) were identified using HEIDI-outlier analysis [12] within the GCTA toolbox (https://cnsgenomics.com/software/gcta/#Overview) which uses a **Generalised Summary-data-based Mendelian Randomisation method to classify pleiotropic SNPs based on the divergence of the beta _dyslexia-cognitive ability_ from that expected under a causal model.**

For each variant, the most probable gene target was identified using the open target genetics portal [13] which draws on evidence from QTL and chromatin interaction experiments, functional predictions, and distance from a gene’s transcription start site. For genome-wide significant genes we considered: loss-of-function intolerance (Probability of Loss-of-function Intolerance (pLI) score); variation intolerance (Residual Variation Intolerance Score, RVIS); variation intolerance in non-coding regions (non-coding RVIS, ncRVIS); evolutionary constraint of non-coding regions (non-coding Genomic Evolutionary Rate Profiling (ncGERP) score); evolutionary constraint of protein-coding regions (protein-coding Genomic Evolutionary Rate Profiling (pcGERP) score); deleteriousness across non-coding regions (non-coding CADD (ncCADD) score); combined functionality of variants in non-coding regions (non-coding Genome-Wide Annotation of Variants (ncGWAVA) score); and expression in 12 brain tissues (Amygdala, Anterior cingulate cortex, Caudate basal ganglia, Cerebellar hemisphere, Cerebellum, Cortex, Frontal cortex, Hippocampus, Hypothalamus, Nucleus accumbens basal ganglia, Putamen basal ganglia, Substantia nigra). All annotations were obtained through FUMA [6] except RVIS, ncGERP, pcGERP, ncCADD, and ncGWAVA which were from Petrovski et al., [14]. Details of each annotation including original sources are in the Supplementary Methods.

Tissue specificity for differentially expressed genes was tested in FUMA using the pre-calculated data sets of differentially expressed genes (DEG) for tissue-specific gene expression. In these DEG sets, genes with a *p* ≤ 0.05 after Bonferroni correction and absolute log fold change ≥ 0.58 were considered differentially expressed for a given tissue compared to other genes. DEG sets were calculated in terms of up-regulation, down-regulation, and two-sided differential expression. Significant genes from genome-wide gene-based tests in MAGMA were tested against all background genes for each of the pre-calculated DEG sets. Tissues with a *p* ≤ 0.05 after Bonferroni correction are considered significantly enriched in DEG.

#### **Partitioned heritability**

We partitioned SNP heritability of dyslexia using stratified LDSC, as described by Finucane et al., [15], to determine whether SNPs which share the greatest proportion of the heritability are also clustered in specific functional categories in the genome. We performed partitioning for the 24 main functional annotations defined by Finucane et al., [15]. LD scores, regression weights, and allele frequencies are from European ancestry samples and were retrieved from: <https://alkesgroup.broadinstitute.org/LDSCORE>. Heritability estimates were considered statistically significant if the *p* value surpassed an α level of 2.08 x 10^-3^, derived by Bonferroni correction based on 24 tests.

We also estimated the enrichment for heritability of dyslexia for tissue-specific annotations, while controlling for the annotations in the baseline model, including: gene expression in three brain cell types, gene expression in 12 brain regions, and chromatin marks H3K4me1 and H3K4me3 in multiple tissues (108 and 114 respectively), since these marks are enriched at enhancers [16] and promoters [17] respectively. Enrichment is the proportion of SNP heritability divided by the proportion of SNPs. For the brain cell types, we estimated enrichment for heritability of dyslexia for genes expressed in neurons, astrocytes, and oligodendrocytes using data from Cahoy et al., [18]. Enrichments were considered statistically significant if the *p* value surpassed an α level of 0.017, derived by Bonferroni correction based on three tests. The gene expression data used to estimate the enrichment of heritability in genes expressed in certain brain regions was from the GTEx database [19] and the Bonferroni-derived α level for enrichment was 4.17 x 10^-3^ (based on 12 tests). Chromatin annotations include data from the Roadmap Epigenomics consortium [20] and EN-TEx [21, 22]. For H3K4me1, the Bonferroni-derived α level for enrichment was 4.63 x 10^-4^ (based on 108 tests) and for H3K4me3, the Bonferroni-derived α level for enrichment was 4.39 x 10^-4^ (based on 114 tests).

##### *Evolutionary annotations*

Although reading and writing is a human cultural invention, it builds on fundamental pathways involved in language processing. Therefore, we investigated whether annotations related to human evolution were significantly enriched for heritability of dyslexia by applying an evolutionary analysis pipeline adapted from Tilot et al., [23]. These analyses capture a range of periods in an evolutionary timeframe on the lineage that led to humans, from approximately 30 million years ago to 50,000 years ago.

Enrichment of heritability was estimated in adult brain Human Gained Enhancers (HGEs) [24], foetal brain HGEs [25], ancient selective sweep regions [26], Neanderthal-introgressed SNPs [27] and Neanderthal-depleted regions [28] (see Supplementary Materials for a description of each annotation); and controlled for using the baselineLD v2 model from Finucane et al., [15]. Heritability enrichment in human adult and foetal HGEs were additionally controlled for adult and foetal brain active regulatory elements from the Roadmap Epigenomics resource [20]. Active regulatory elements were defined using chromHMM [29]. Enrichment *p* values were judged by an α level of 10^-2^, derived by Bonferroni correction based on five tests.

#### **Genetic correlations**

##### *Genetic correlations within the 23andMe GWAS of dyslexia*

Genetic correlation between self-reported dyslexia diagnosis in males and females, and between younger (< 55 years old) and older (> 55 years old) adults was calculated using LD Score regression (LDSC) [30, 31].

##### *Genetic correlations of dyslexia with other traits*

We present the pairwise genetic correlation of dyslexia with 98 traits. Summary statistics for most of these traits are publicly-available through LD Hub [30-32], a centralised database and web interface that automates the LDSC regression analysis pipeline. A selection of brain MRI measures obtained from the Enhancing Neuro Imaging Genetic through Meta-Analysis (ENIGMA-3) consortium [33-36], and measures of reading and spelling accuracy, and performance IQ from the GenLang Consortium (Eising *et al*., in preparation) were analysed locally using LDSC. Word reading accuracy in GenLang was measured by the number of correct words read aloud from a list in a time restricted or unrestricted fashion. Examples of tools that include this measure are Test of Word Reading Efficiency (TOWRE), the British Ability Scale (BAS) and the Wide Range Achievement Test (WRAT). Spelling accuracy in GenLang was measured by the number of words correctly spelled orally or in writing. The words were dictated as single words or in a sentence. Examples of tools that include this measure are the BAS, WRAT and Wechsler Objective Reading Dimensions (WORD). Performance IQ in GenLang was based on subtests of IQ tests that did not depend on verbal cues, as included for example in the BAS and Wechsler Intelligence Scale for Children (WISC). Trait descriptions and summary statistic sources are in Supplementary Table 22. Bonferroni correction for multiple testing derived an adjusted critical *p* value of 5.1 x 10^-4^ from 98 independent tests.

Genetic correlations were further estimated in a targeted analysis of structural brain MRI measures from UK Biobank, which were more comprehensive than those currently available from ENIGMA, along with further advantages such as hemisphere-specific data and greater homogeneity in cohort and scanning procedures. GWAS summary statistics from brain imaging-derived phenotypes for 33,000 participants were downloaded from the Oxford Brain Imaging Genetics Server [37]. Structural brain imaging traits encompassed both diffusion tensor imaging (DTI) and surface based morphometric (SBM) phenotypes [38] where selected tracts or regions of interests (ROIs) had a known link to language. For DTI, fractional anisotropy (FA) values derived from both tract-based-spatial statistics (TBSS) and probabilistic tractography were used for available tracts spanning the extended language network [39]. For SBM (cortical volume, surface area and thickness) GWAS summary statistics for ROIs derived from the Desikan-Killiany atlas (white surface) where used, again selected for their relevance in language processing, based on prior literature [40-43]. To correct for multiple testing, phenotypic correlations between the UK Biobank imaging indices were derived and analysed by PhenoSpD [44] to obtain the number of independent variables (36.08) to use for Bonferroni correction (adjusted critical *p* value of 1.39 x 10^-3^).

#### **Mendelian randomisation**

We examined causal effects of dyslexia on several traits of interest. These traits were: cognitive performance, defined as the first unrotated component from a principal components analysis of tests from at least three cognitive domains (*N* = 257,841) [45]; ADHD (*N* = 20,183 cases, 35,191 controls) [46]; educational achievement (Completion of Secondary school advanced level subjects (A levels/AS levels or equivalent, *N* = 123,661) versus none (*N* = 334,418), and College/University degree (*N* = 148,722) versus none (*N* = 309,357); UK Biobank; Bycroft et al., [47]), and economic position (average total household income before tax, N = 397,7510; Townsend deprivation index, N = 462,464). Causal effects were estimated by two sample mendelian randomisation (MR) using the TwoSampleMR package [48] in R. Exposure variants were selected using a significance threshold of *p* < 5 x 10^-8^, a genomic distance of < 10,000 kb, and *r*^2^ < .001 to create a genetic instrument of 51 SNPs associated with dyslexia. ADHD (genetic instrument of 10 SNPs) which is a developmental trait, and cognitive performance (genetic instrument of 103 SNPs) which is genetically stable from childhood to adulthood (r*_g_* of .71, [49]) were also tested as an exposure variable with dyslexia as an outcome. Outcome summary statistics were downloaded from the MRC Integrative Epidemiology Unit (IEU) GWAS database [50] (see Supplementary Table 24 for outcome IDs). Effects were estimated using four MR methods: MR Egger, weighted median, inverse variance weighted, and weighted mode.

#### **Polygenic score analyses**

Dyslexia polygenic scores were based on increasingly larger numbers of SNPs corresponding to their association *p* values from the 23andMe GWAS (*p* < 5 x 10^-8^, *p* < 1 x 10^-5^, *p* < .001, *p* < .01, *p* < .05, *p* < .1, *p* < .5, 1). They were calculated in four independent cohorts. Two were general population cohorts from Australia: *N* = 1,640 (772 families) adolescents/young adults (Brisbane Adolescents) [51]; *N* = 1,165 (966 families) older adults (Brisbane Adults) [52]. The other two were family-based samples selected for dyslexia, one from the UK (UKdys): *N* = 930 (595 families); the other from the USA (Colorado Learning Disabilities Research Center, CLDRC) *N* = 717 (336 families) [53]. In the Australian samples, polygenic scores were calculated on 1000 Genomes Phase 3 (version 20101123) imputed genetic data using PLINK [54]. Only reliably imputed SNPs (*R*^2^ > .80) and those with a minor allele frequency > .01 were included, and the default clumping procedure was used where index SNPs formed a clump with other SNPs in LD (*R*^2^ > .1) and within a 250 kb distance. In the UKdys and CLDRC samples, polygenic scores were calculated on HRC imputed genetic data using PRSice [55], with the same imputation quality and MAF exclusions for the base (23andMe GWA) sample, and clumping parameters.

Polygenic scores were then used as predictors in linear models of quantitative trait outcomes (Australia: word, nonword (phonetic), irregular word (lexical) reading and spelling tests from an extended version of the Components of Reading Examination [56], and two nonword repetition tests which are sensitive to developmental language disorders – Dollaghan and Campbell [57], Gathercole and Baddeley [58]; UKdys and CLDRC: word recognition). All quantitative traits were pre-adjusted for sex, age and ancestry principal components (10 PCs in UKdys and CLDR; 20 PCs in Australian samples). Further adjustments were made for imputation run (separate runs for different genotyping arrays) in the Australian samples, and for nonverbal IQ in all samples (except for the Australian Adults), and for hearing difficulties in the Australian older adults. Because the cohorts included related family members (twins or siblings), linear mixed models (lme) were specified in RStudio [59] with family membership modelled as a random effect and the dyslexia polygenic score as a fixed effect. Where MZ twins were present, their trait scores were averaged and they were used as a single case.

##### **Evaluation of candidates from prior literature**

We used the results of the 23andMe dyslexia GWAS to assess variants, genes, and biological pathways previously associated with or implicated in dyslexia and/or variation in reading and spelling ability in past association studies, linkage analyses and other studies.

###### *Previously-reported variants*

We assessed 75 previously-reported variants within our summary statistics, adopting a replication/validation significance threshold of *p* < 7.28 x 10^-4^, derived by Bonferroni correction based on 68.7 independent tests derived through matrix spectral decomposition, taking into account linkage disequilibrium (see Doust et al., [52] for details on how these variants were selected ). The sources for each variant are provided in Supplementary Table 26.

###### *Dyslexia candidate genes*

We evaluated gene-based results from MAGMA v1.08 [5] for overrepresentation of genome-wide significant variants from the 23andMe dyslexia GWAS within the loci of 14 candidate genes from prior literature: *CMIP*, *CNTNAP2*, *CYP19A1*, *DCDC2*, *DIP2A*, *DYX1C1*, *GCFC2*, *KIAA0319*, *KIAA0319L*, *MRPL19*, *PCNT*, *PRMT2*, *S100B*, and *ROBO1*. The rationale for this selection is detailed by Luciano et al., [60] and Doust et al., [52]. The critical *p* value, based on Bonferroni correction for 14 tests, was 3.57 x 10^-3^.

###### *Candidate dyslexia gene sets*

We performed a gene-set analysis in MAGMA to test for overrepresentation of genome-wide significant variants within 1) a set of transcriptional targets of *FOXP2,* a highly conserved transcription factor linked to speech and language impairment [61]; and 2) two biological pathways previously suggested to play a role in dyslexia susceptibility [62, 63]: axon guidance (GO:0007411: ‘chemotaxis process that directs the migration of an axon growth cone to a specific target site’; 216 genes) and neuron migration (GO:0001764: ‘movement of an immature neuron from germinal zones to specific positions where they will reside as they mature’; 145 genes). An adjusted critical *p* value of .017 was derived using Bonferroni correction based on three independent tests.

#### **REFERENCES**

1.	Katusic, S.K., et al., *Incidence of reading disability in a population-based birth cohort, 1976-1982, Rochester, Minn.* Mayo Clin Proc, 2001. **76**(11): p. 1081-92.

2.	Fontanillas, P., et al., *Disease risk scores for skin cancers.* Nature Communications, 2021. **12**(1): p. 160.

3.	Das, S., et al., *Next-generation genotype imputation service and methods.* Nat Genet, 2016. **48**(10): p. 1284-1287.

4.	Gialluisi, A., et al., *Genome-wide association scan identifies new variants associated with a cognitive predictor of dyslexia.* Translational Psychiatry, 2019. **9**(1): p. 77.

5.	de Leeuw, C.A., et al., *MAGMA: Generalized Gene-Set Analysis of GWAS Data.* PLoS Computational Biology, 2015. **11**(4): p. e1004219.

6.	Watanabe, K., et al., *Functional mapping and annotation of genetic associations with FUMA.* Nat Commun, 2017. **8**(1): p. 1826.

7.	Subramanian, A., et al., *Gene set enrichment analysis: A knowledge-based approach for interpreting genome-wide expression profiles.* Proceedings of the National Academy of Sciences, 2005. **102**(43): p. 15545-15550.

8.	Liberzon, A., et al., *Molecular signatures database (MSigDB) 3.0.* Bioinformatics, 2011. **27**(12): p. 1739-1740.

9.	Wakefield, J., *A Bayesian measure of the probability of false discovery in genetic epidemiology studies.* American journal of human genetics, 2007. **81**(2): p. 208-227.

10.	Maller, J.B., et al., *Bayesian refinement of association signals for 14 loci in 3 common diseases.* Nat Genet, 2012. **44**(12): p. 1294-301.

11.	Howe, K.L., et al., *Ensembl 2021.* Nucleic Acids Research, 2020. **49**(D1): p. D884-D891.

12.	Zhu, Z., et al., *Causal associations between risk factors and common diseases inferred from GWAS summary data.* Nature Communications, 2018. **9**(1): p. 224.

13.	Carvalho-Silva, D., et al., *Open Targets Platform: new developments and updates two years on.* Nucleic Acids Research, 2018. **47**(D1): p. D1056-D1065.

14.	Petrovski, S., et al., *The Intolerance of Regulatory Sequence to Genetic Variation Predicts Gene Dosage Sensitivity.* PLOS Genetics, 2015. **11**(9): p. e1005492.

15.	Finucane, H.K., et al., *Partitioning heritability by functional annotation using genome-wide association summary statistics.* Nature genetics, 2015. **47**(11): p. 1228-1235.

16.	Rada-Iglesias, A., *Is H3K4me1 at enhancers correlative or causative?* Nature Genetics, 2018. **50**(1): p. 4-5.

17.	Heintzman, N.D., et al., *Distinct and predictive chromatin signatures of transcriptional promoters and enhancers in the human genome.* Nature Genetics, 2007. **39**(3): p. 311-318.

18.	Cahoy, J.D., et al., *A Transcriptome Database for Astrocytes, Neurons, and Oligodendrocytes: A New Resource for Understanding Brain Development and Function.* The Journal of Neuroscience, 2008. **28**(1): p. 264.

19.	The GTEx Consortium, *The Genotype-Tissue Expression (GTEx) pilot analysis: Multitissue gene regulation in humans.* Science, 2015. **348**(6235): p. 648.

20.	Kundaje, A., et al., *Integrative analysis of 111 reference human epigenomes.* Nature, 2015. **518**(7539): p. 317-30.

21.	Finucane, H.K., et al., *Heritability enrichment of specifically expressed genes identifies disease-relevant tissues and cell types.* Nature Genetics, 2018. **50**(4): p. 621-629.

22.	Dunham, I., et al., *An integrated encyclopedia of DNA elements in the human genome.* Nature, 2012. **489**(7414): p. 57-74.

23.	Tilot, A.K., et al., *The Evolutionary History of Common Genetic Variants Influencing Human Cortical Surface Area.* Cerebral Cortex, 2020. **31**(4): p. 1873-1887.

24.	Vermunt, M.W., et al., *Epigenomic annotation of gene regulatory alterations during evolution of the primate brain.* Nat Neurosci, 2016. **19**(3): p. 494-503.

25.	Reilly, S.K., et al., *Evolutionary genomics. Evolutionary changes in promoter and enhancer activity during human corticogenesis.* Science, 2015. **347**(6226): p. 1155-9.

26.	Peyrégne, S., et al., *Detecting ancient positive selection in humans using extended lineage sorting.* Genome Res, 2017. **27**(9): p. 1563-1572.

27.	Simonti, C.N., et al., *The phenotypic legacy of admixture between modern humans and Neandertals.* Science, 2016. **351**(6274): p. 737-741.

28.	Vernot, B., et al., *Excavating Neandertal and Denisovan DNA from the genomes of Melanesian individuals.* Science, 2016. **352**(6282): p. 235-239.

29.	Ernst, J. and M. Kellis, *ChromHMM: automating chromatin-state discovery and characterization.* Nature Methods, 2012. **9**(3): p. 215-216.

30.	Bulik-Sullivan, B.K., et al., *LD Score regression distinguishes confounding from polygenicity in genome-wide association studies.* Nature Genetics, 2015. **47**(3): p. 291-295.

31.	Bulik-Sullivan, B.K., et al., *An atlas of genetic correlations across human diseases and traits.* Nature Genetics, 2015. **47**(11): p. 1236-1241.

32.	Zheng, J., et al., *LD Hub: a centralized database and web interface to perform LD score regression that maximizes the potential of summary level GWAS data for SNP heritability and genetic correlation analysis.* Bioinformatics, 2016. **33**(2): p. 272-279.

33.	Grasby, K.L., et al., *The genetic architecture of the human cerebral cortex.* Science, 2020. **367**(6484): p. eaay6690.

34.	Satizabal, C.L., et al., *Genetic architecture of subcortical brain structures in 38,851 individuals.* Nat Genet, 2019. **51**(11): p. 1624-1636.

35.	Hibar, D.P., et al., *Novel genetic loci associated with hippocampal volume.* Nature Communications, 2017. **8**(1): p. 13624.

36.	Adams, H.H., et al., *Novel genetic loci underlying human intracranial volume identified through genome-wide association.* Nat Neurosci, 2016. **19**(12): p. 1569-1582.

37.	Smith, S.M., et al., *Enhanced Brain Imaging Genetics in UK Biobank.* bioRxiv, 2020: p. 2020.07.27.223545.

38.	Alfaro-Almagro, F., et al., *Image processing and Quality Control for the first 10,000 brain imaging datasets from UK Biobank.* Neuroimage, 2018. **166**: p. 400-424.

39.	Forkel, S.J. and M. Catani, *The Oxford Handbook of Neurolinguistics: Diffusion Imaging Methods in Language Sciences*. 2019, Oxford: Oxford University Press.

40.	Price, C.J., *The anatomy of language: a review of 100 fMRI studies published in 2009.* Ann N Y Acad Sci, 2010. **1191**: p. 62-88.

41.	Richardson, F.M. and C.J. Price, *Structural MRI studies of language function in the undamaged brain.* Brain structure & function, 2009. **213**(6): p. 511-523.

42.	Perdue, M.V., et al., *Gray matter structure is associated with reading skill in typically developing young readers.* Cerebral Cortex, 2020. **30**(10): p. 5449-5459.

43.	Roehrich-Gascon, D., S.L. Small, and P. Tremblay, *Structural correlates of spoken language abilities: A surface-based region-of interest morphometry study.* Brain and language, 2015. **149**: p. 46-54.

44.	Zheng, J., et al. *PhenoSpD: an integrated toolkit for phenotypic correlation estimation and multiple testing correction using GWAS summary statistics*. GigaScience, 2018. **7**, DOI: 10.1093/gigascience/giy090.

45.	Lee, J.J., et al., *Gene discovery and polygenic prediction from a genome-wide association study of educational attainment in 1.1 million individuals.* Nat Genet, 2018. **50**(8): p. 1112-1121.

46.	Demontis, D., et al., *Discovery of the first genome-wide significant risk loci for attention deficit/hyperactivity disorder.* Nature Genetics, 2019. **51**(1): p. 63-75.

47.	Bycroft, C., et al., *The UK Biobank resource with deep phenotyping and genomic data.* Nature, 2018. **562**(7726): p. 203-209.

48.	Hemani, G., J. Bowden, and G. Davey Smith, *Evaluating the potential role of pleiotropy in Mendelian randomization studies.* Human Molecular Genetics, 2018. **27**(R2): p. R195-R208.

49.	Hill, W.D., et al., *Age-Dependent Pleiotropy Between General Cognitive Function and Major Psychiatric Disorders.* Biological psychiatry, 2016. **80**(4): p. 266-273.

50.	Elsworth, B., et al., *The MRC IEU OpenGWAS data infrastructure.* bioRxiv, 2020: p. 2020.08.10.244293.

51.	Luciano, M., et al., *A genome-wide association study for reading and language abilities in two population cohorts.* Genes Brain and Behavior, 2013. **12**(6): p. 645-652.

52.	Doust, C., et al., *The Association of Dyslexia and Developmental Speech and Language Disorder Candidate Genes with Reading and Language Abilities in Adults.* Twin Research and Human Genetics, 2020. **23**(1): p. 23-32.

53.	Gialluisi, A., et al., *Genome-wide screening for DNA variants associated with reading and language traits.* Genes Brain Behav, 2014. **13**(7): p. 686-701.

54.	Purcell, S., et al., *PLINK: a tool set for whole-genome association and population-based linkage analyses.* American Journal of Human Genetics, 2007. **81**(3): p. 559-575.

55.	Euesden, J., C.M. Lewis, and P.F. O'Reilly, *PRSice: Polygenic Risk Score software.* Bioinformatics, 2015. **31**(9): p. 1466-8.

56.	Bates, T.C., et al., *Behaviour genetic analyses of reading and spelling: A component processes approach.* Australian Journal of Psychology, 2004. **56**(2): p. 115-126.

57.	Dollaghan, C. and T.F. Campbell, *Nonword repetition and child language impairment.* Journal of Speech, Language, and Hearing Research, 1998. **41**(5): p. 1136-1146.

58.	Gathercole, S.E., et al., *The Children's Test of Nonword Repetition: a test of phonological working memory.* Memory, 1994. **2**(2): p. 103-27.

59.	RStudio Team, *RStudio: Integrated Development for R*. 2020: Boston, MA.

60.	Luciano, M., et al., *The Influence of Dyslexia Candidate Genes on Reading Skill in Old Age.* Behavior Genetics, 2018. **48**(5): p. 351-360.

61.	Ayub, Q., et al., *FOXP2 Targets Show Evidence of Positive Selection in European Populations.* The American Journal of Human Genetics, 2013. **92**(5): p. 696-706.

62.	Poelmans, G., et al., *A theoretical molecular network for dyslexia: integrating available genetic findings.* Molecular Psychiatry, 2011. **16**(4): p. 365-82.

63.	Guidi, L.G., et al., *The neuronal migration hypothesis of dyslexia: A critical evaluation 30 years on.* European Journal of Neuroscience, 2018. **48**(10): p. 3212-3233.
