## Supplementary Figures for "Discovery of 42 Genome-Wide Significant Loci Associated with Dyslexia"

Catherine Doust^1^, Pierre Fontanillas^2^, Else Eising^3^, Scott D Gordon^4^, Zhengjun Wang^5^, Gökberk Alagöz^3^, Barbara Molz^3^, 23andMe Research Team^2^, Quantitative Trait Working Group of the GenLang Consortium, Beate St Pourcain^3,6^, Clyde Francks^3,6^, Riccardo E Marioni^7^, Jingjing Zhao^5^, Silvia Paracchini^8^, Joel B Talcott^9^, Anthony P Monaco^10^, John F Stein^11^, Jeffrey R Gruen^12^, Richard K Olson^13^, Erik G Willcutt^13^, John C DeFries^13^, Bruce F Pennington^14^, Shelley D Smith^15^, Margaret J Wright^16^, Nicholas G Martin^4^, Adam Auton^2^, Timothy C Bates^1^, Simon E Fisher^3,6^ & Michelle Luciano^1^

^1^ Department of Psychology, The University of Edinburgh, Edinburgh, EH8 9JZ, UK

^2^ 23andMe, Inc., Sunnyvale, CA, USA

^3^ Language and Genetics Department, Max Planck Institute for Psycholinguistics, 6500 AH, Nijmegen, The Netherlands

^4^ Genetic Epidemiology Laboratory, QIMR Berghofer Medical Research Institute, Brisbane, 4029, Queensland, Australia

^5^ School of Psychology, Shaanxi Normal University and Shaanxi Key Research Center of Child Mental and Behavioral Health, Xi’an, 710062, China

^6^ Donders Institute for Brain, Cognition and Behaviour, 6500 EH, Nijmegen, The Netherlands

^7^ Centre for Genomic & Experimental Medicine, Institute of Genetics & Cancer, University of Edinburgh, Edinburgh, EH4 2XU, UK

^8^ School of Medicine, Medical & Biological Sciences, University of St Andrews, St Andrews, KY16 9TF, UK

^9^ Developmental Cognitive Neuroscience, Aston Brain Centre, Birmingham, B4 7ET, UK

^10^ Office of the President, Tufts University, Medford, Massachusetts, 02155, USA

^11^ Department of Physiology, Anatomy and Genetics, Oxford University, **Oxford, OX1 3PT, UK**

^12^ Departments of Pediatrics and Genetics, Yale Medical School, New Haven, Connecticut, 06520-8081, USA

^13^ Department of Psychology and Neuroscience, University of Colorado, Boulder, Colorado, 80309-0345, USA

^14^ Department of Psychology at the University of Denver, Denver, Colorado, 80208, USA

^15^ Department of Neurological Sciences, College of Medicine, University of Nebraska Medical Center, Omaha, Nebraska, 68198-8440, USA

^16^ Queensland Brain Institute, University of Queensland, Brisbane, 4072, Queensland, Australia

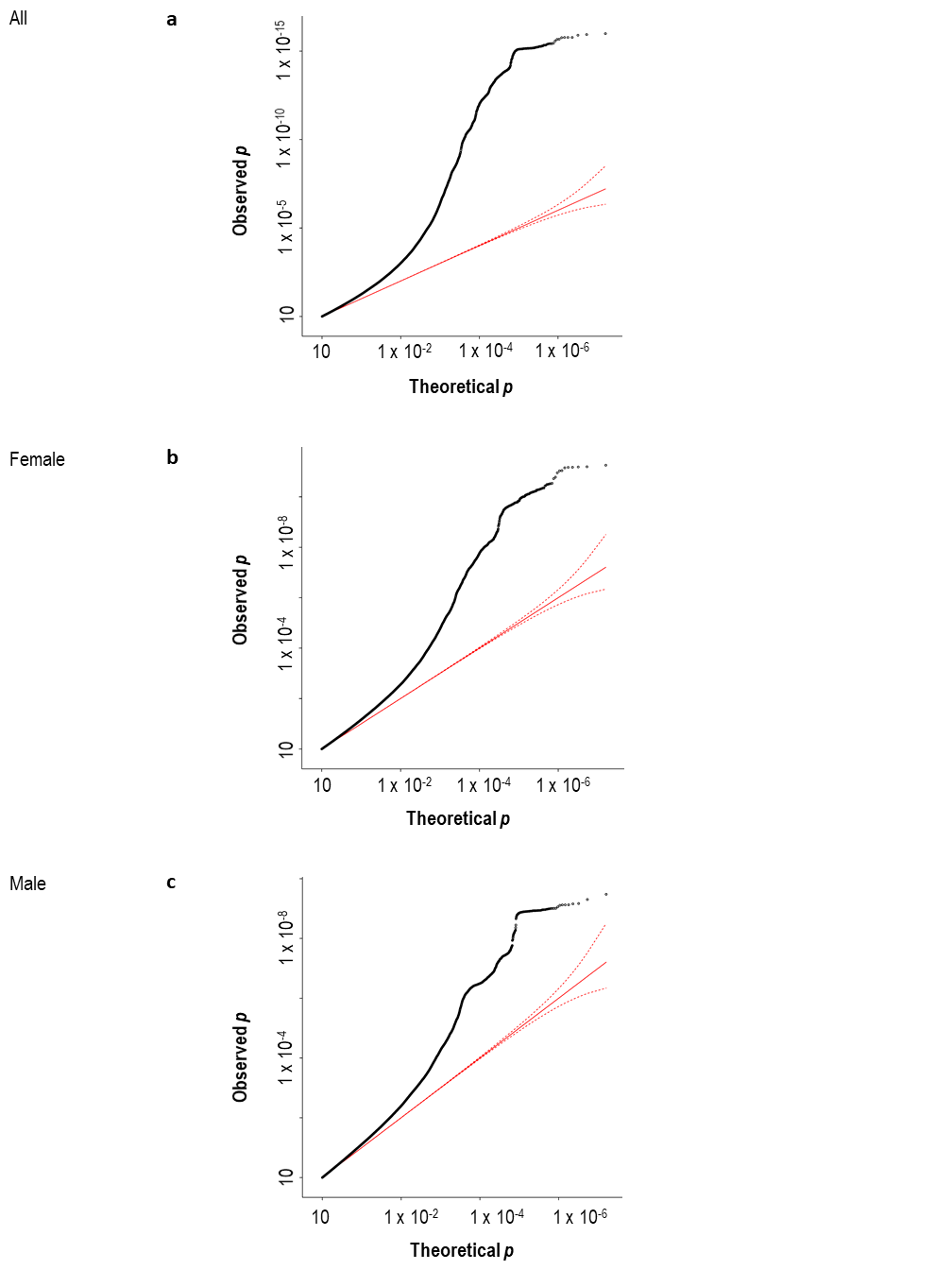

### Supplementary Figure 1 QQ plot of dyslexia GWAS results

Quantile-quantile (Q-Q) plots of observed versus expected *p* values for associations of single nucleotide polymorphisms with self-reported dyslexia diagnosis in a genome-wide association analysis for all participants (a), female participants (b), and male participants (c). The solid red line represents the distribution of *p* values under the null hypothesis and the dashed red line represent 95% confidence intervals. The black circles represent the observed distribution of p values.

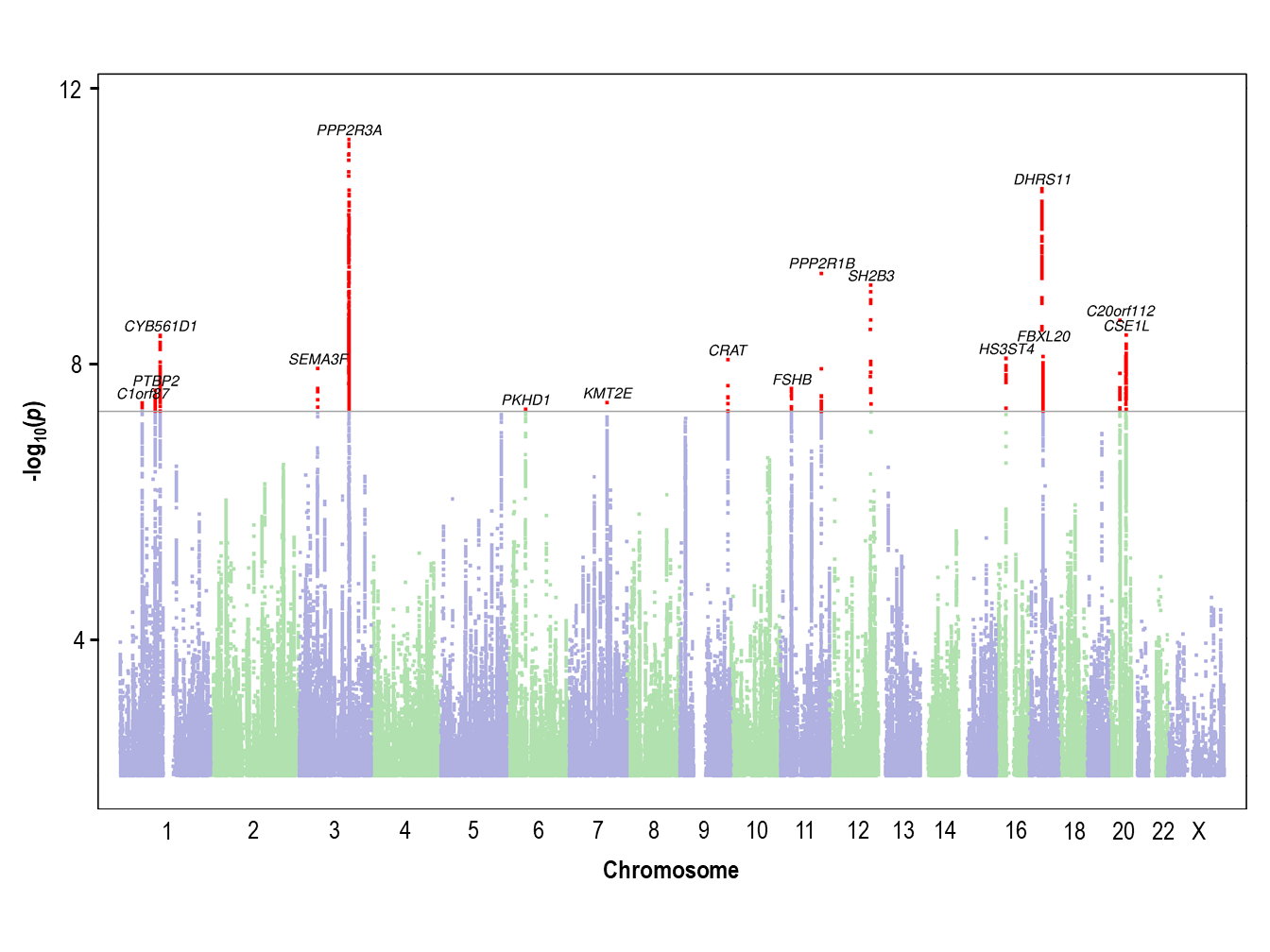

### Supplementary Figure 2 Manhattan plot of dyslexia GWAS results for females

The *y* axis represents the negative log_10_ *p* value for association of single nucleotide polymorphisms with self-reported dyslexia diagnosis from 30,287 female individuals and 641,016 female controls. The threshold for genome-wide significance (*p* < 5 x 10^-8^) is represented by a horizontal purple line. Genome-wide significant variants in the 17 genome-wide significant loci are red. Variants located within a distance of 250 kb of each other are considered as one locus.

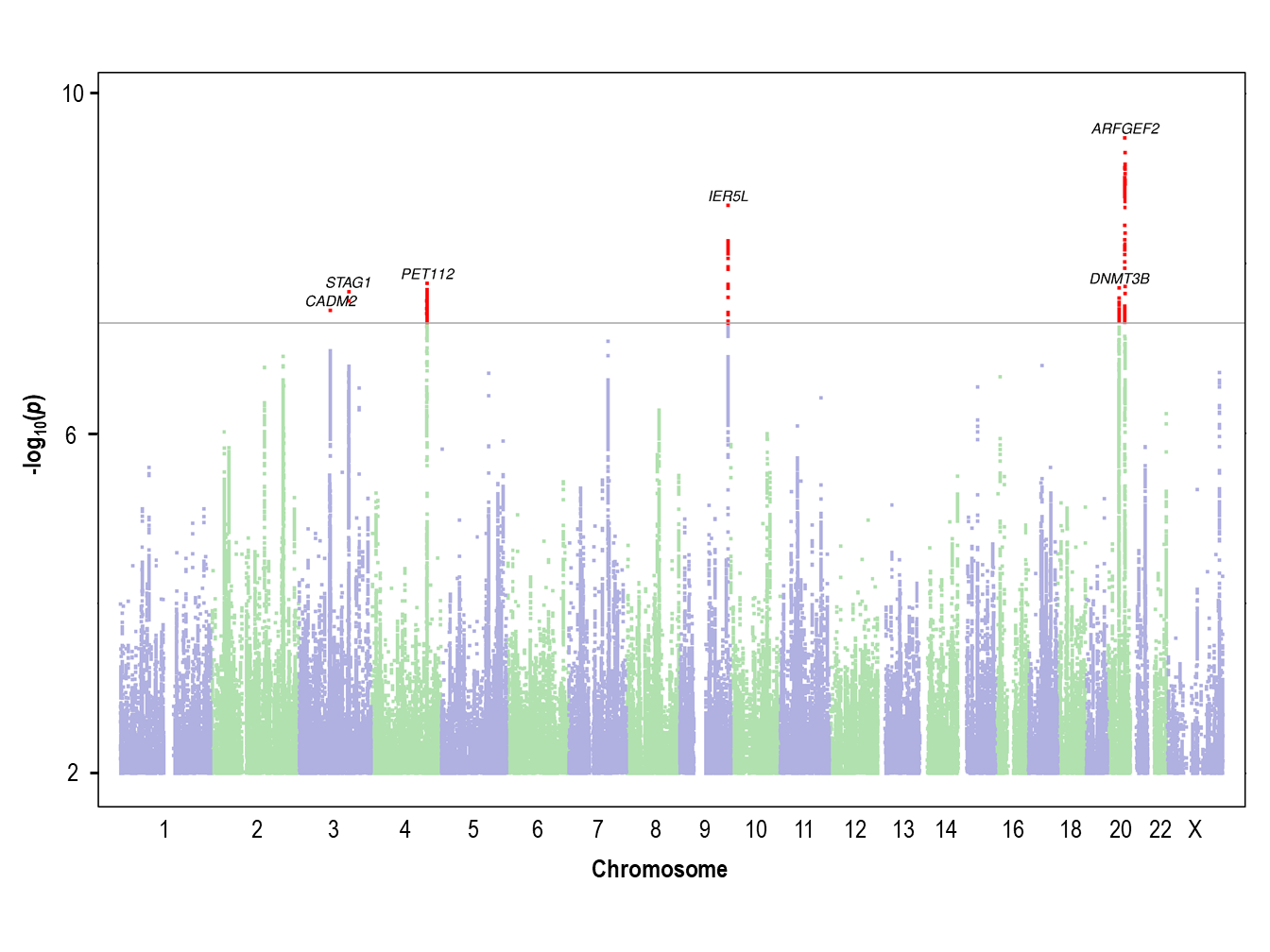

### Supplementary Figure 3 Manhattan plot of dyslexia GWAS results for males

The *y* axis represents the negative log_10_ *p* value for association of single nucleotide polymorphisms with self-reported dyslexia diagnosis from 21,513 male individuals and 446,054 male controls. The threshold for genome-wide significance (*p* < 5 x 10^-8^) is represented by a horizontal purple line. Genome-wide significant variants in the 6 genome-wide significant loci are red. Variants located within a distance of 250 kb of each other are considered as one locus.

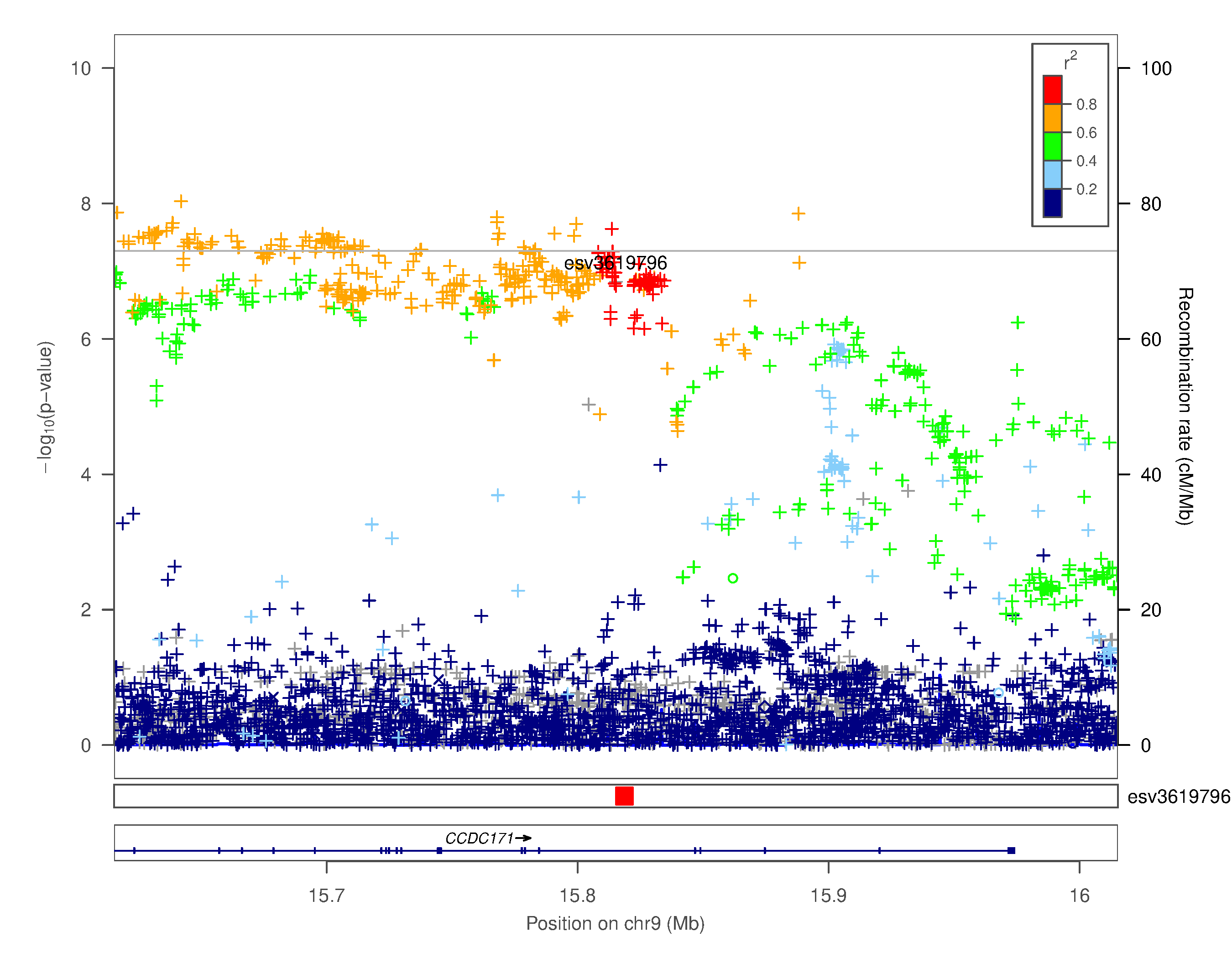
 **Supplementary Figure 4.i. Regional association** plots **for chr9p22.3 esv3619796 structural variant nearby rs3122702**

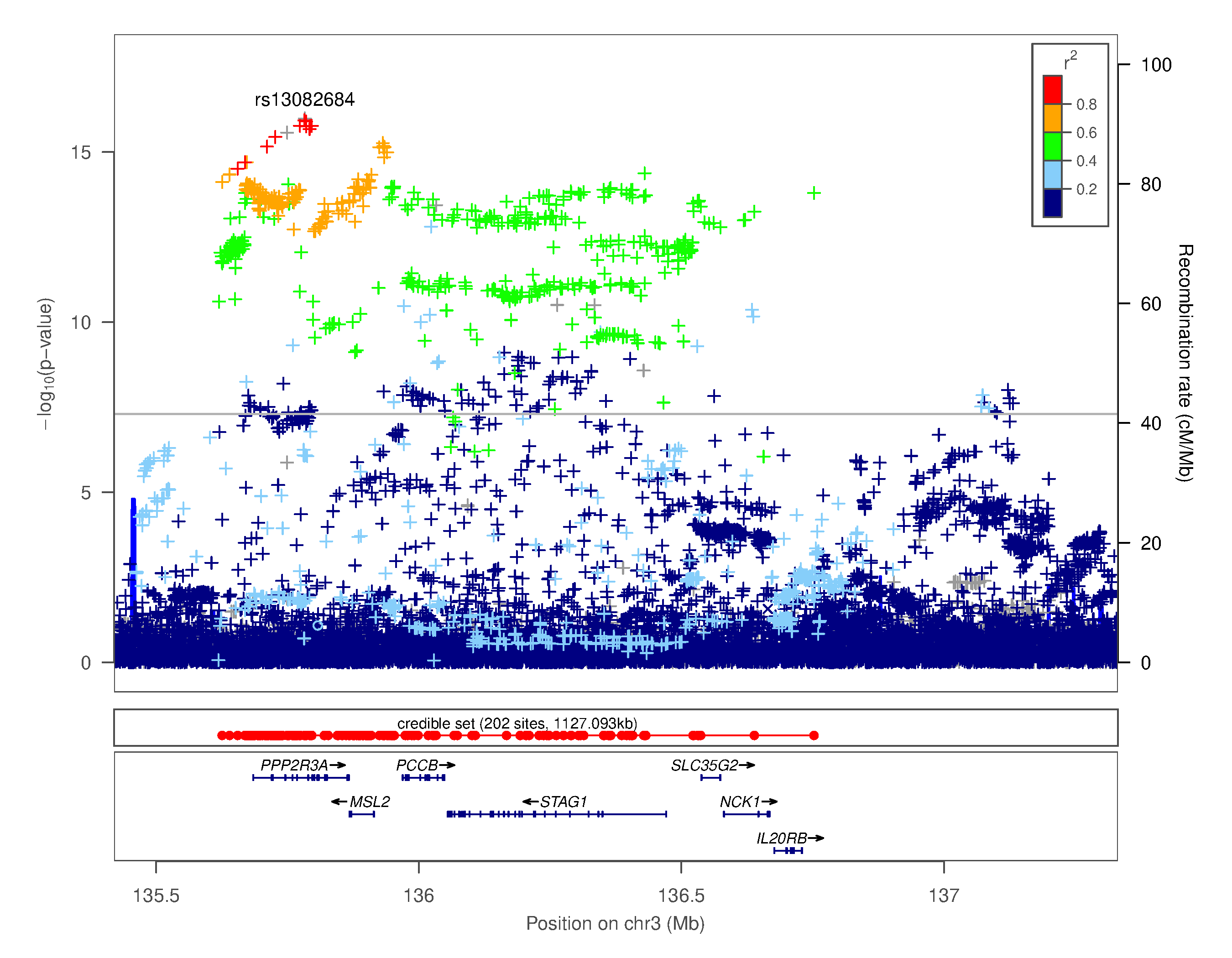

### Supplementary Figure 4.ii. Regional association plot for chr3q22.3 rs13082684

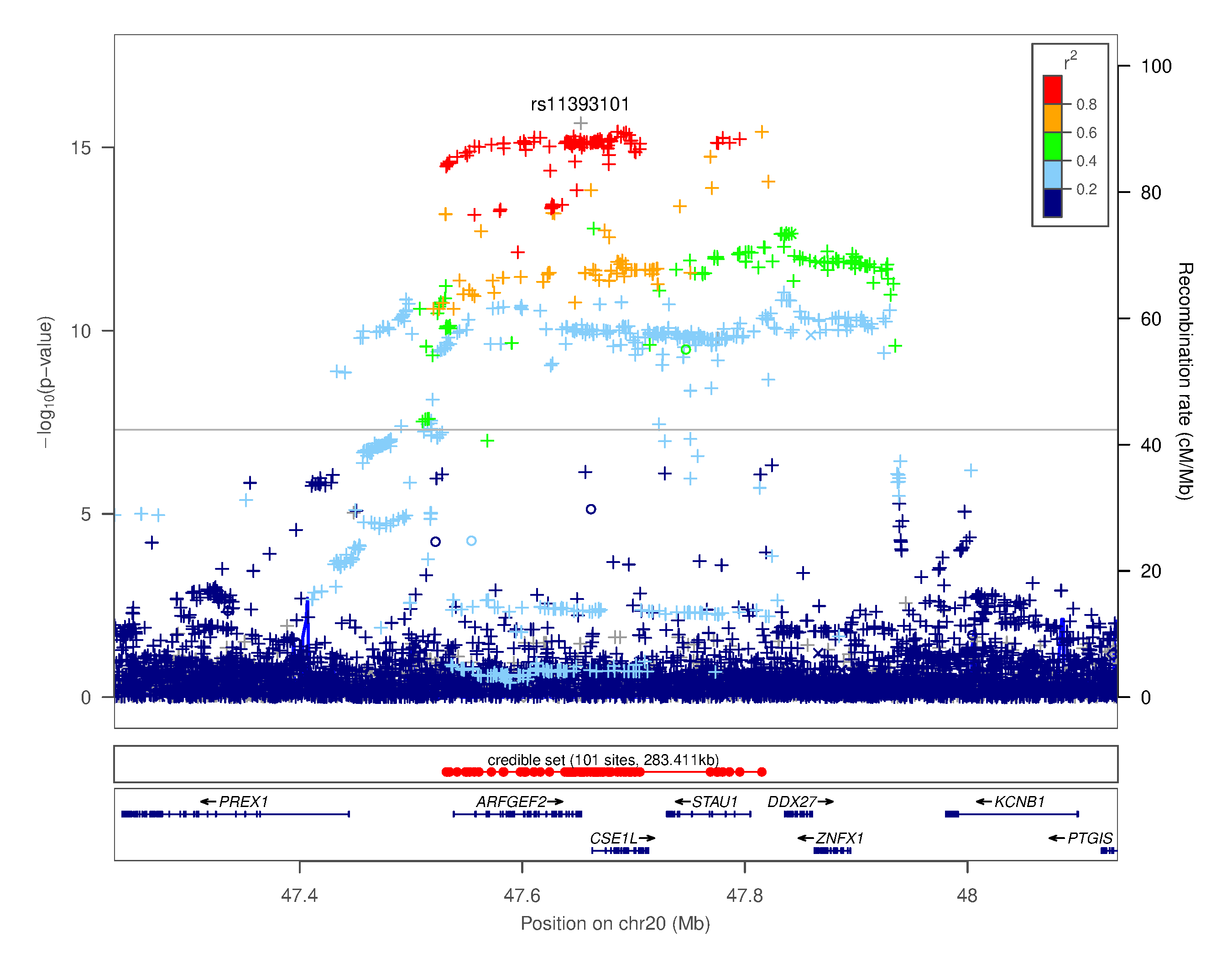

### Supplementary Figure 4.iii. Regional association plot for chr20q13.13 rs11393101

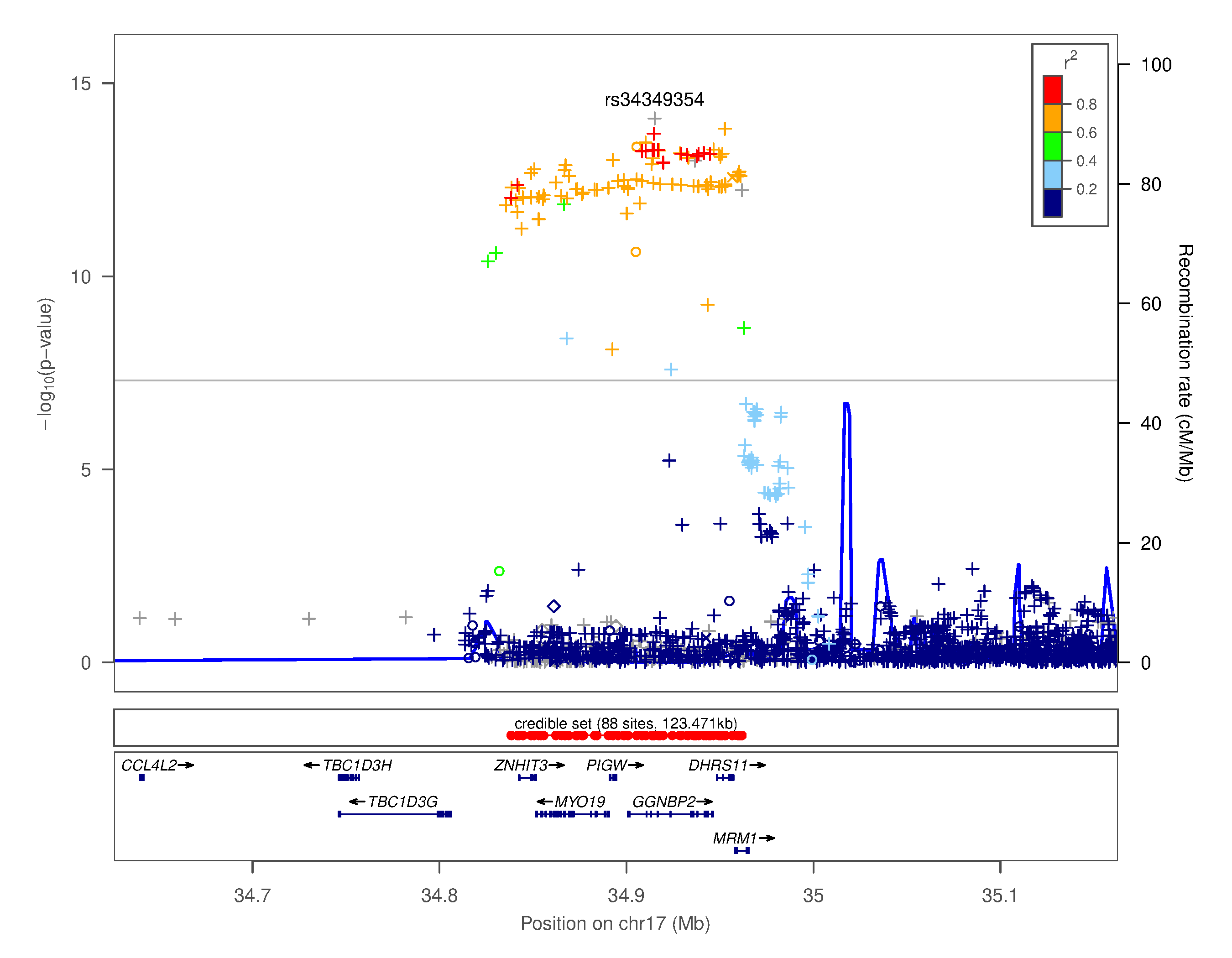

### Supplementary Figure 4.iv. Regional association plot for chr17q12 rs34349354

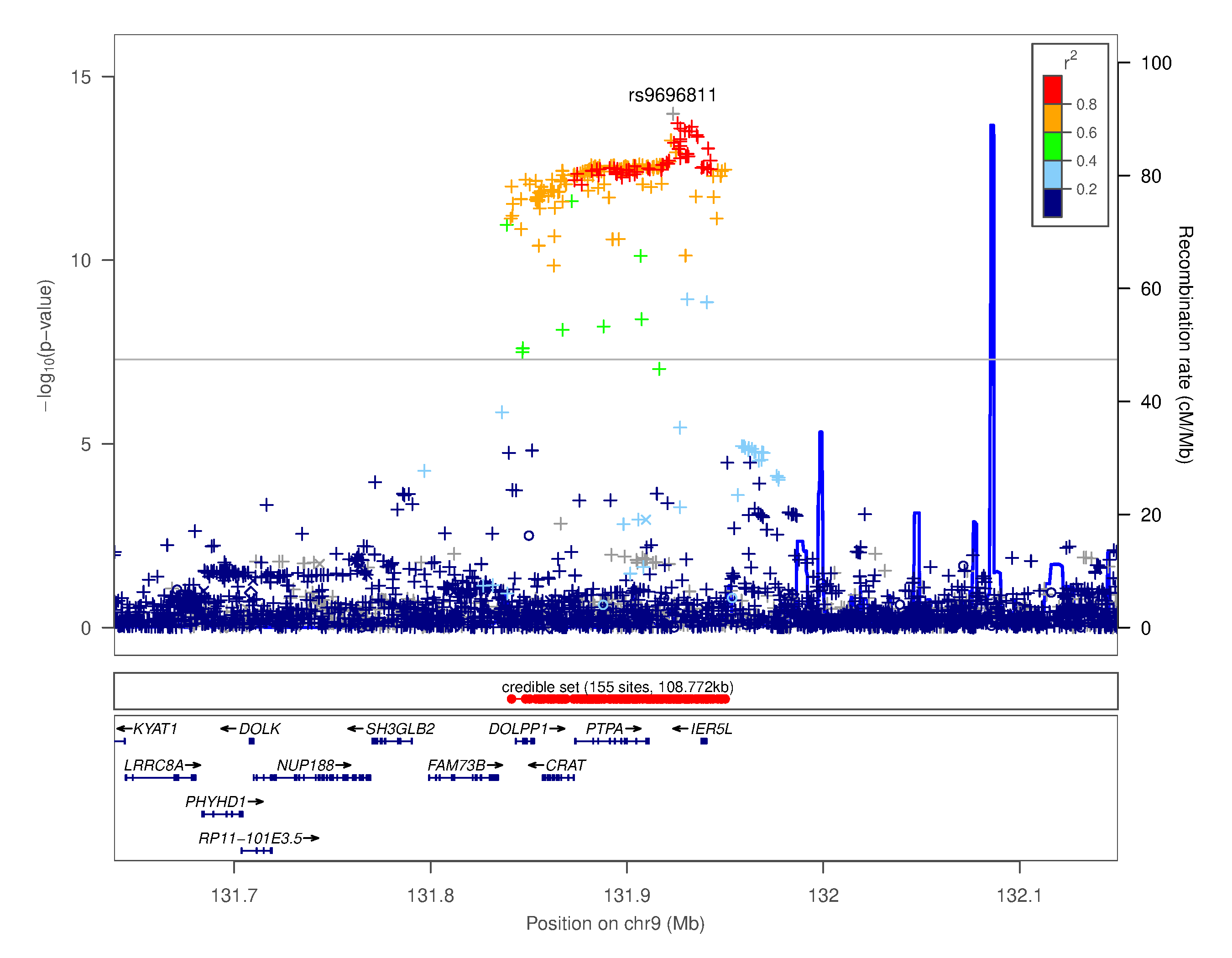

### Supplementary Figure 4.v. Regional association plot for chr9q34.11 rs9696811

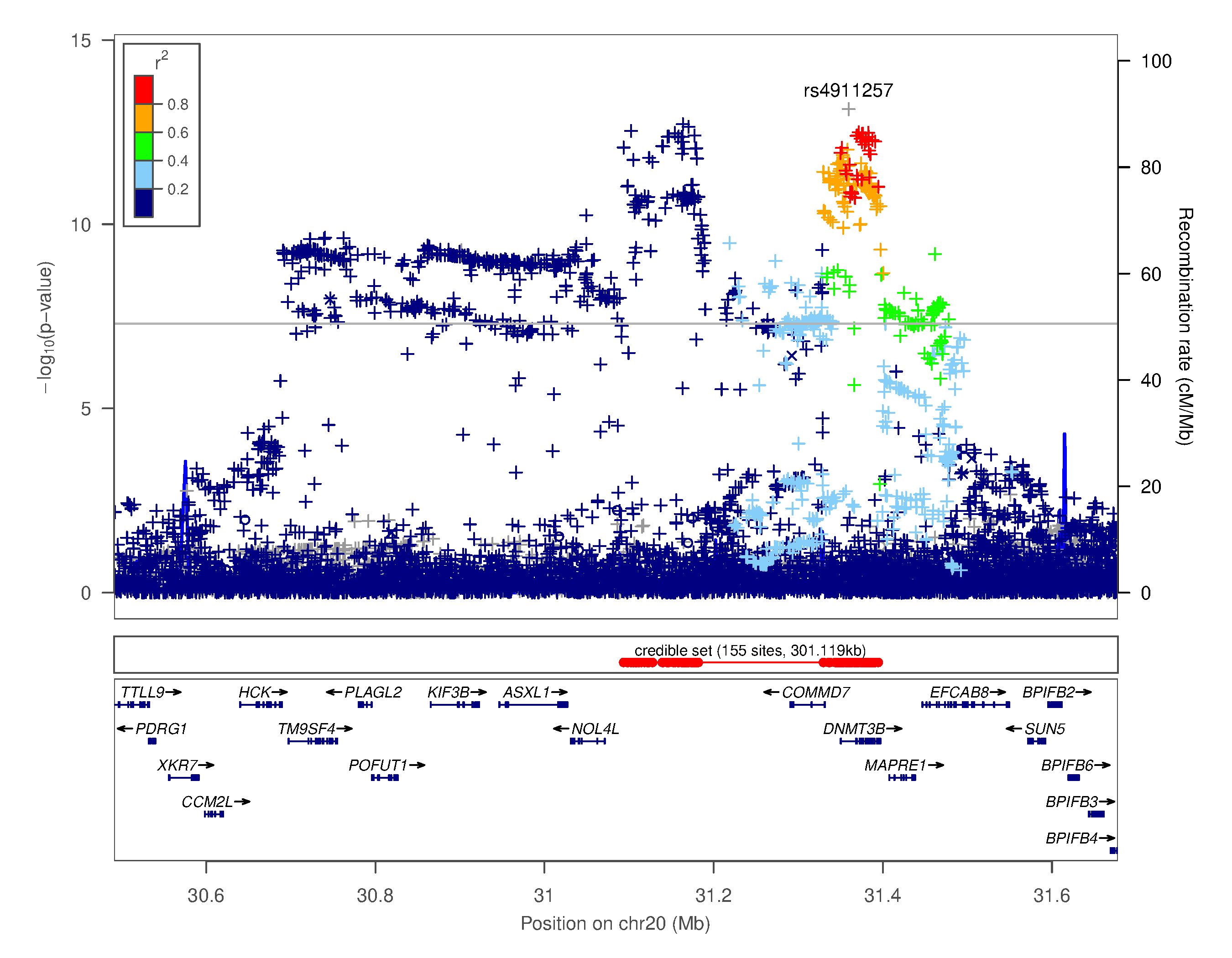

### Supplementary Figure 4.vi. Regional association plot for chr20q11.21 rs4911257

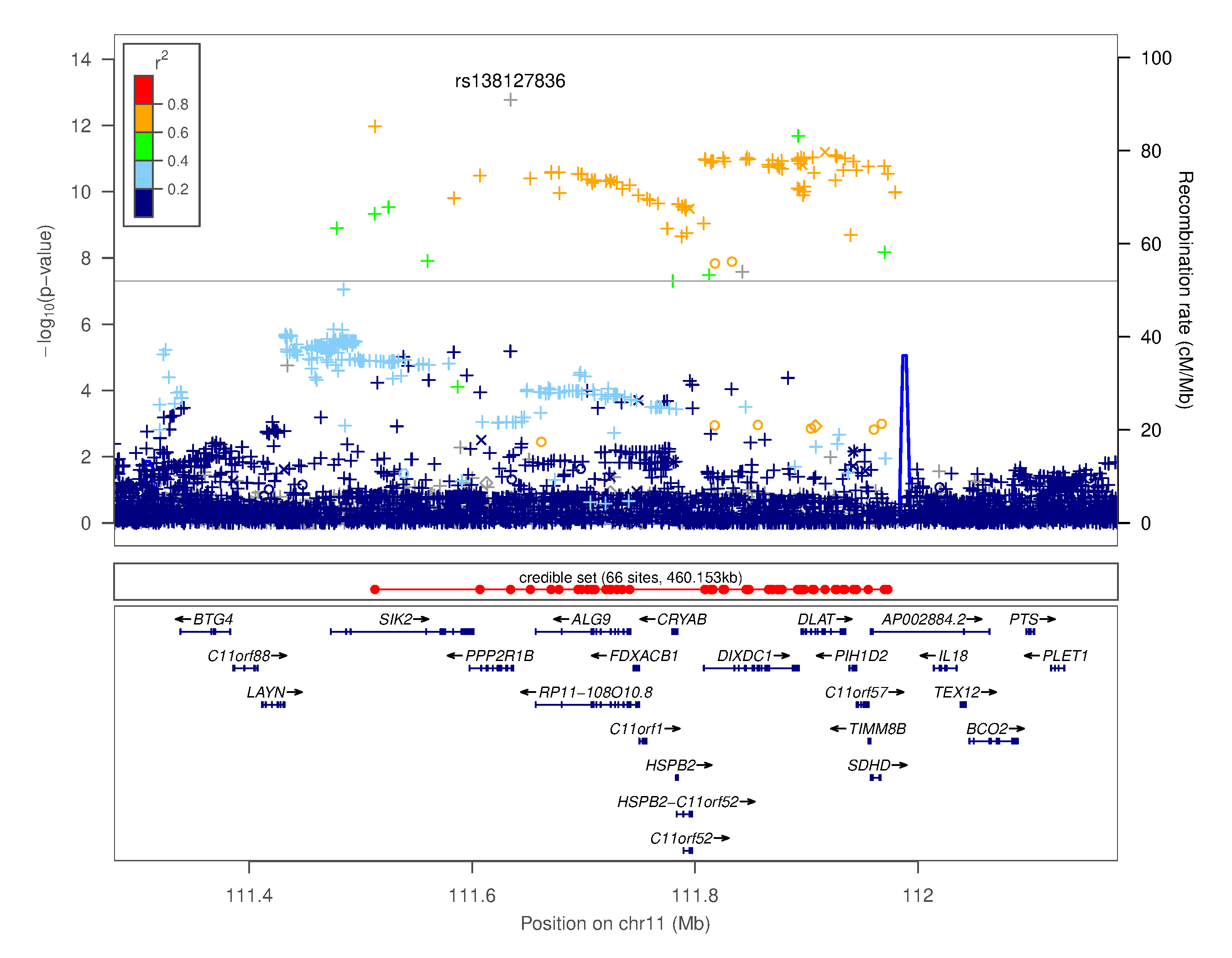

### Supplementary Figure 4.vii. Regional association plot for chr11q23.1 rs138127836

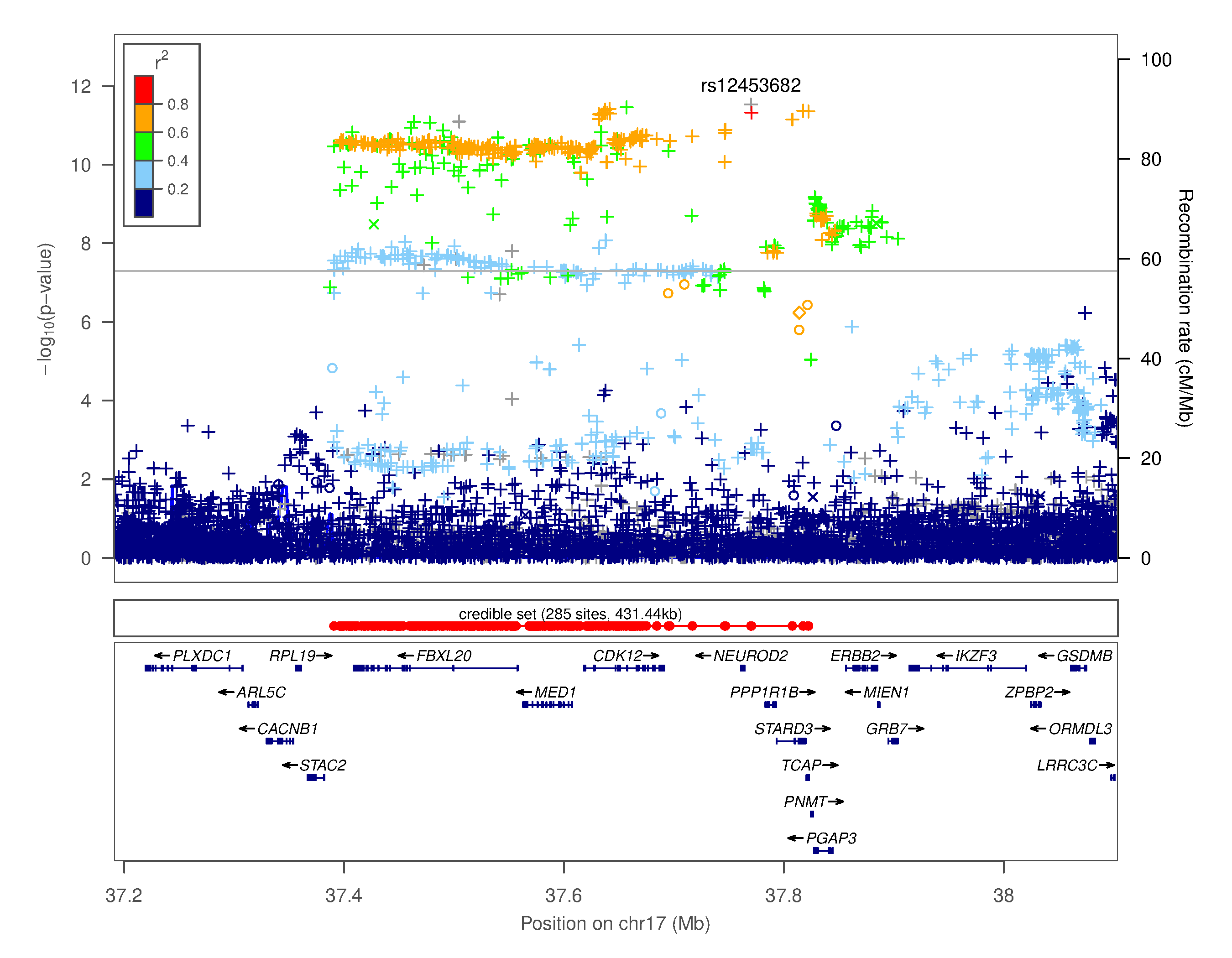

### Supplementary Figure 4.viii. Regional association plot for chr17q12 rs12453682

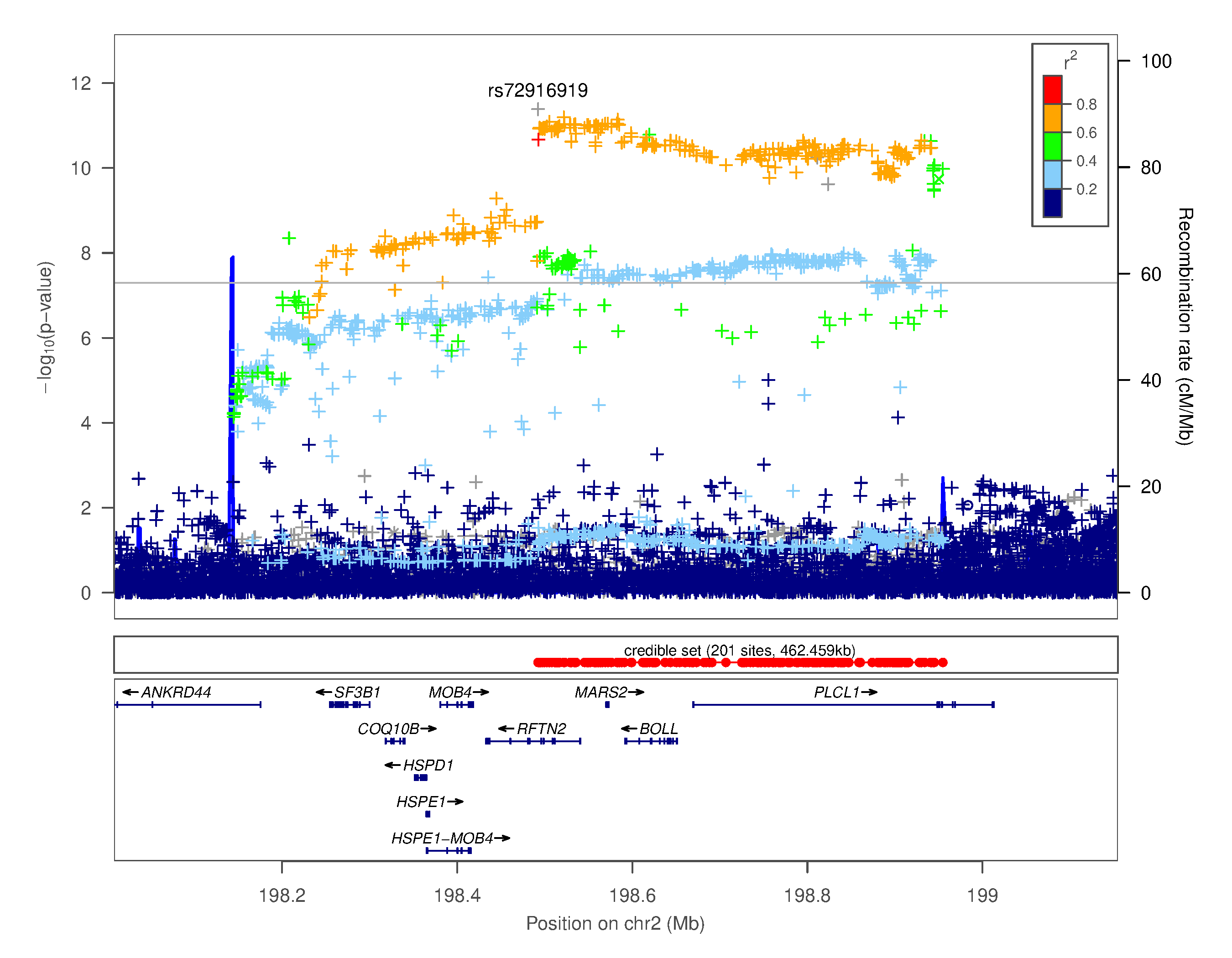

### Supplementary Figure 4.ix. Regional association plot for chr2q33.1 rs72916919

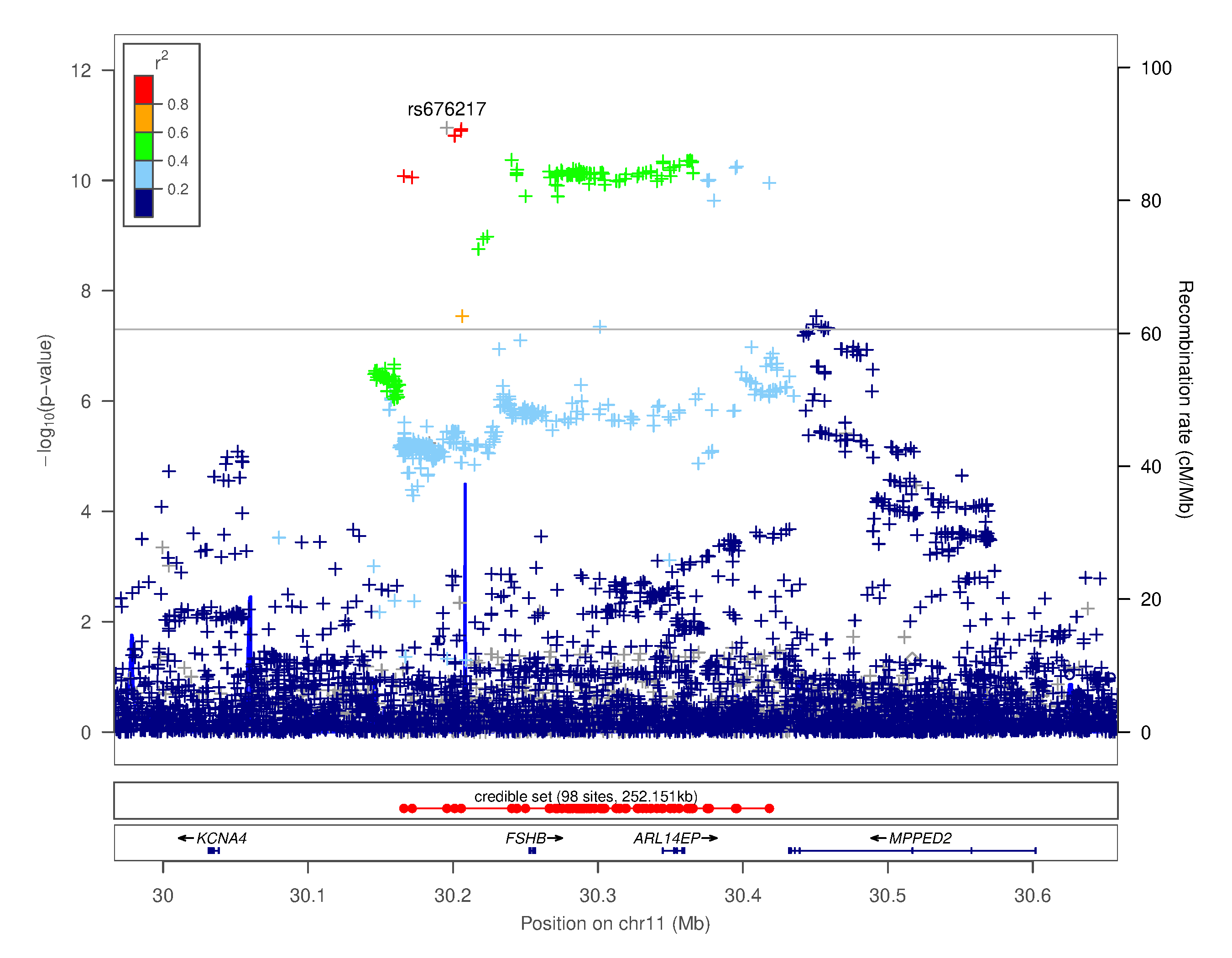

### Supplementary Figure 4.x. Regional association plot for chr11p14.1 rs676217

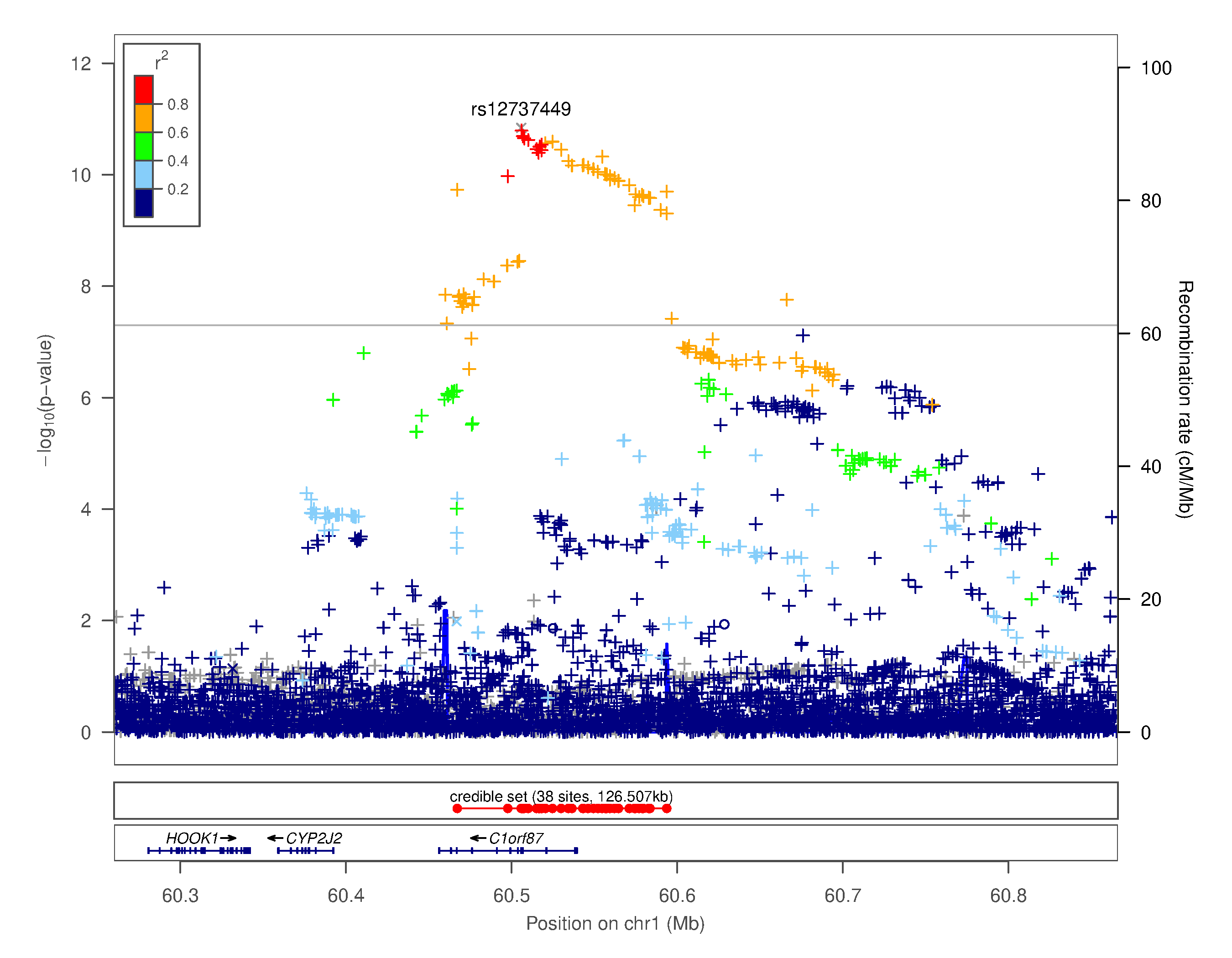

### Supplementary Figure 4.xi. Regional association plot for chr1p32.1 rs12737449

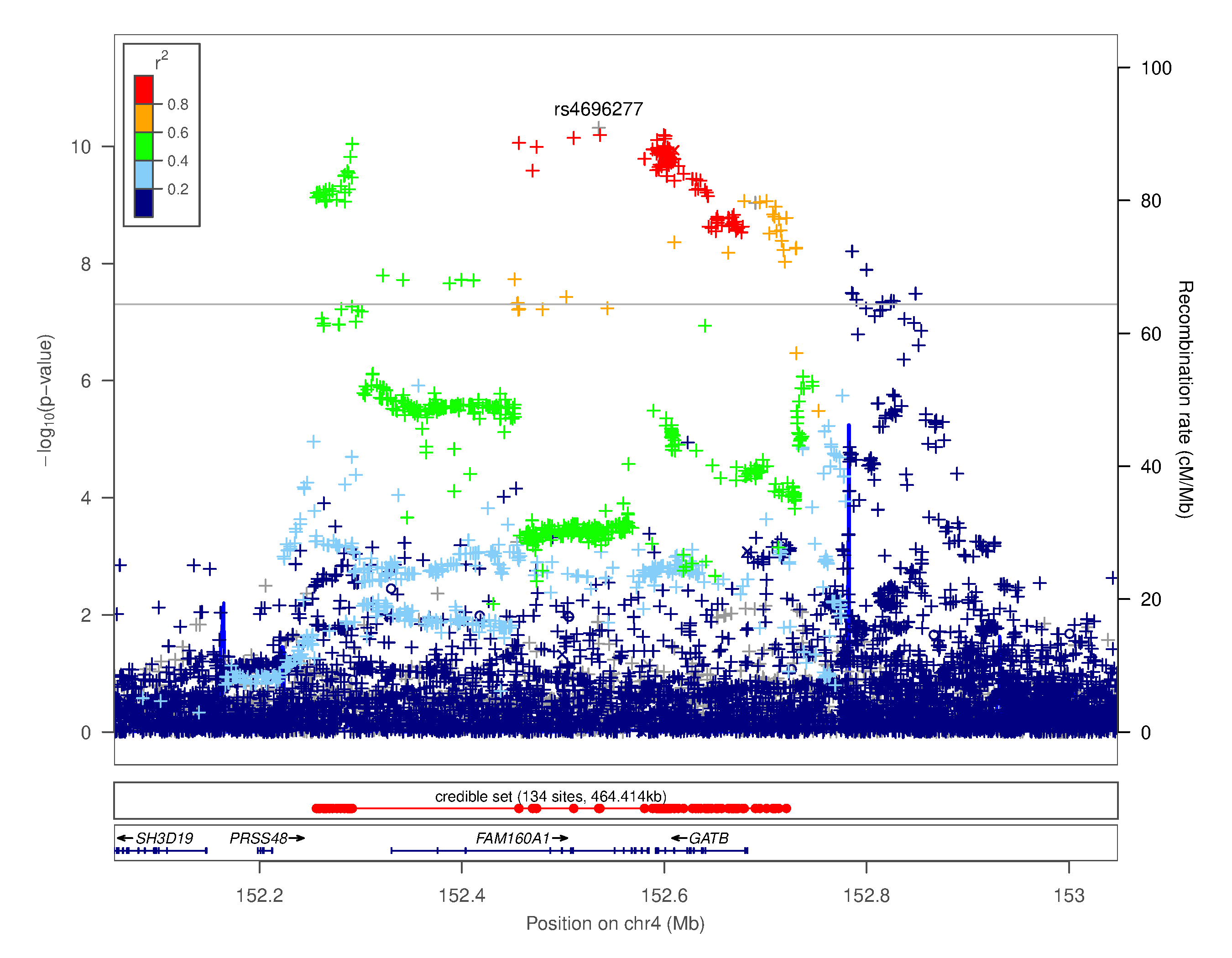

### Supplementary Figure 4.xii. Regional association plot for chr4q31.3 rs4696277

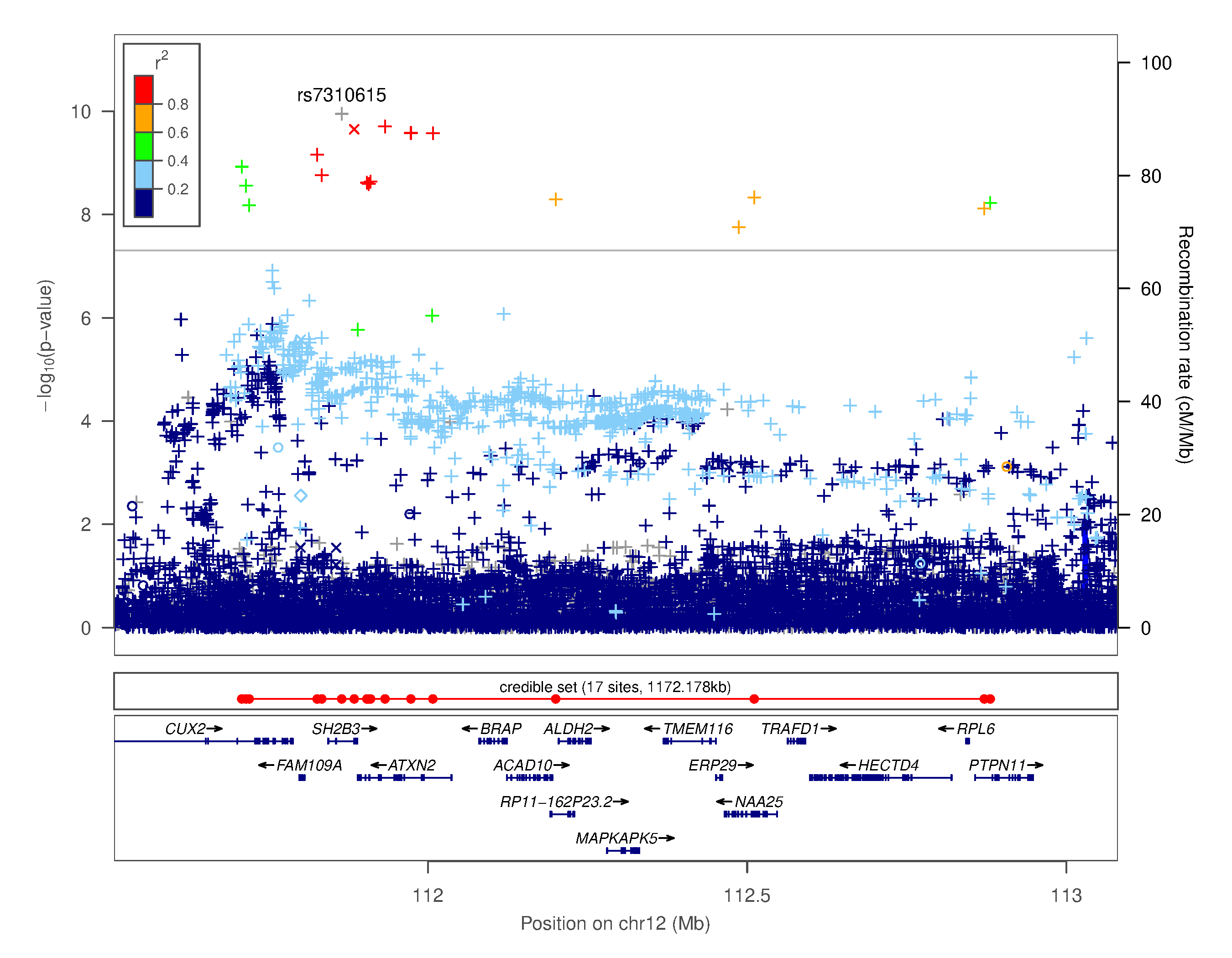

### Supplementary Figure 4.xiii. Regional association plot for chr12q24.12 rs7310615

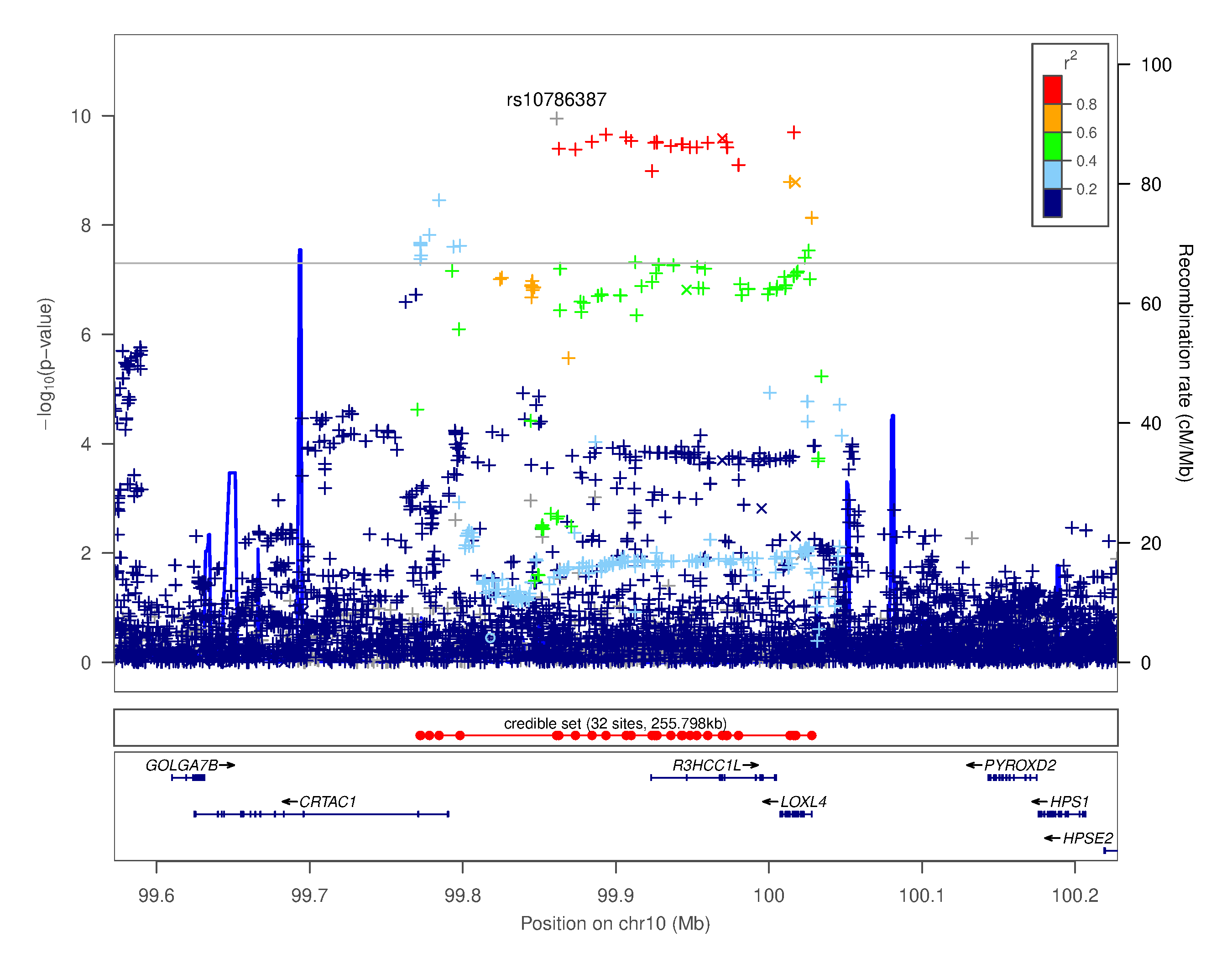

### Supplementary Figure 4.xiv. Regional association plot for chr10q24.2 rs10786387

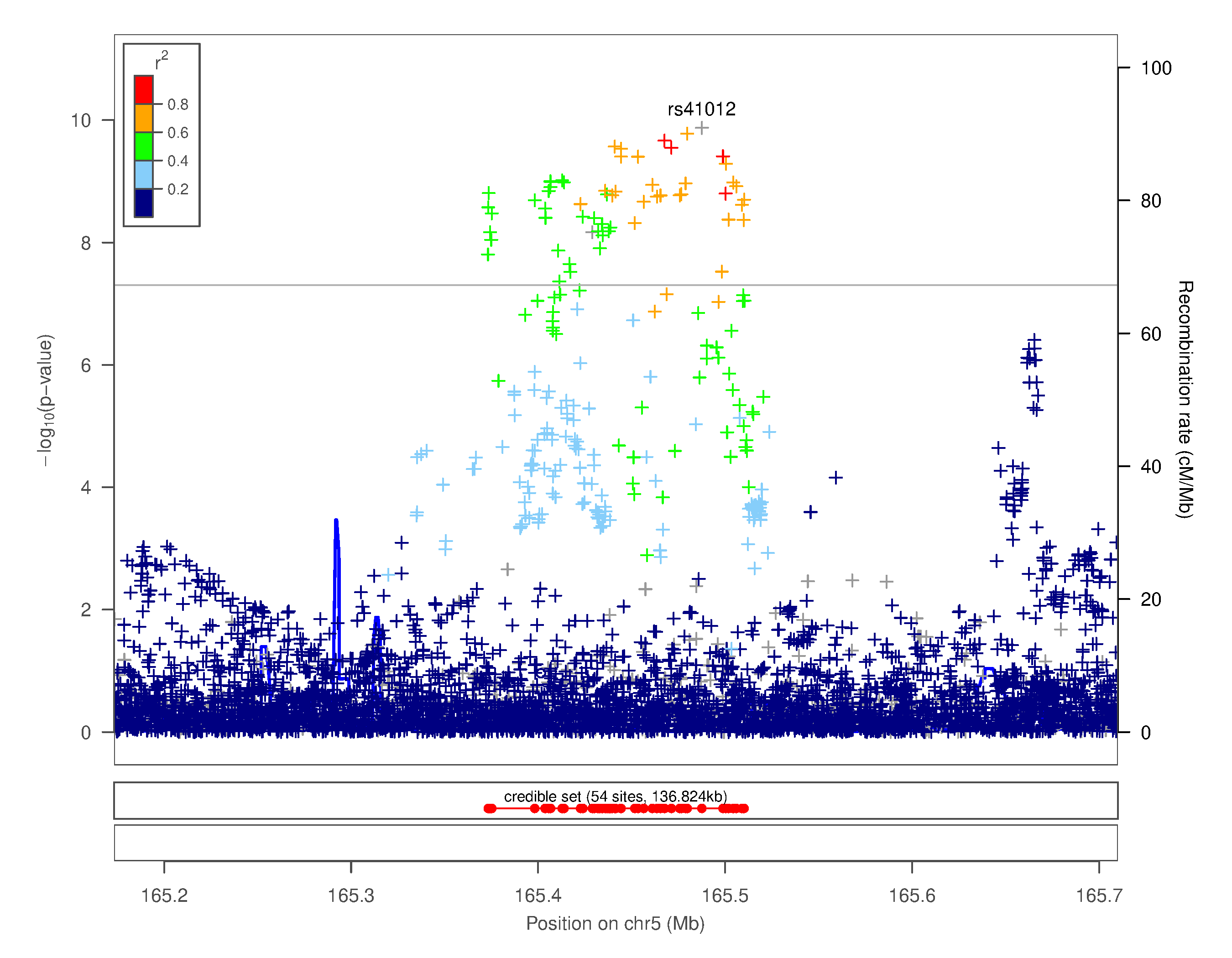

### Supplementary Figure 4.xv. Regional association plot for chr5q34 rs41012

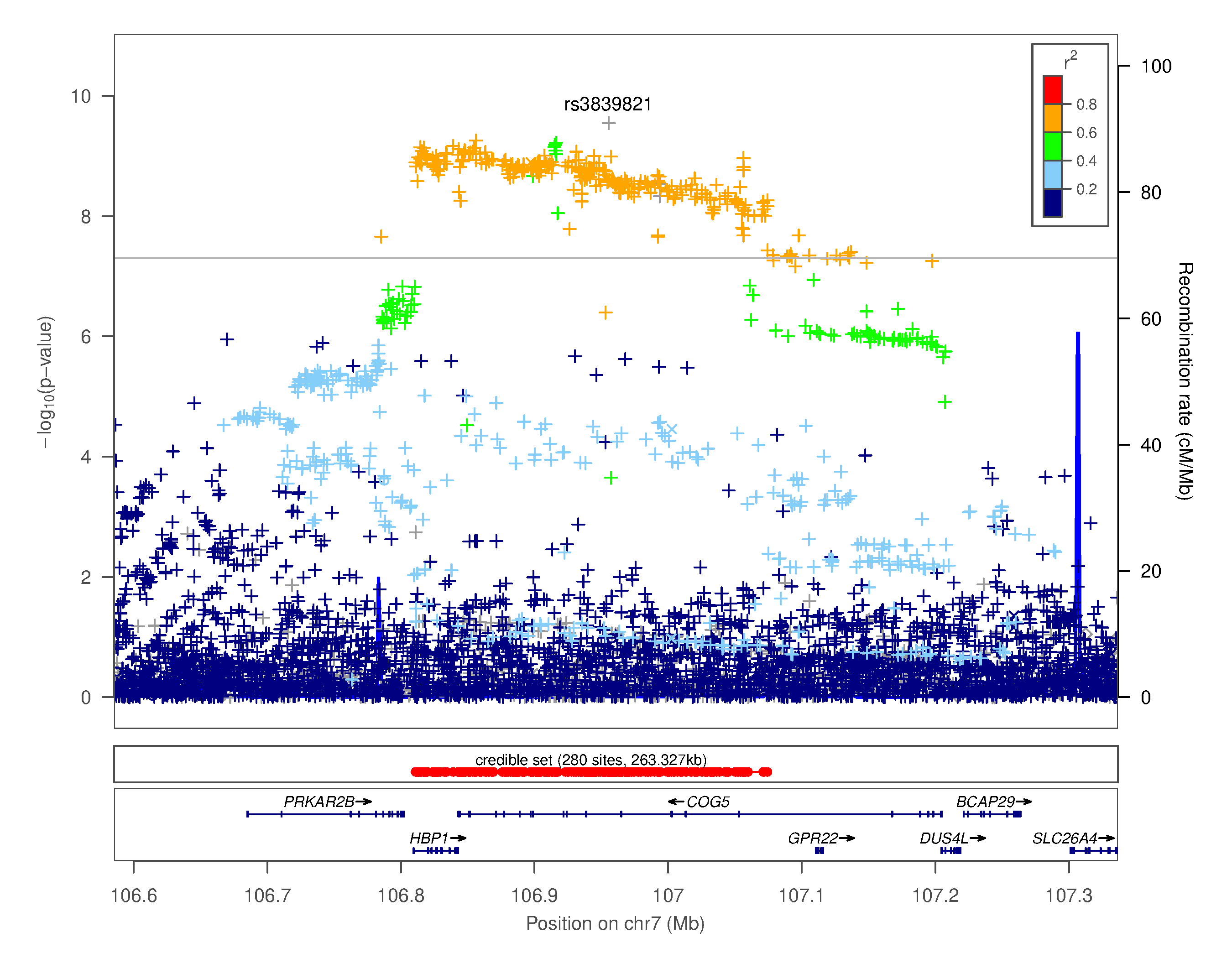

### Supplementary Figure 4.xvi. Regional association plot for chr7q22.3 rs3839821

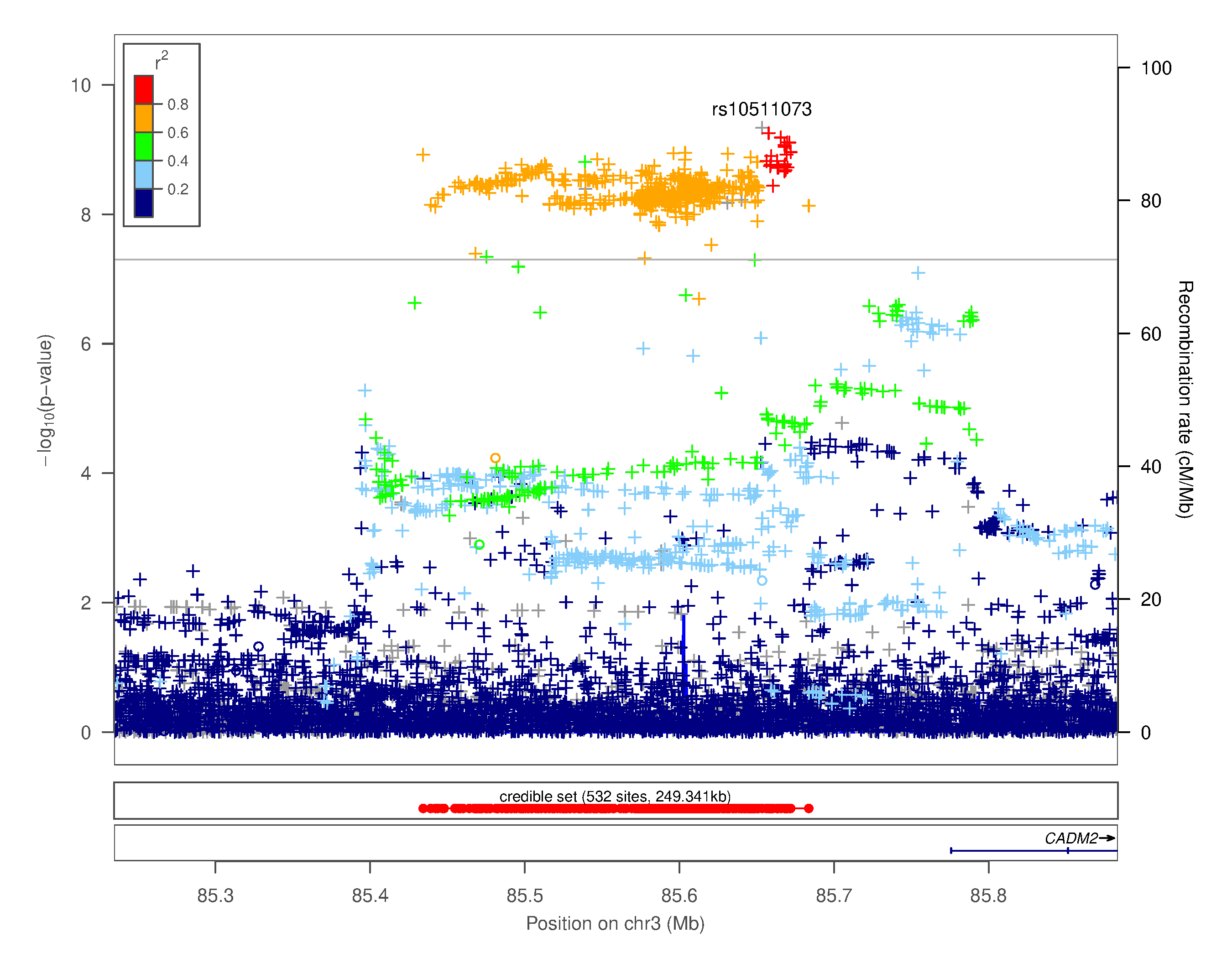

### Supplementary Figure 4.xvii. Regional association plot for chr3p12.1 rs10511073

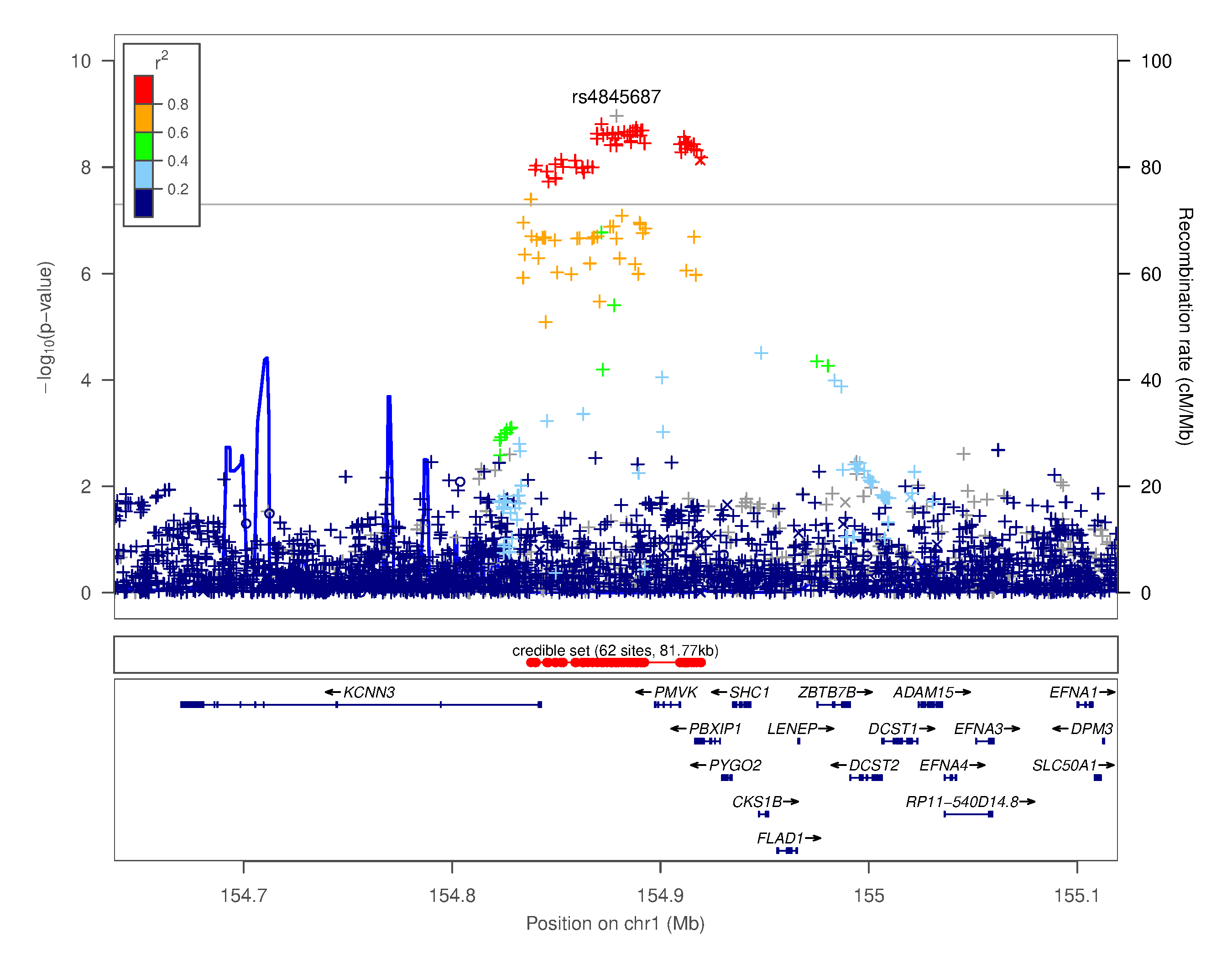

### Supplementary Figure 4.xviii. Regional association plot for chr1q21.3 rs4845687

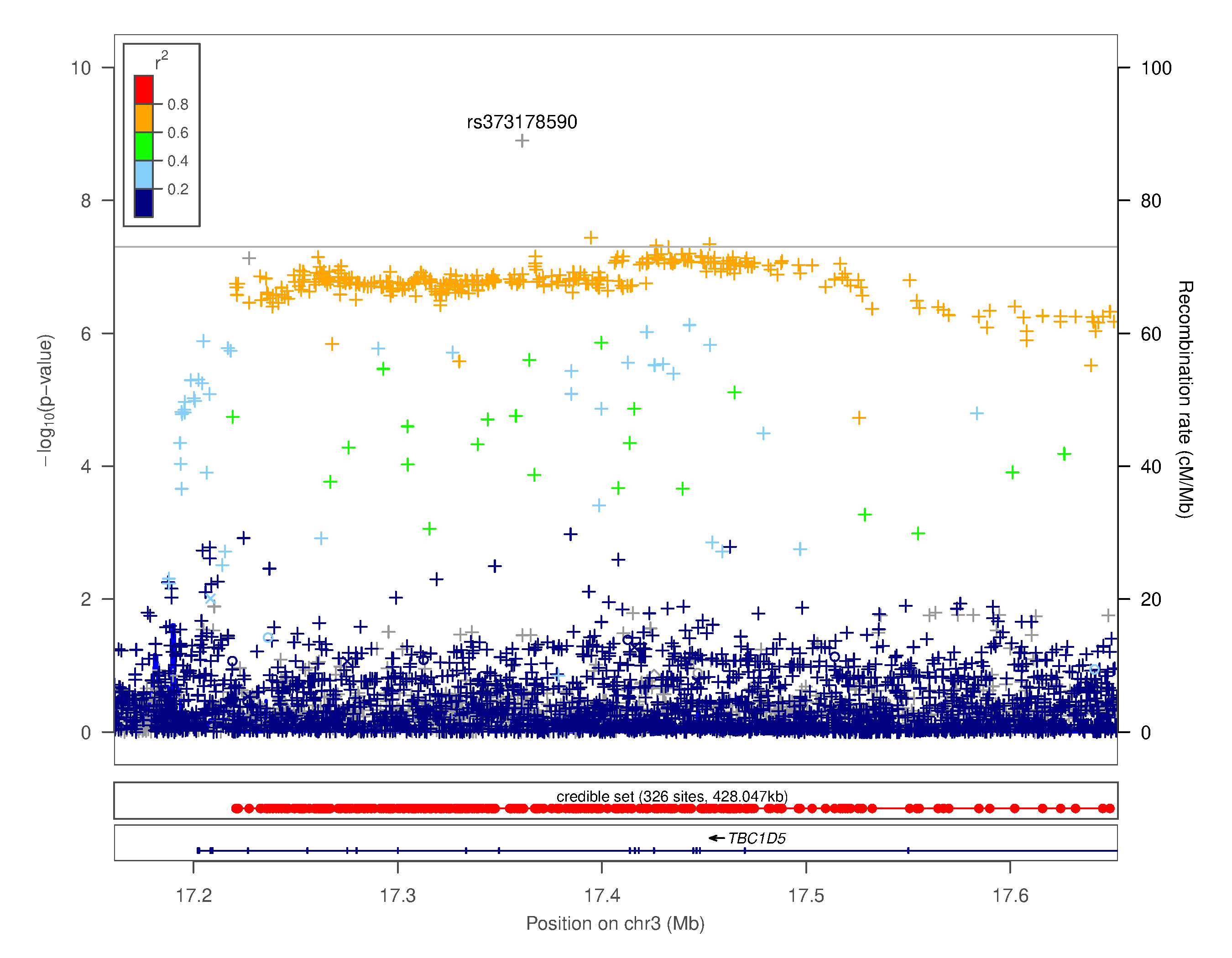

### Supplementary Figure 4.xix. Regional association plot for chr3p24.3 rs373178590

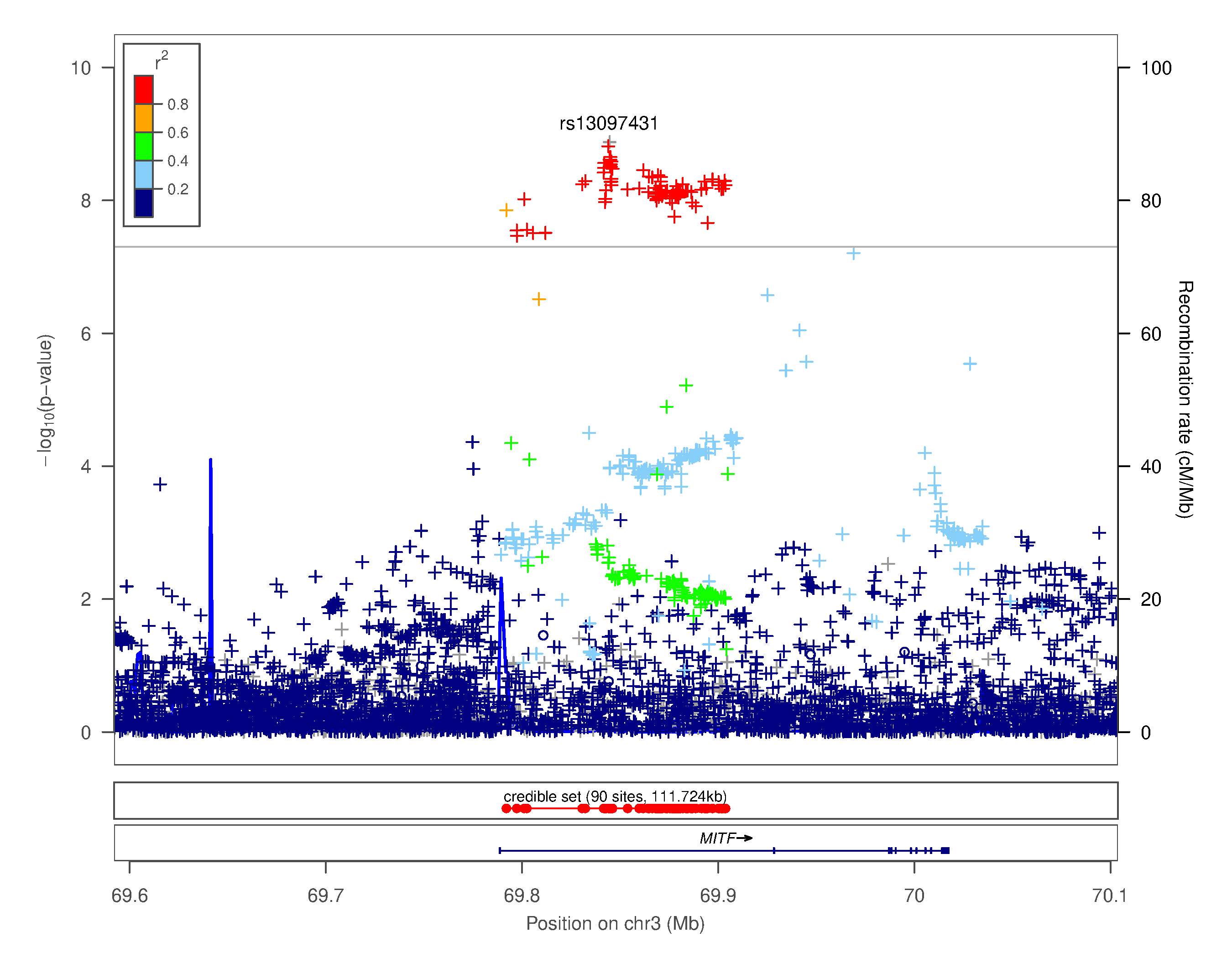

### Supplementary Figure 4.xx. Regional association plot for chr3p13 rs13097431

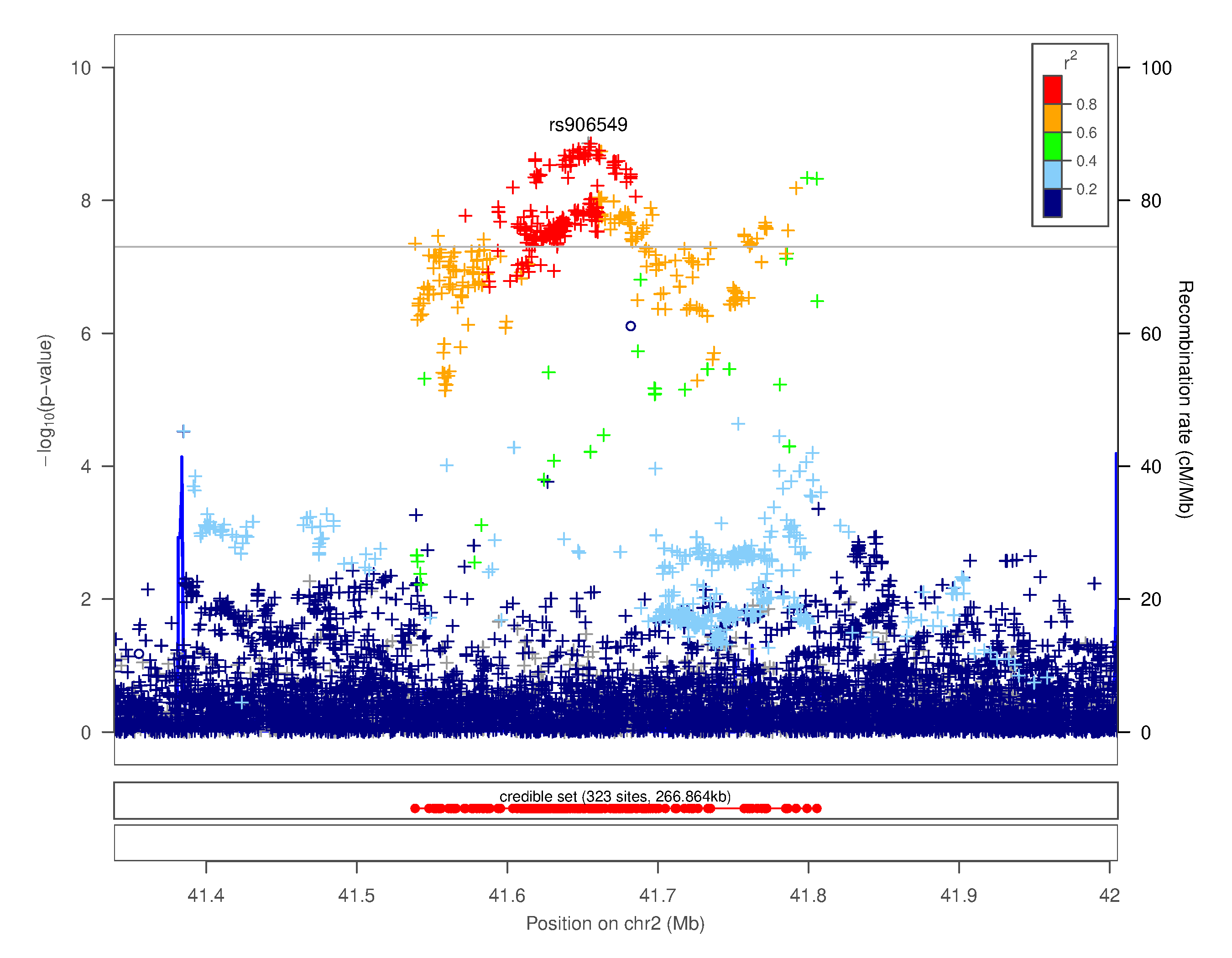

### Supplementary Figure 4.xxi. Regional association plot for chr2p22.1 rs906549

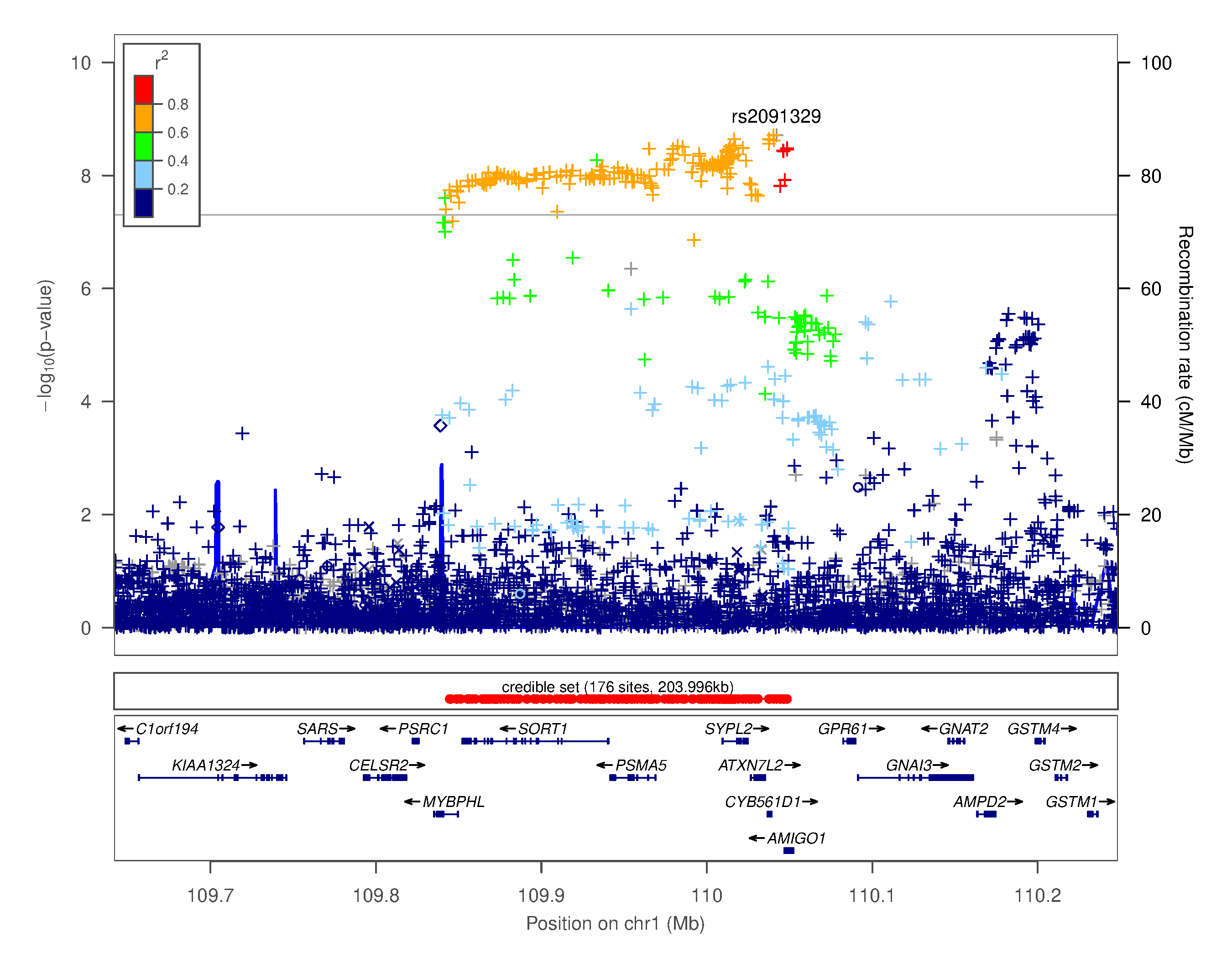

### Supplementary Figure 4.xxii. Regional association plot for chr1p13.3 rs2091329

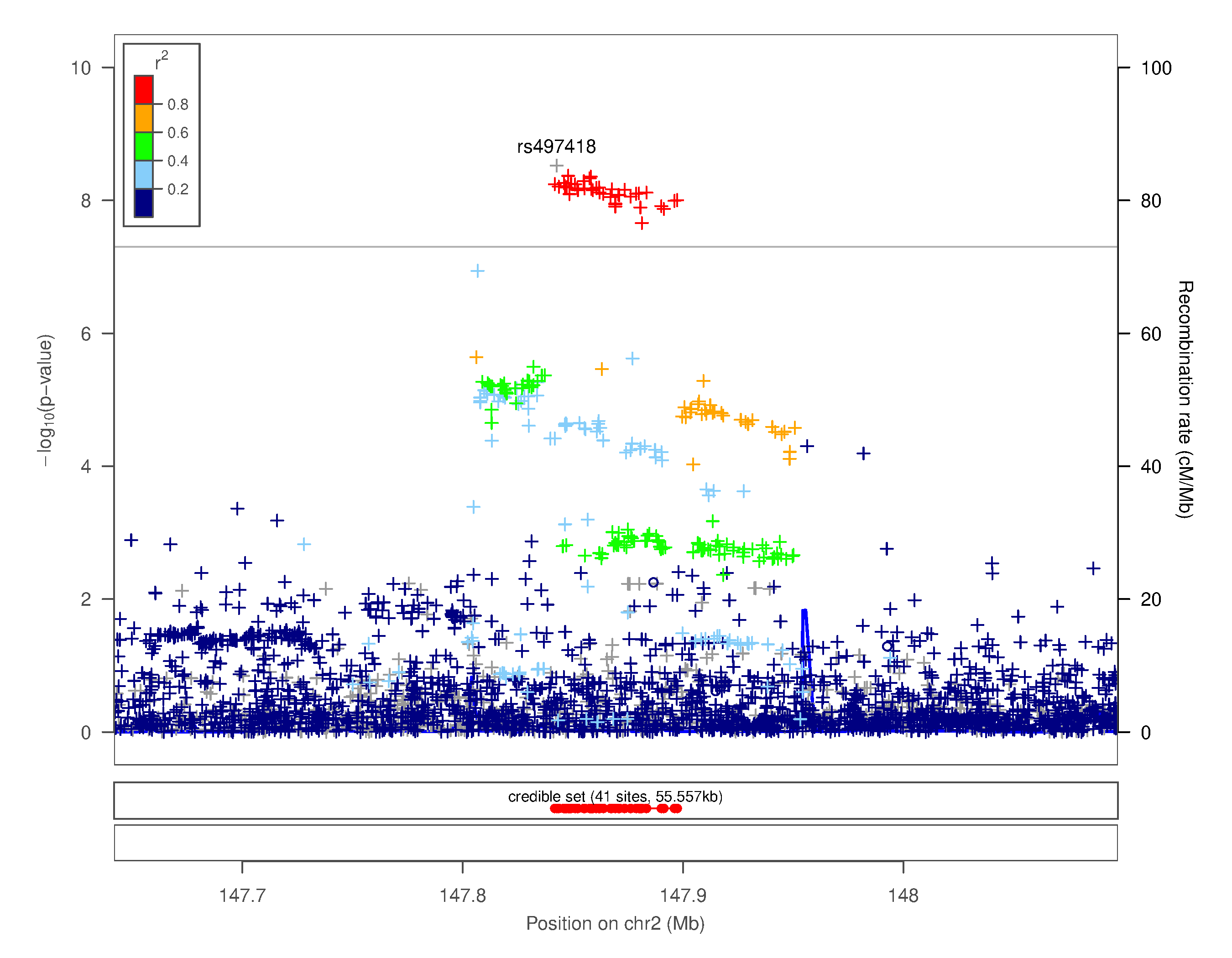

### Supplementary Figure 4.xxiii. Regional association plot for chr2q22.3 rs497418

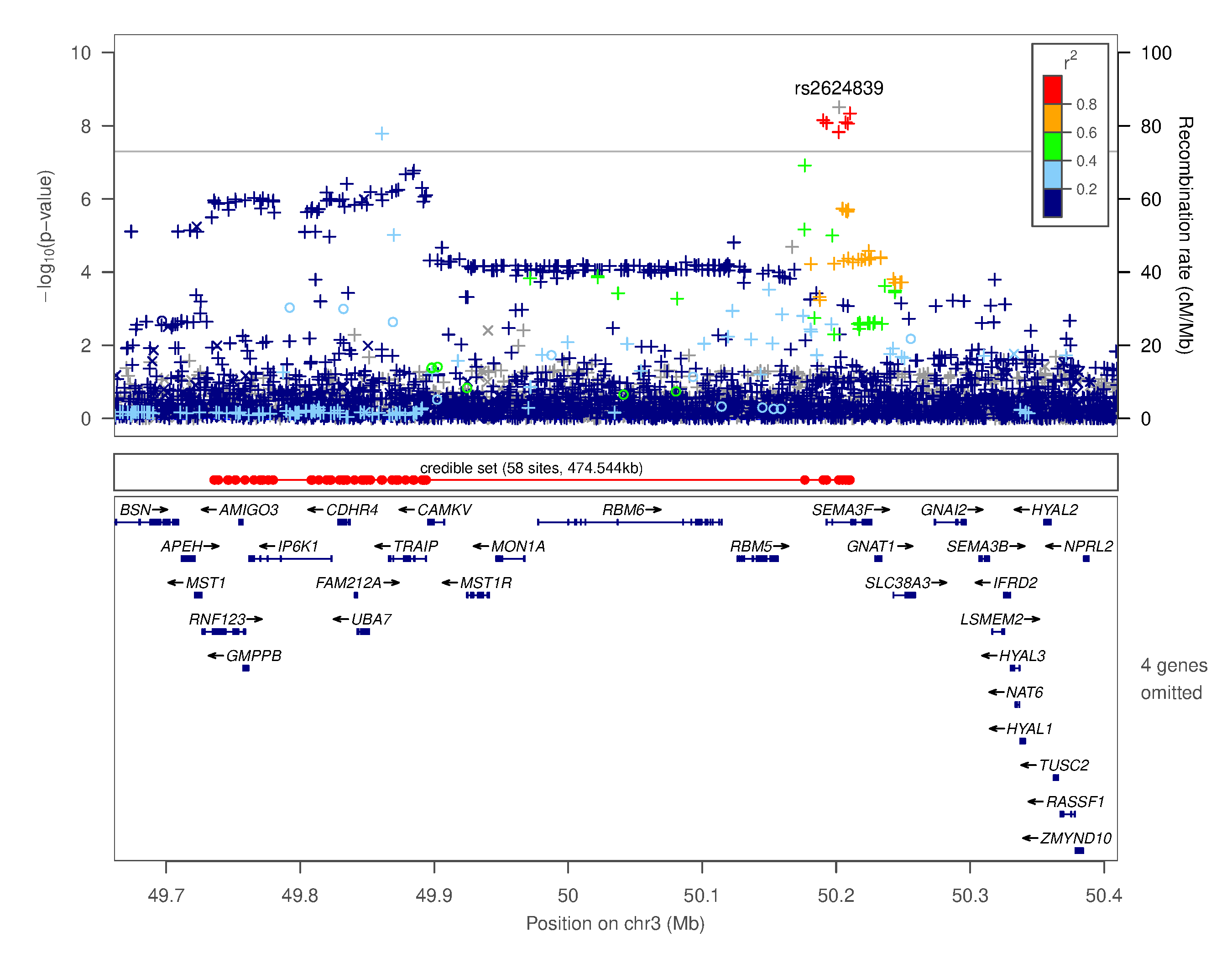

### Supplementary Figure 4.xxiv. Regional association plot for chr3p21.31 rs2624839

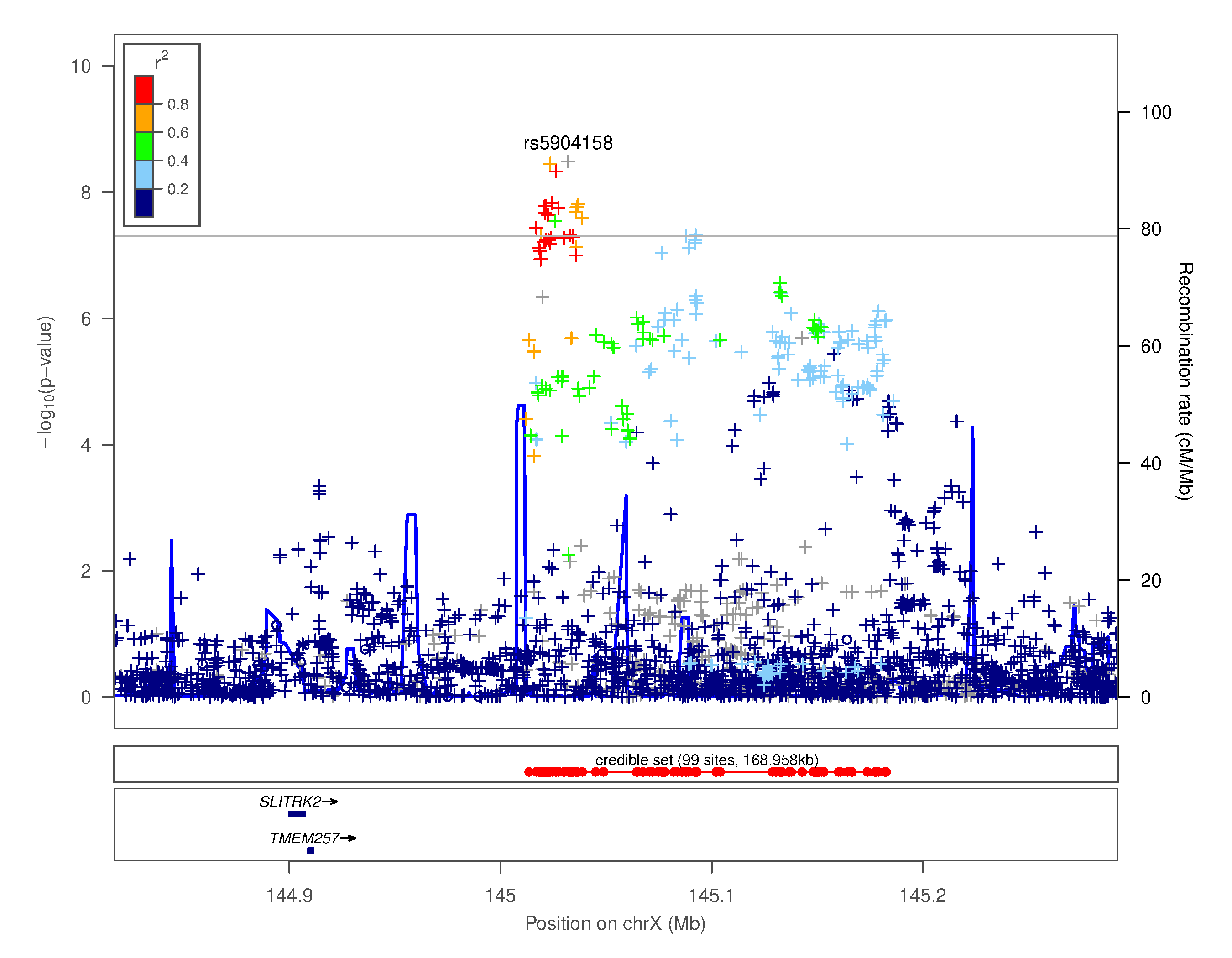

### Supplementary Figure 4.xxv. Regional association plot for chrXq27.3 rs5904158

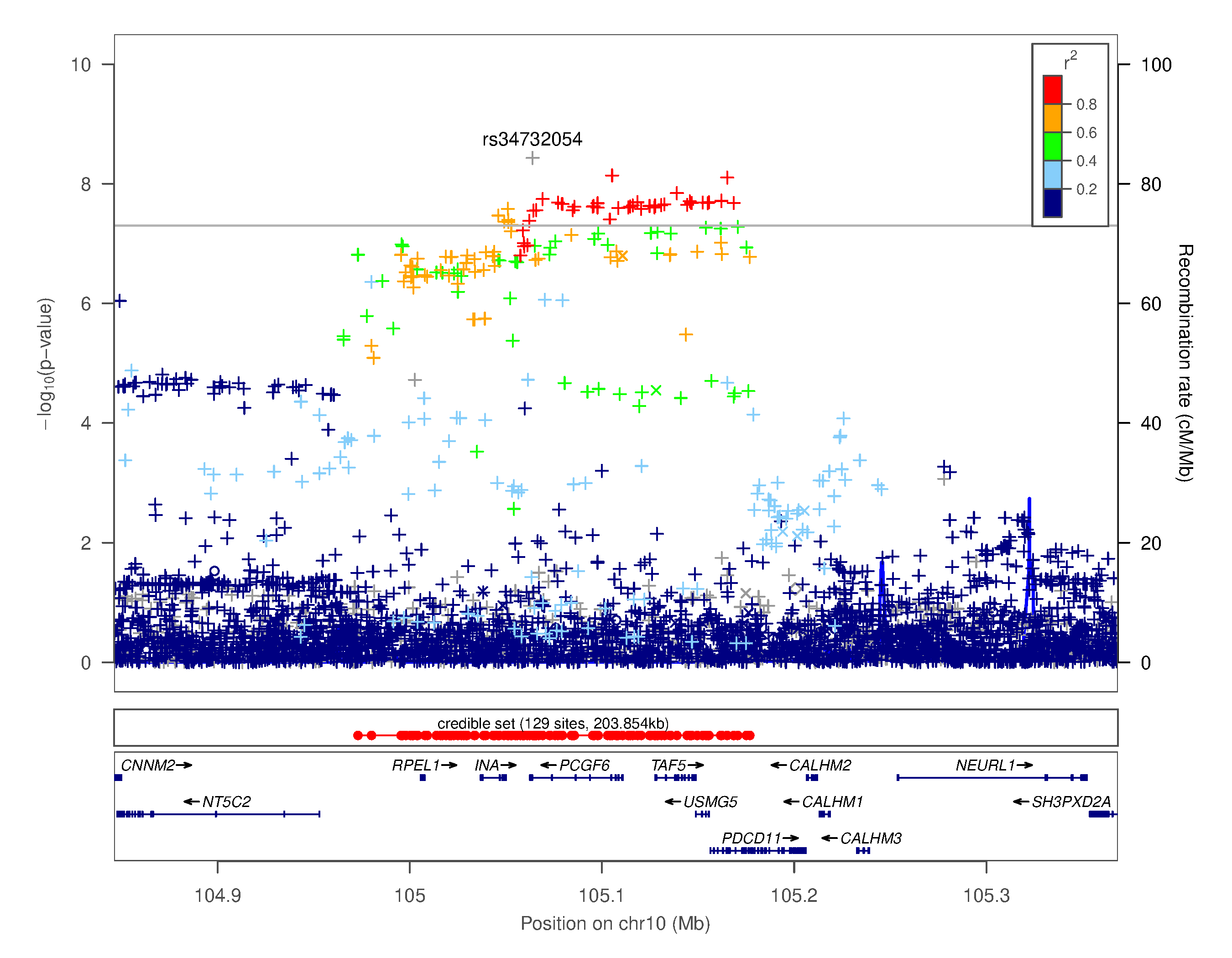

### Supplementary Figure 4.xxvi. Regional association plot for chr10q24.33 rs34732054

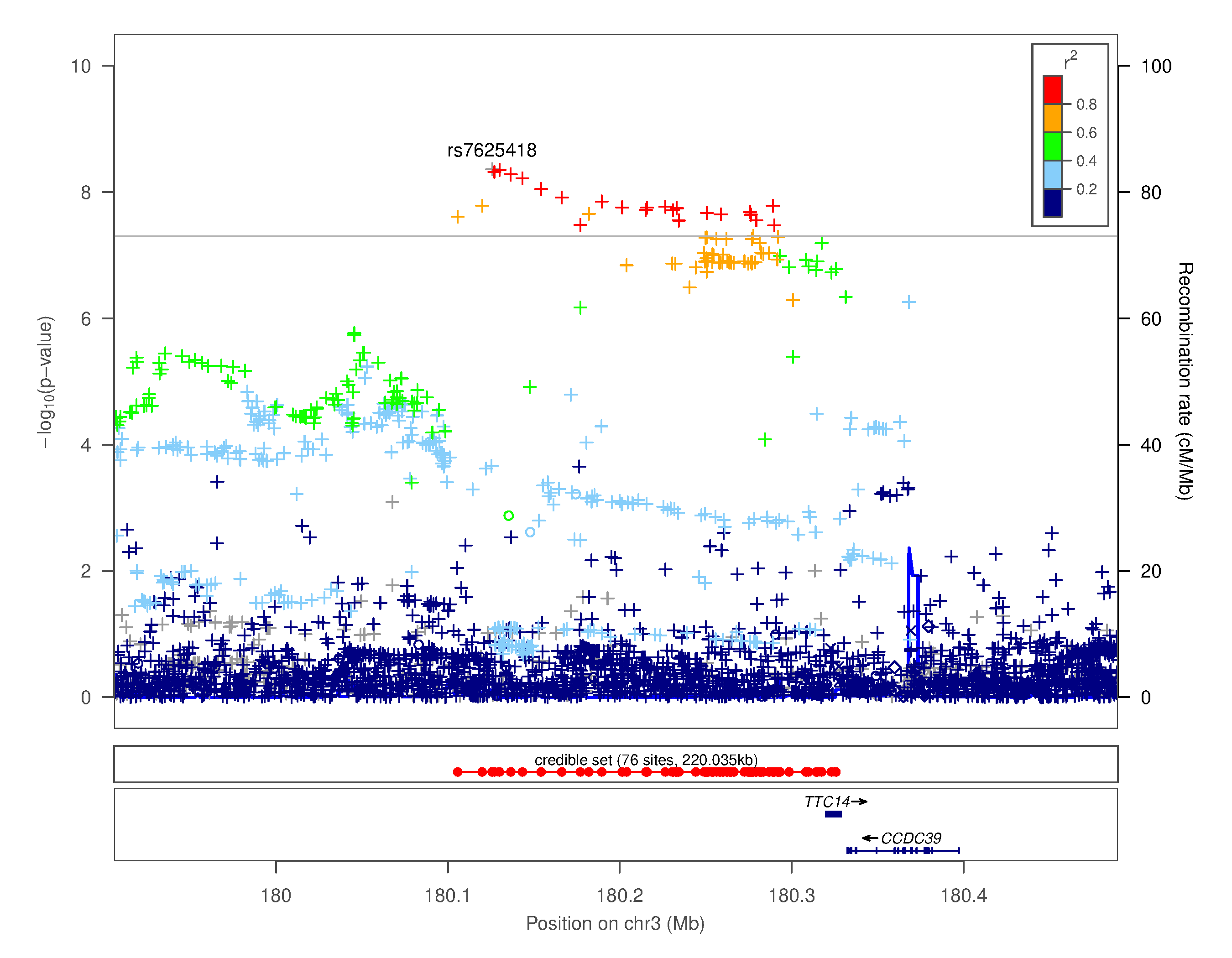

### Supplementary Figure 4.xxvii. Regional association plot for chr3q26.33 rs7625418

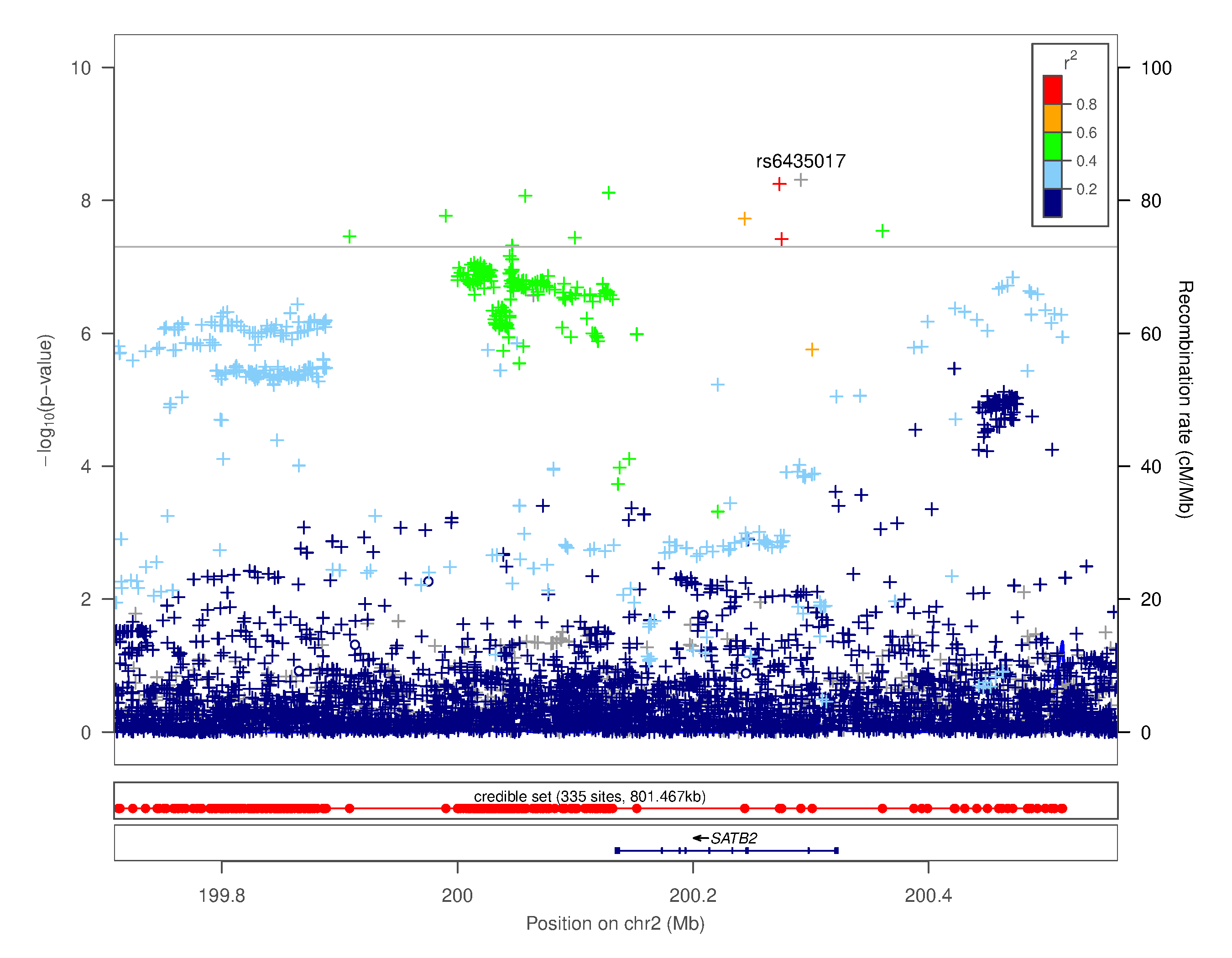

### Supplementary Figure 4.xxviii. Regional association plot for chr2q33.1 rs6435017

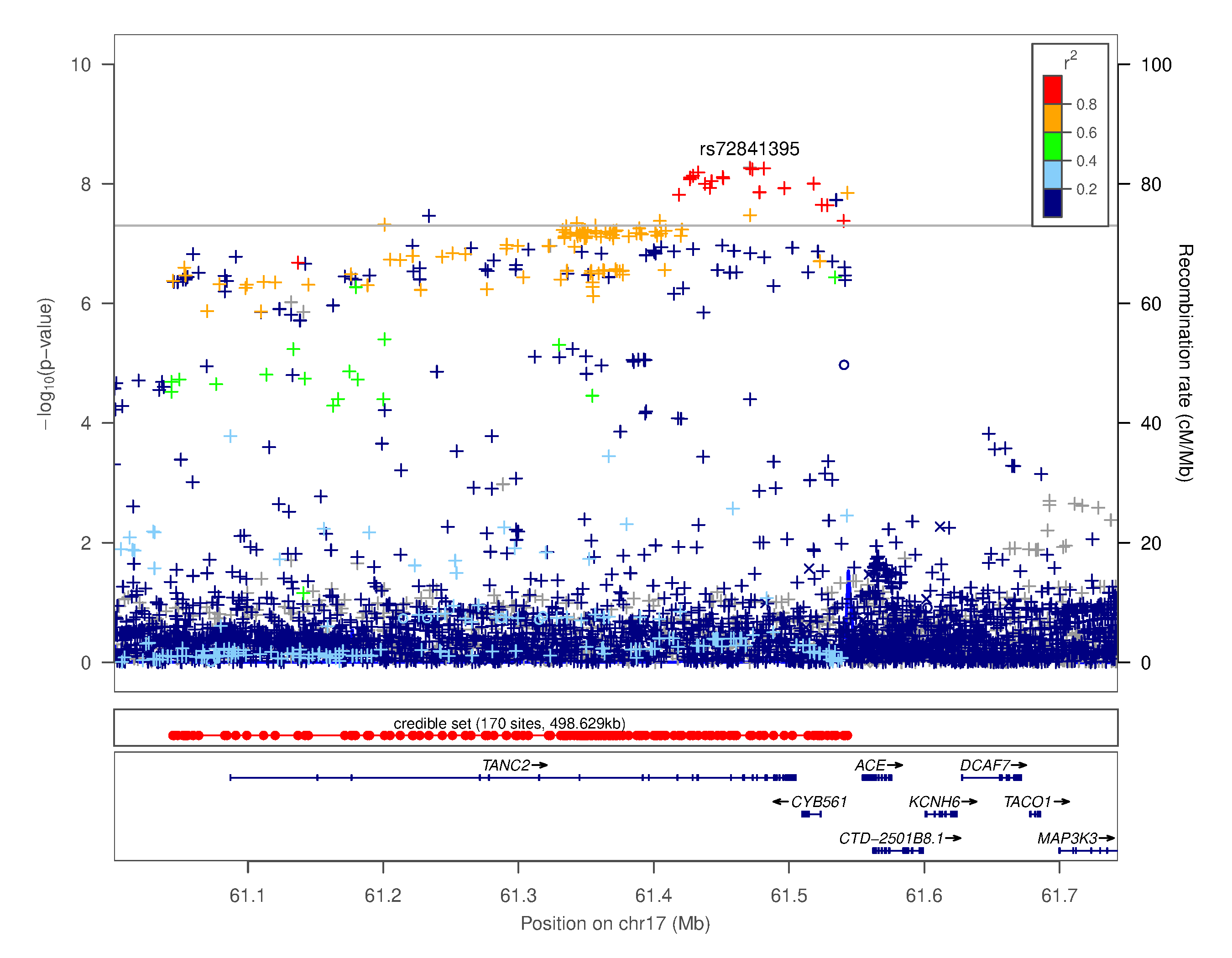

### Supplementary Figure 4.xxix. Regional association plot for chr17q23.3 rs72841395

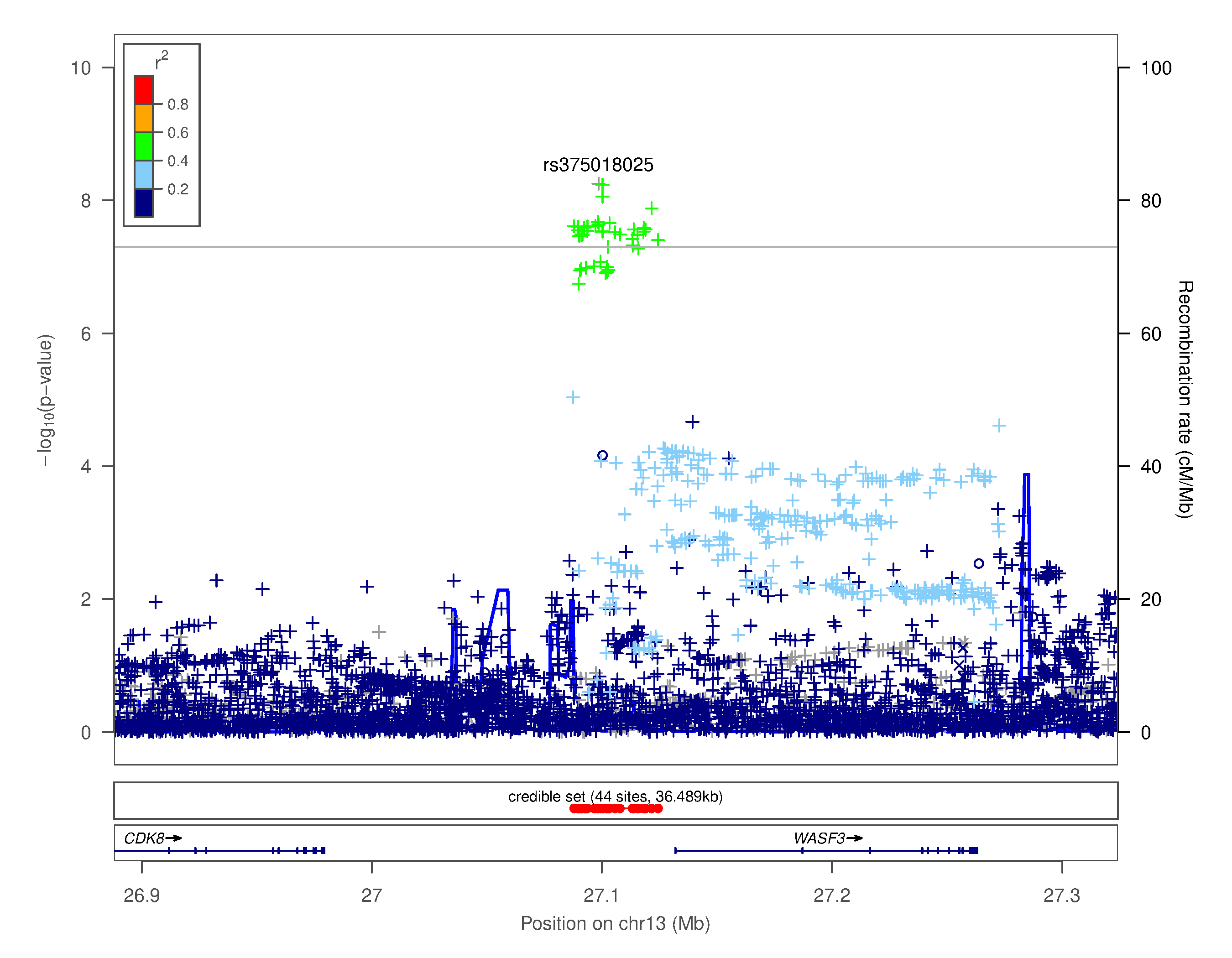

### Supplementary Figure 4.xxx. Regional association plot for chr13q12.13 rs375018025

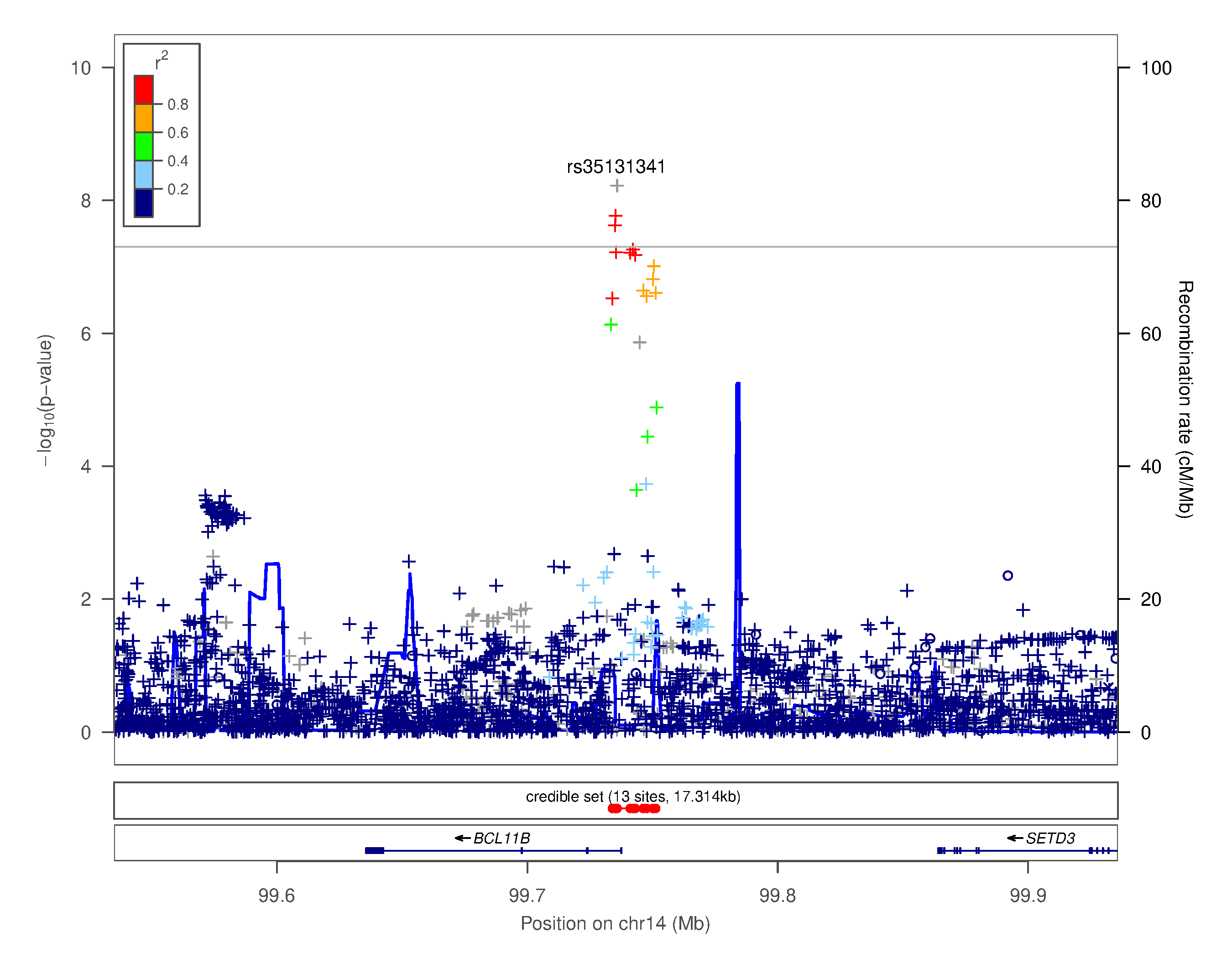

### Supplementary Figure 4.xxxi. Regional association plot for chr14q32.2 rs35131341

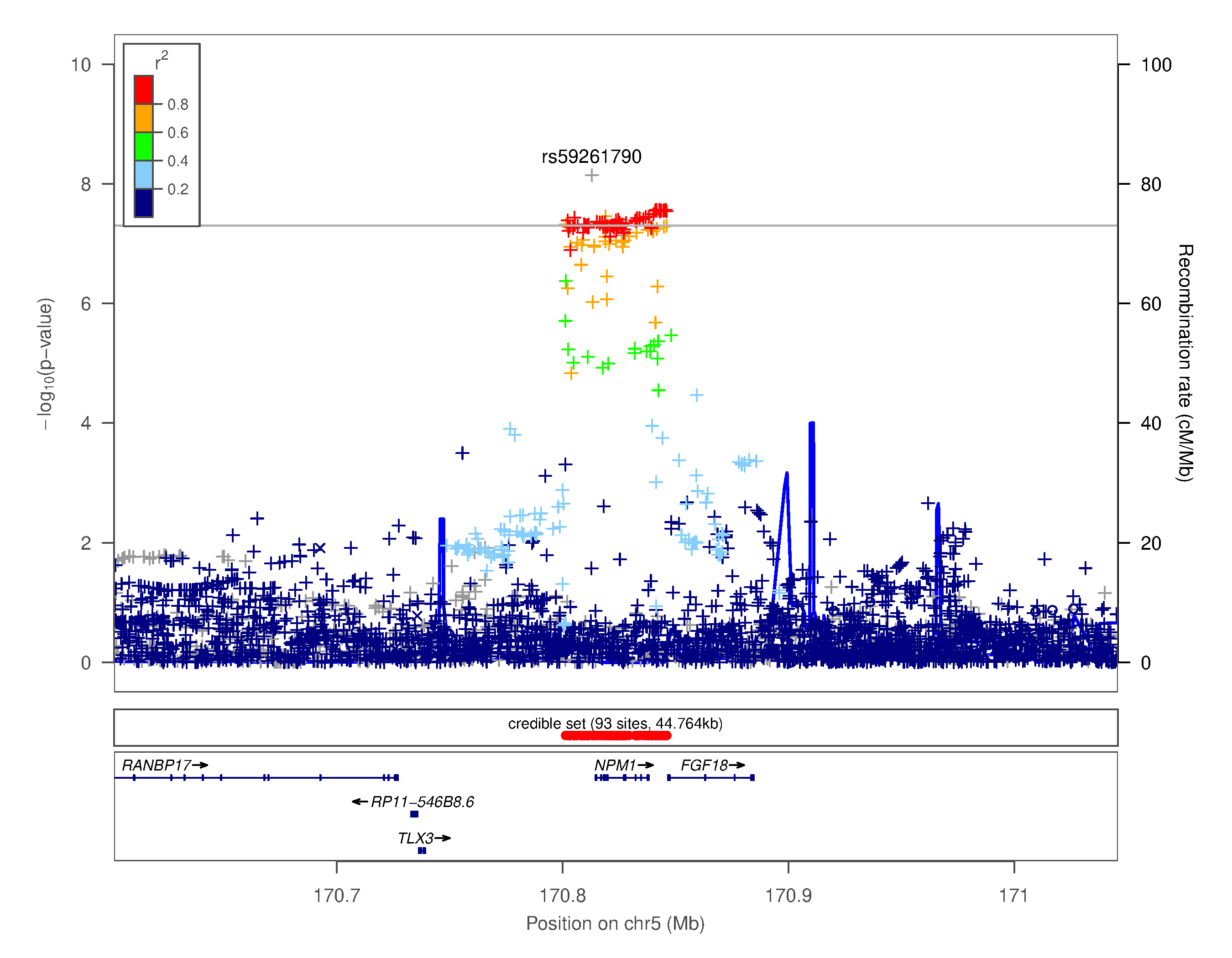

### Supplementary Figure 4.xxxii. Regional association plot for chr5q35.1 rs59261790

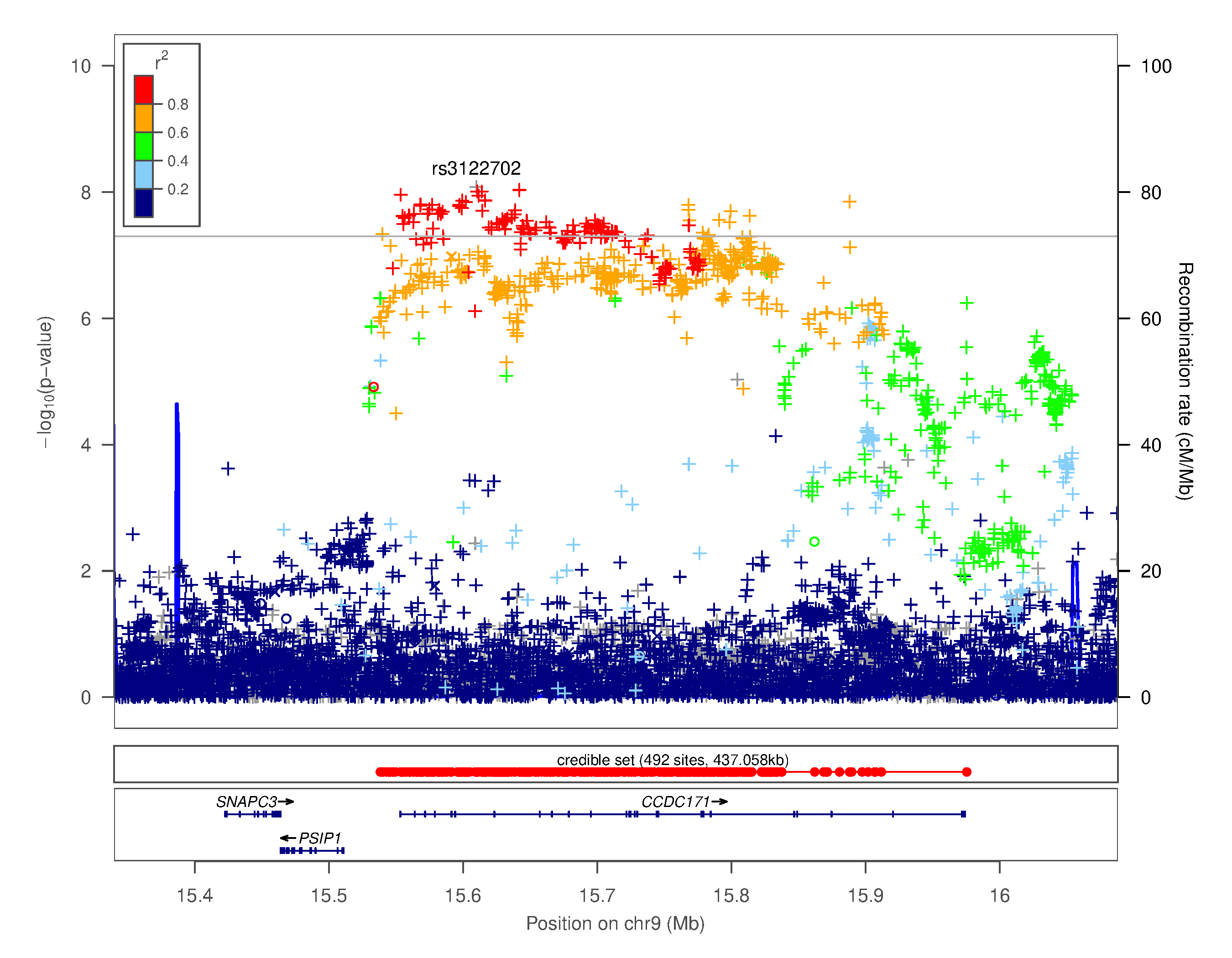

### Supplementary Figure 4.xxxiii. Regional association plot for chr9p22.3 rs3122702

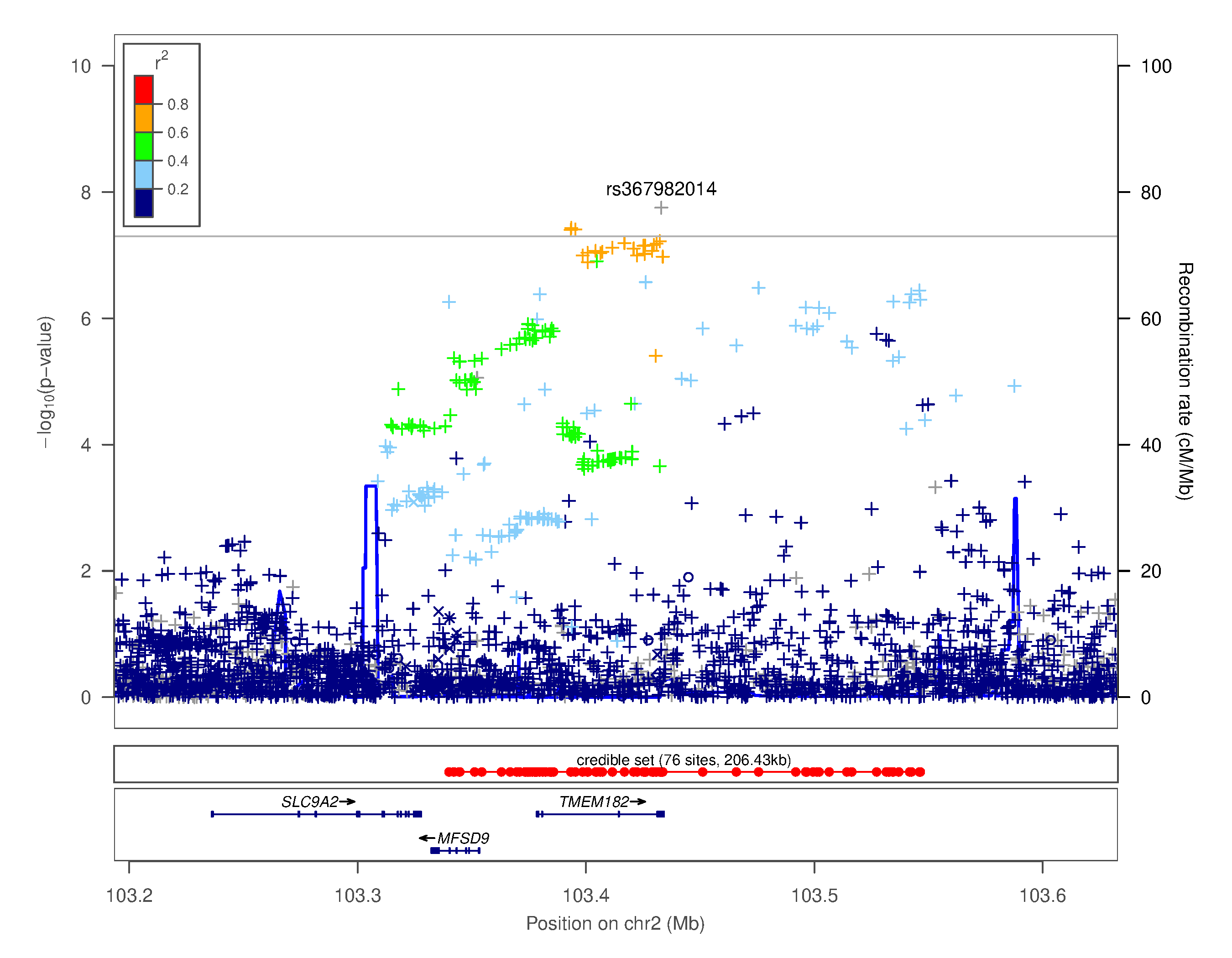

### Supplementary Figure 4.xxxiv. Regional association plot for chr2q12.1 rs367982014

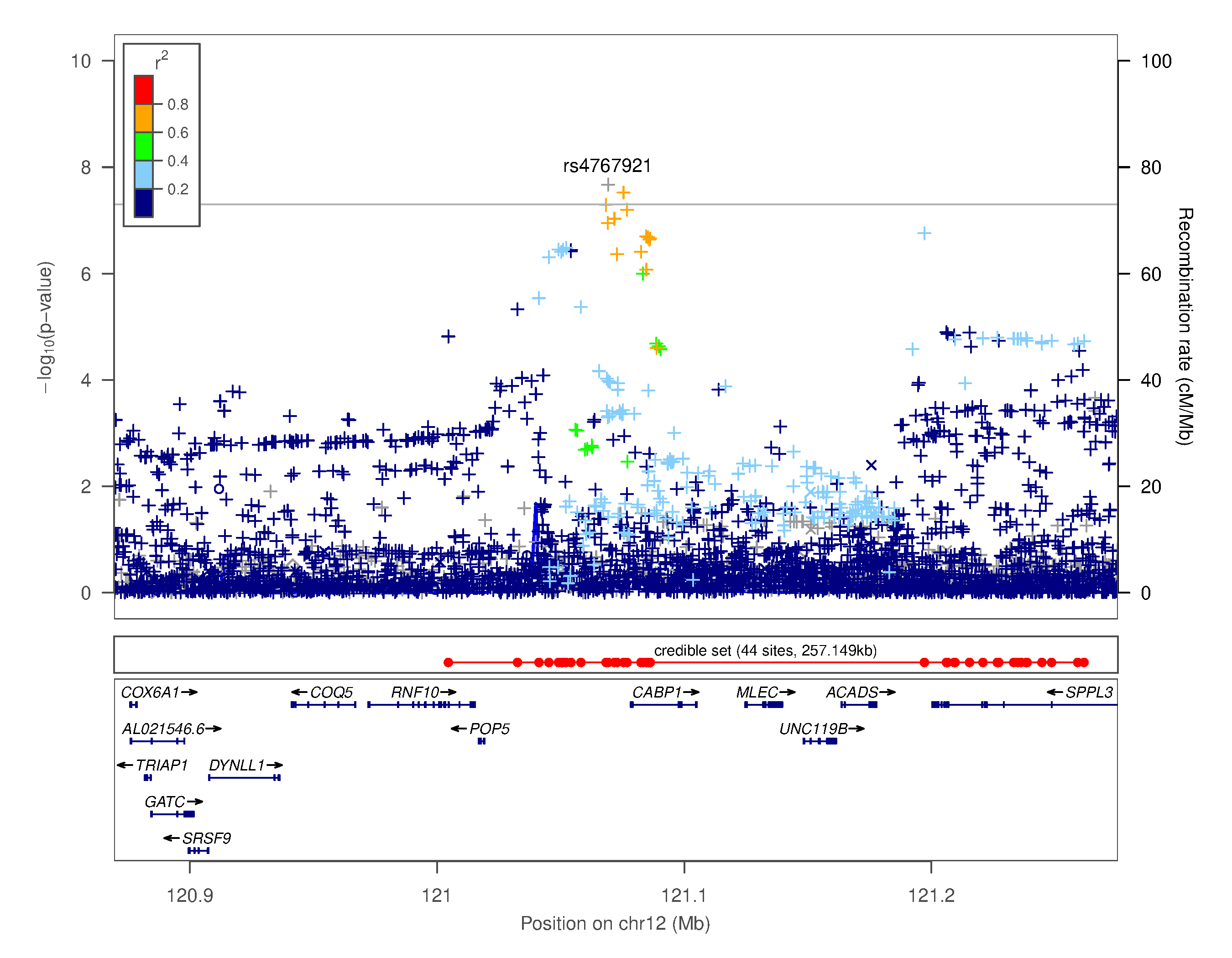

### Supplementary Figure 4.xxxv. Regional association plot for chr12q24.31 rs4767921

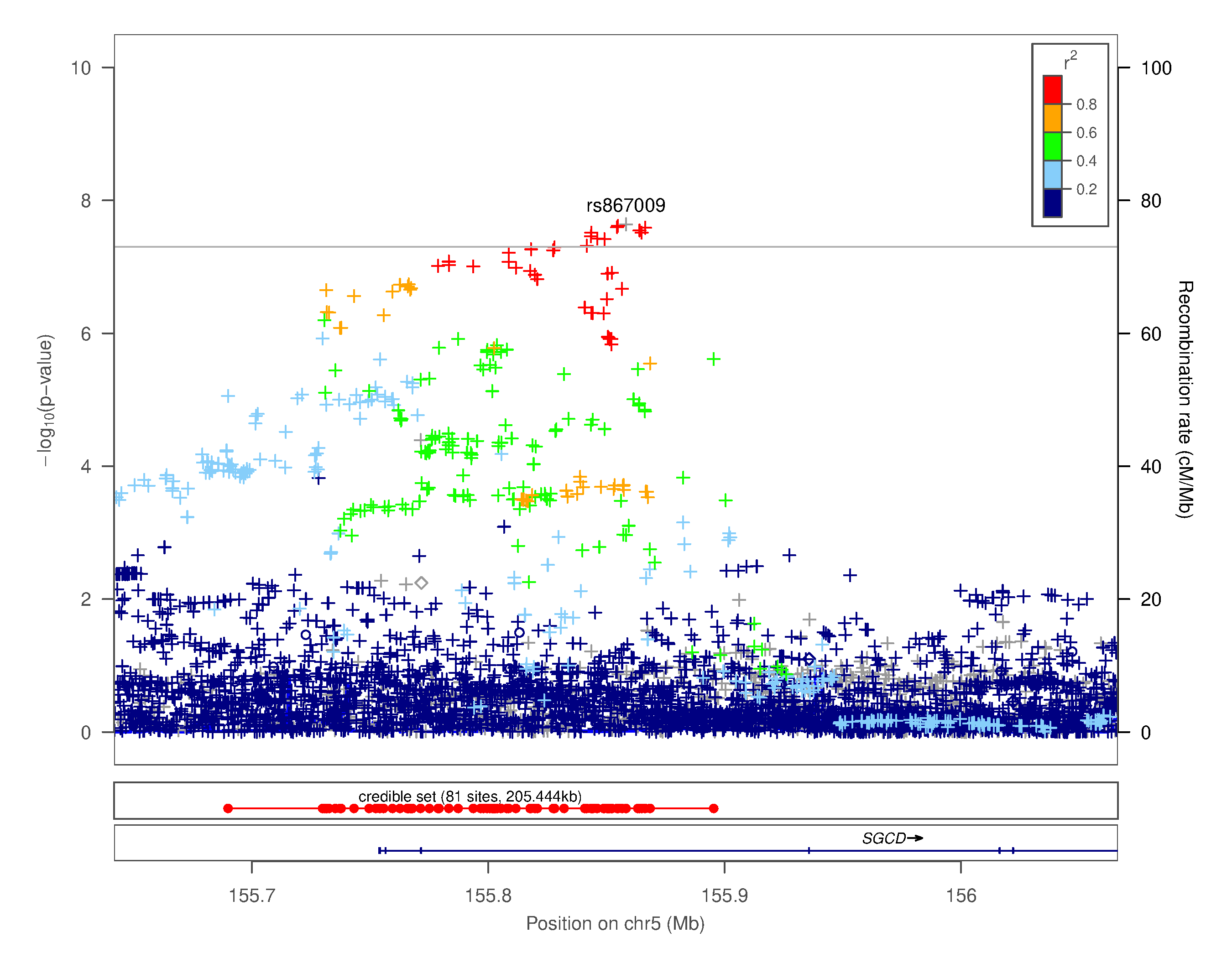

### Supplementary Figure 4.xxxvi. Regional association plot for chr5q33.3 rs867009

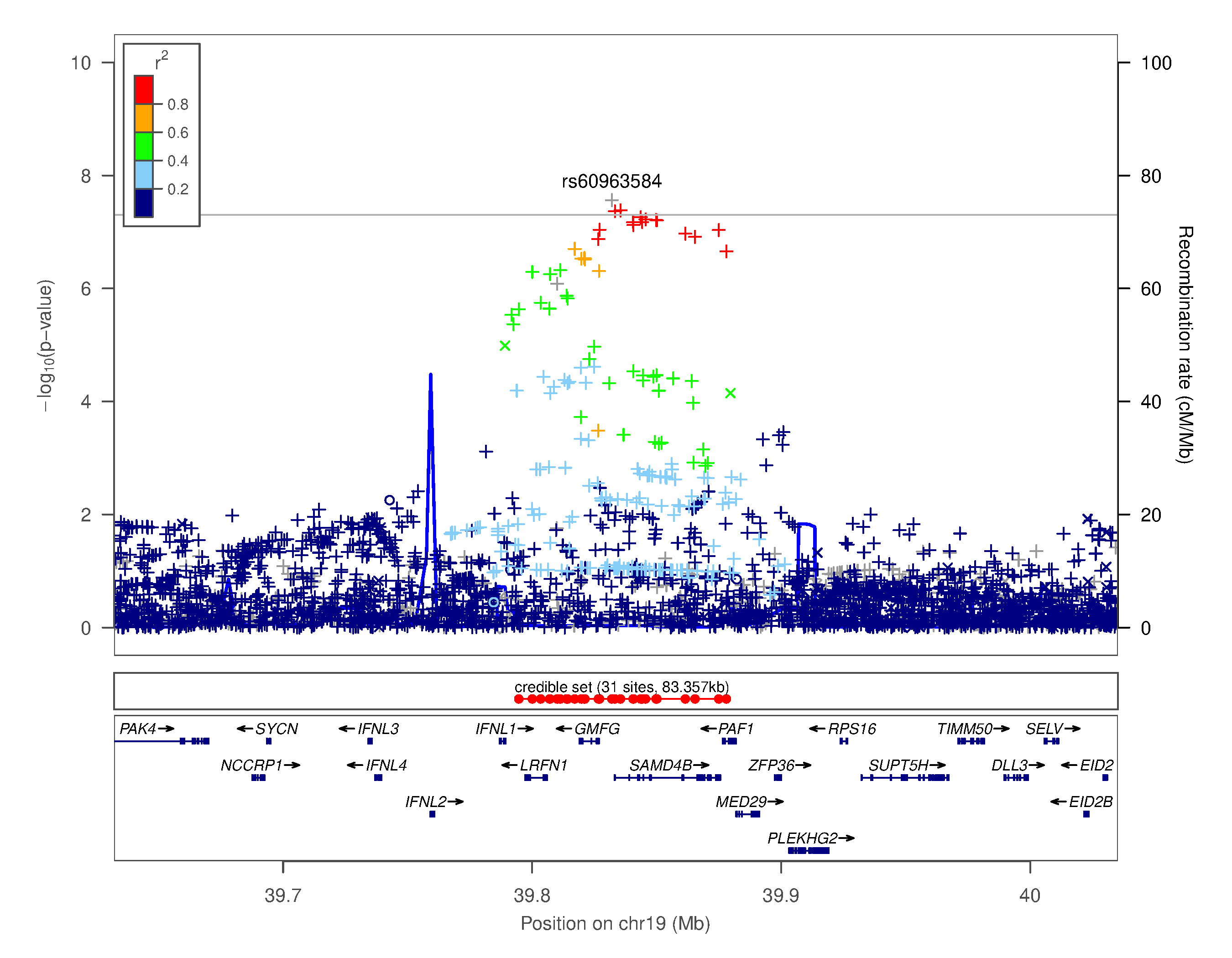

### Supplementary Figure 4.xxxvii. Regional association plot for chr19q13.2 rs60963584

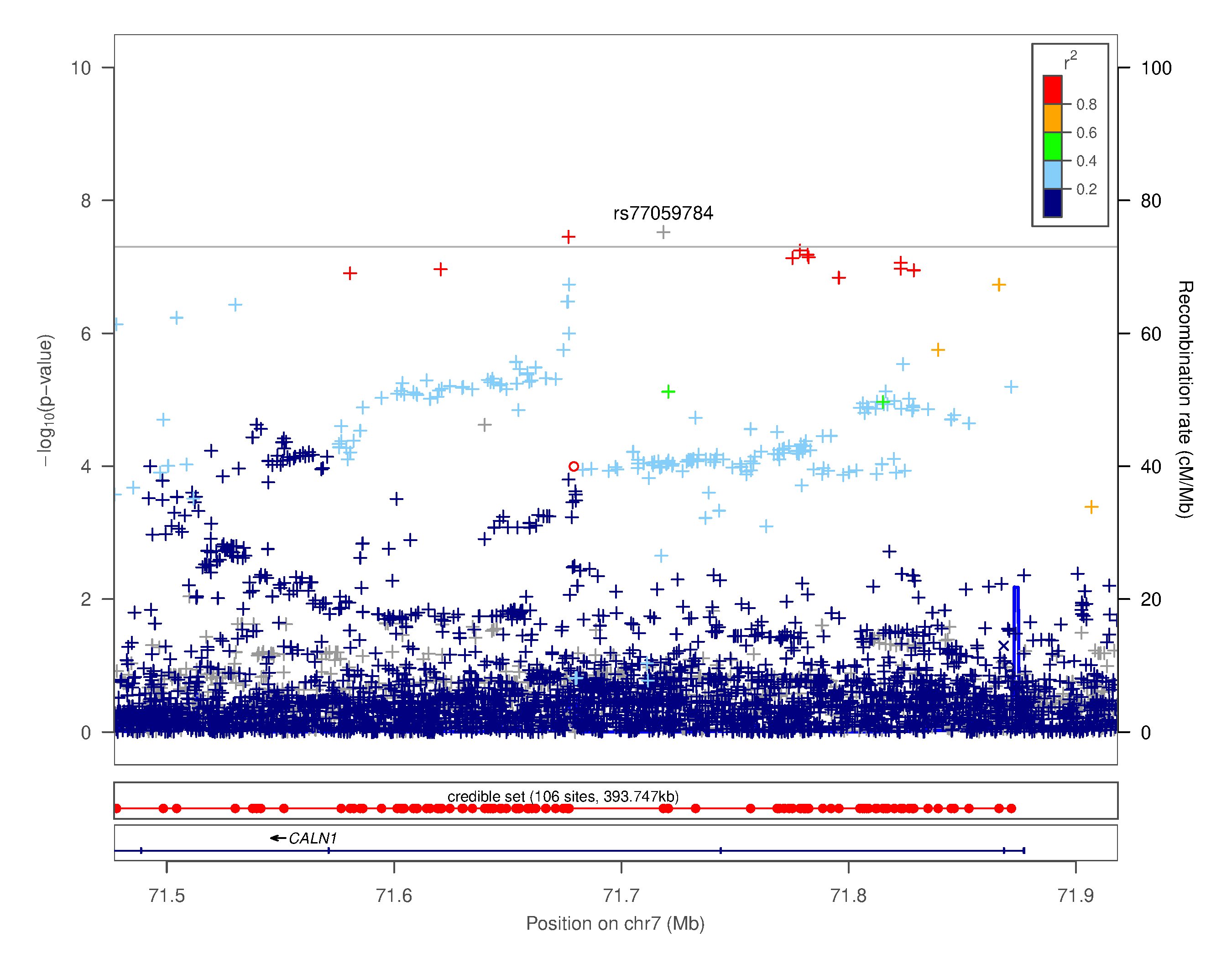

### Supplementary Figure 4.xxxviii. Regional association plot for chr7q11.22 rs77059784

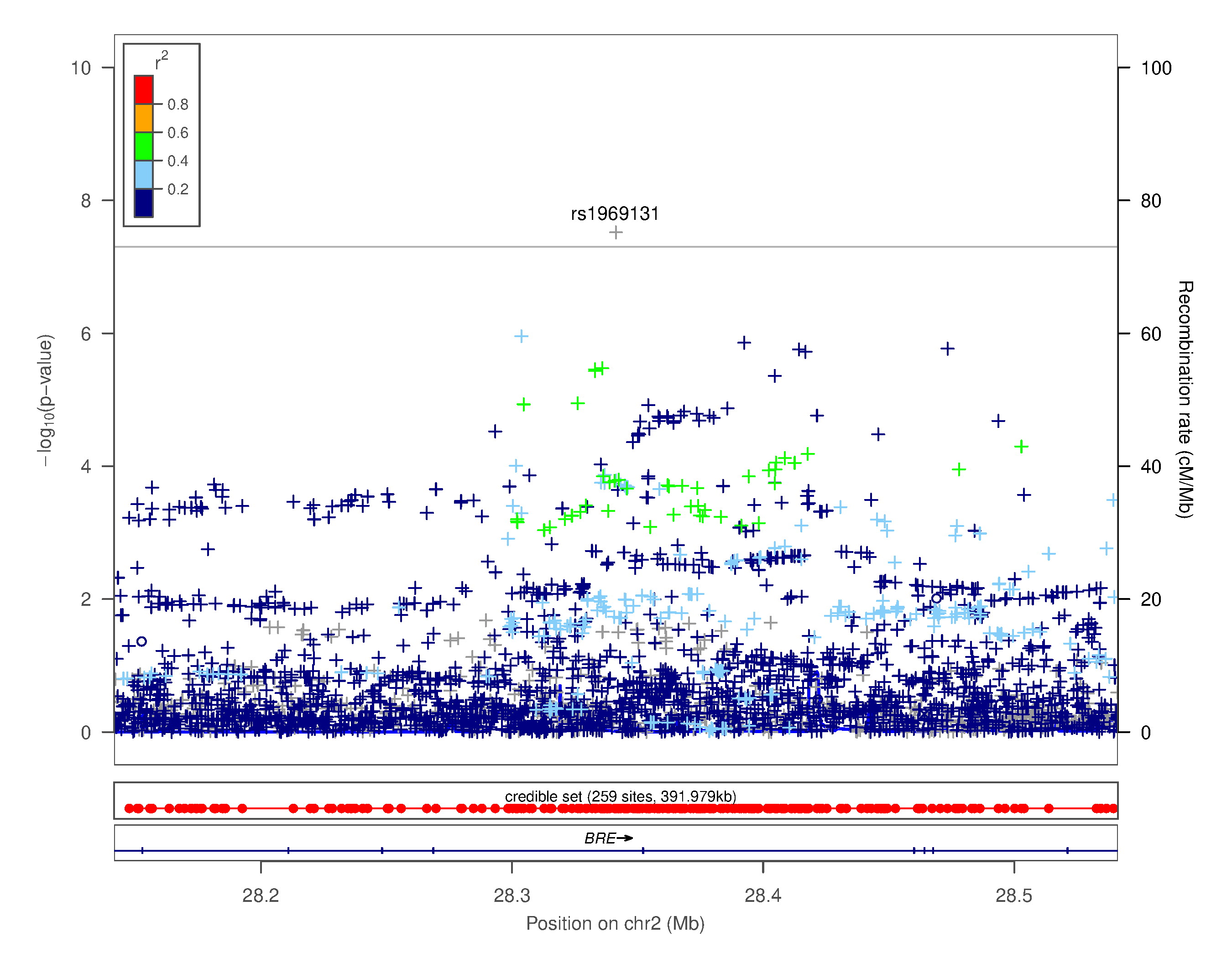

### Supplementary Figure 4.xxxix. Regional association plot for chr2p23.2 rs1969131

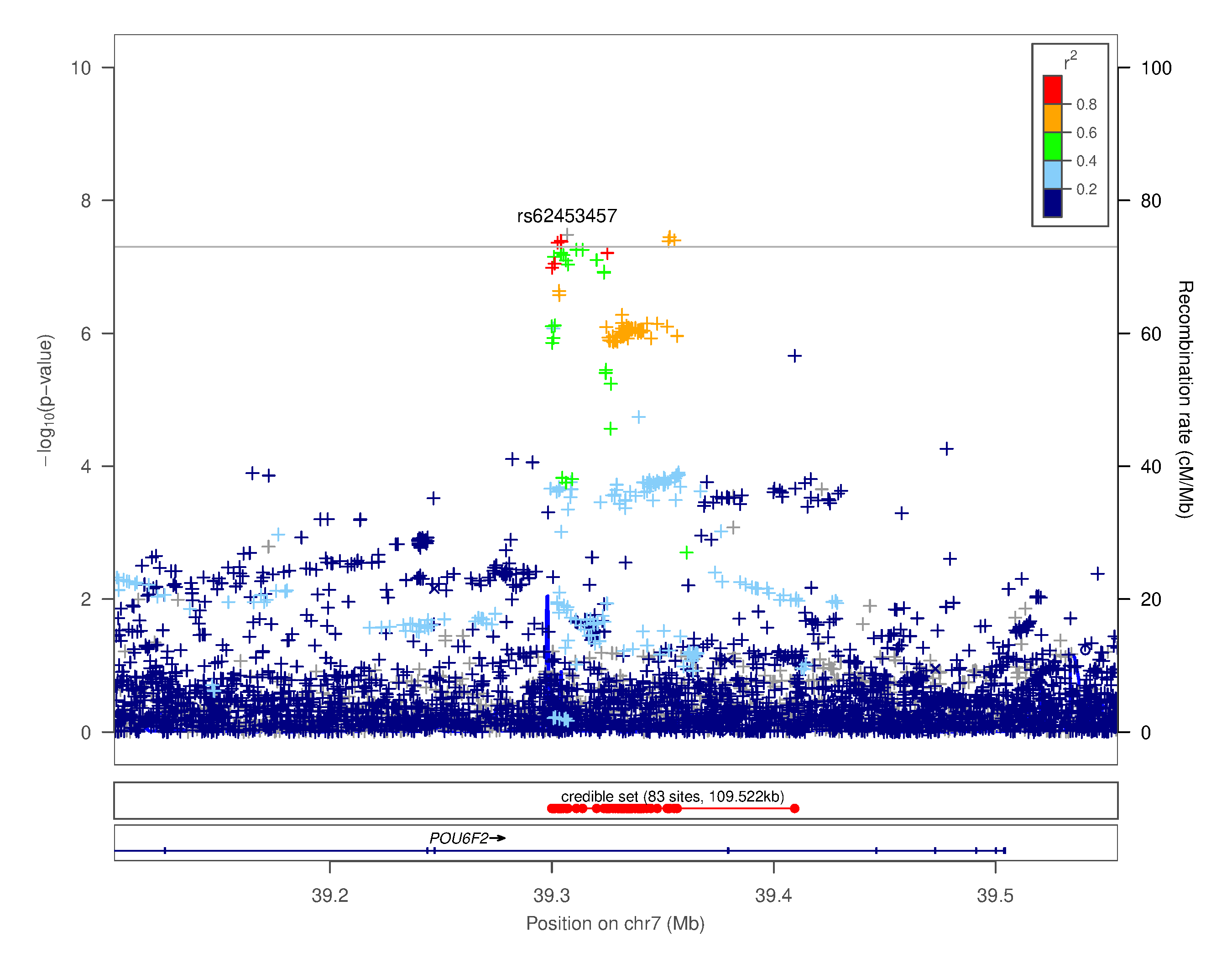

### Supplementary Figure 4.xl. Regional association plot for chr7p14.1 rs62453457

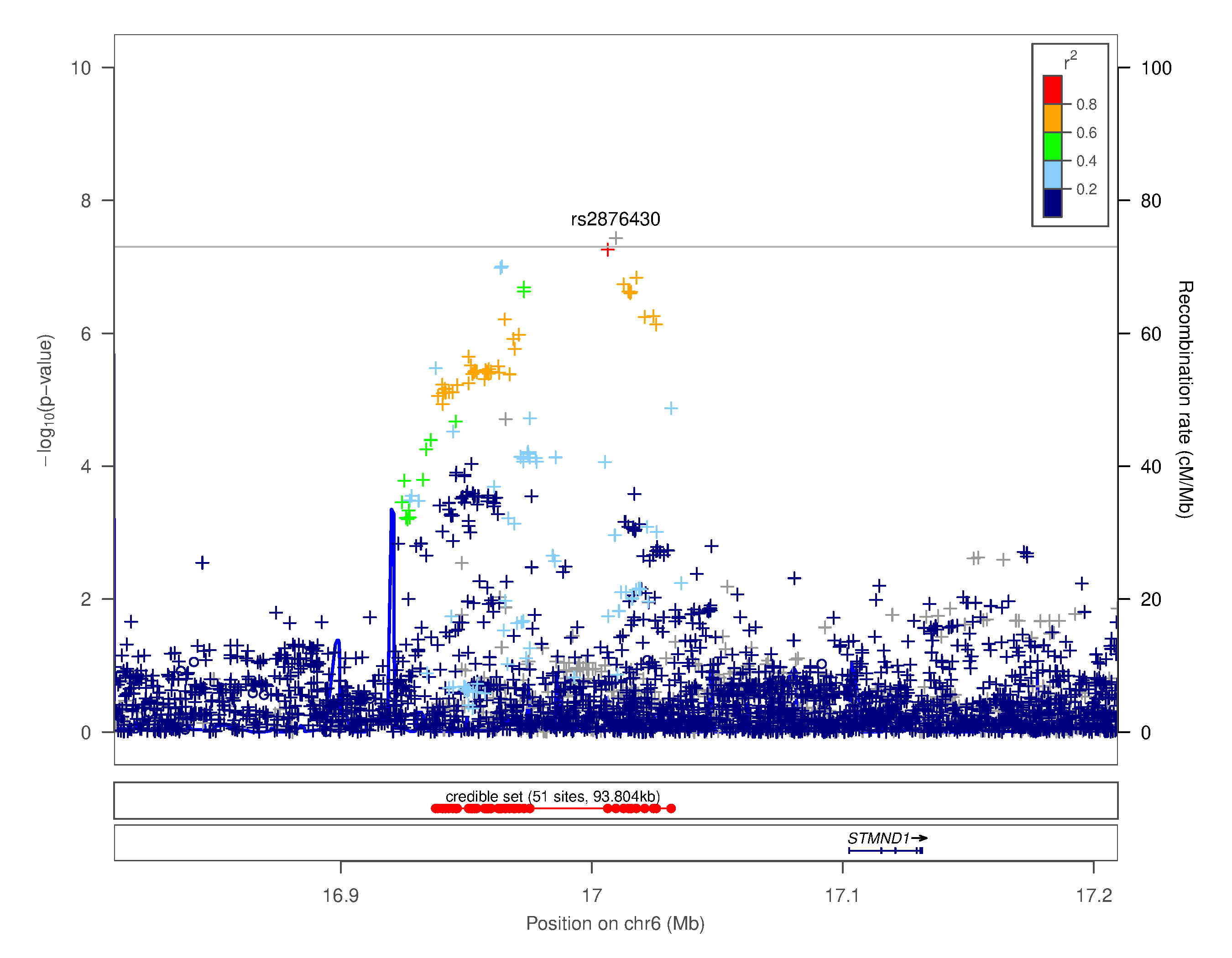

### Supplementary Figure 4.xli. Regional association plot for chr6p22.3 rs2876430

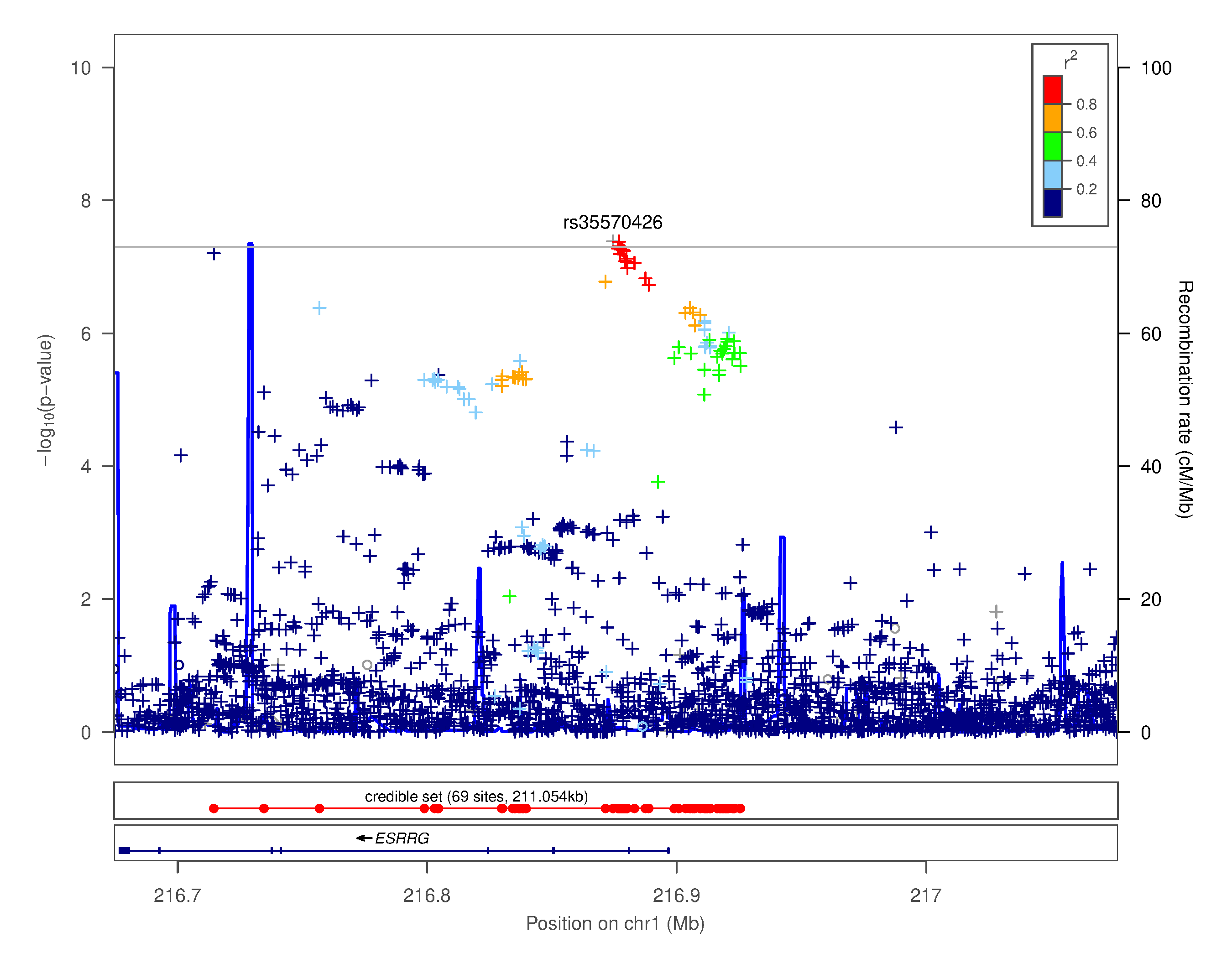

### Supplementary Figure 4.xlii. Regional association plot for chr1q41 rs35570426

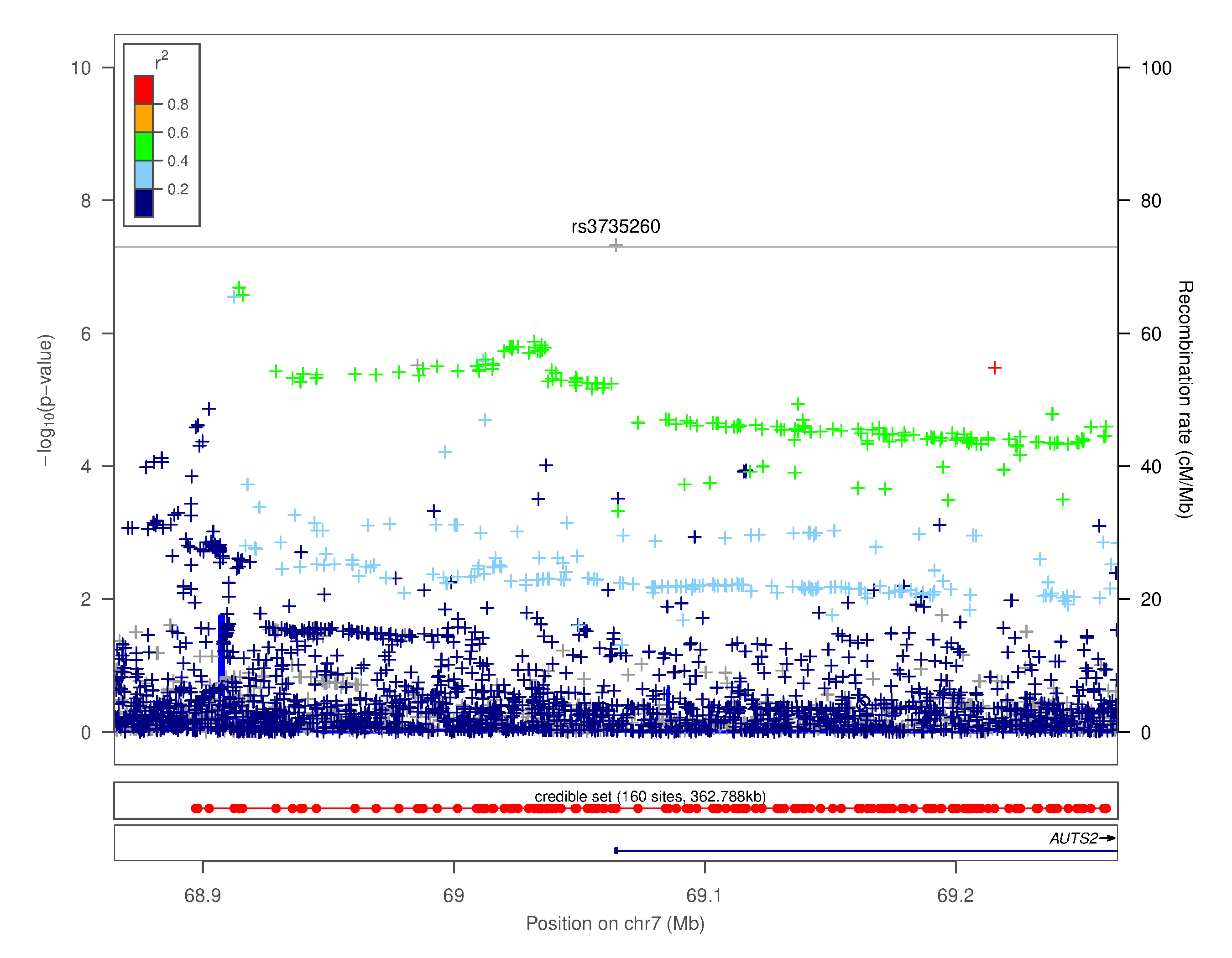

### Supplementary Figure 4.xliii. Regional association plot for chr7q11.22 rs3735260

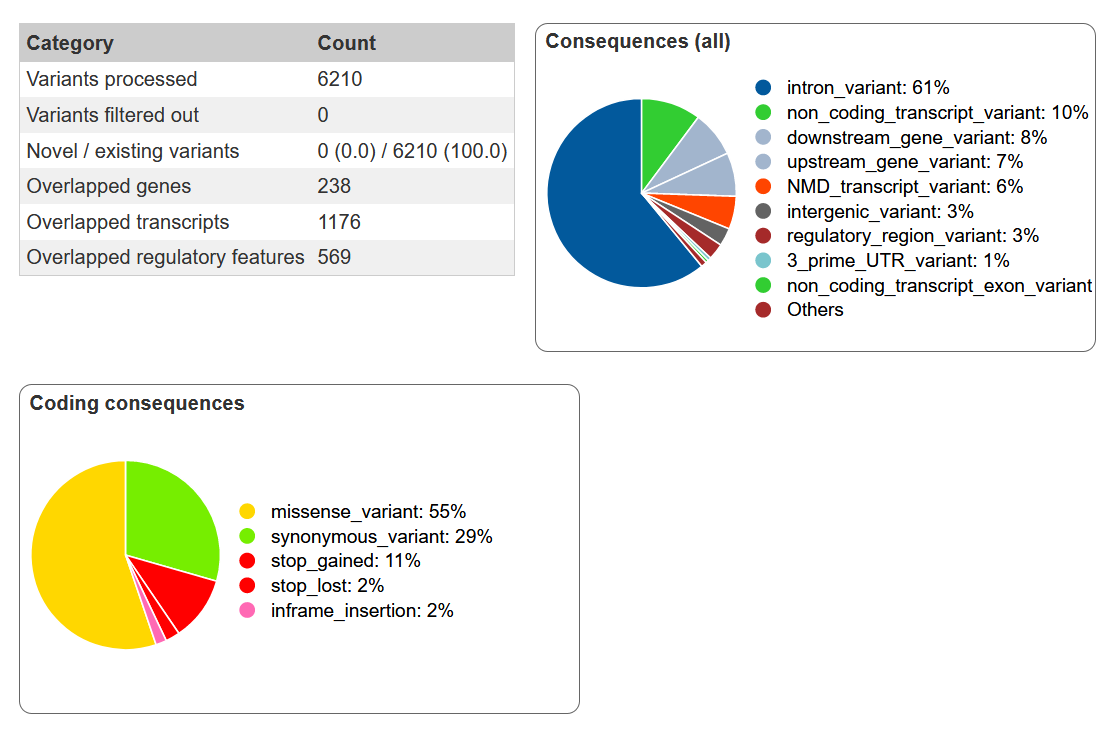

### Supplementary Figure 5 Variant effect predictor summary for the credible set of variants significantly associated with dyslexia

Summary information is output from the online variant effect predictor in ENSEMBL (release 104). All our variants were present in the 1000 Genomes reference panel so are considered existing, and no pre-filtering (e.g., on MAF; consequence type) was done.

**a)**

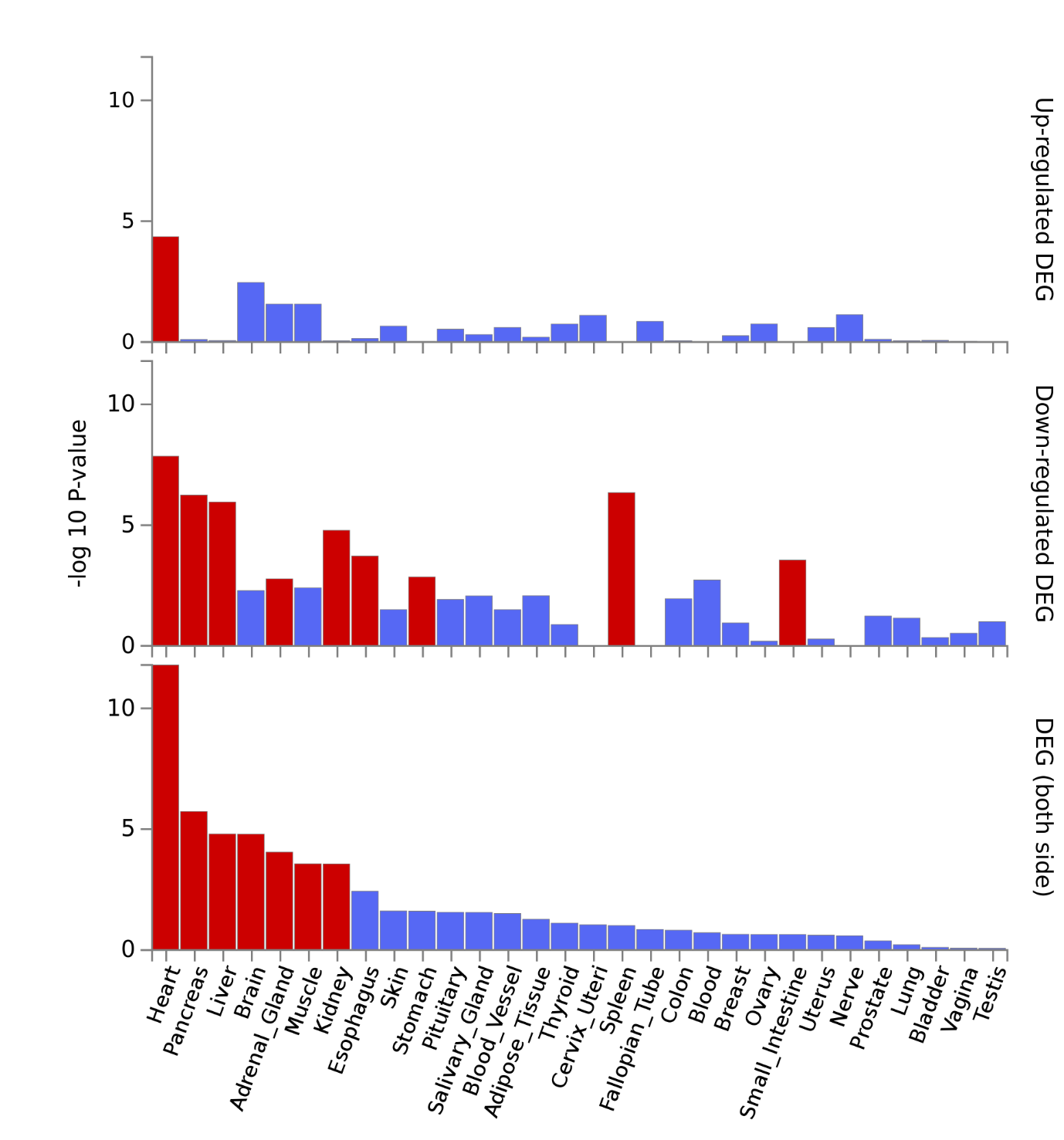

**b)**

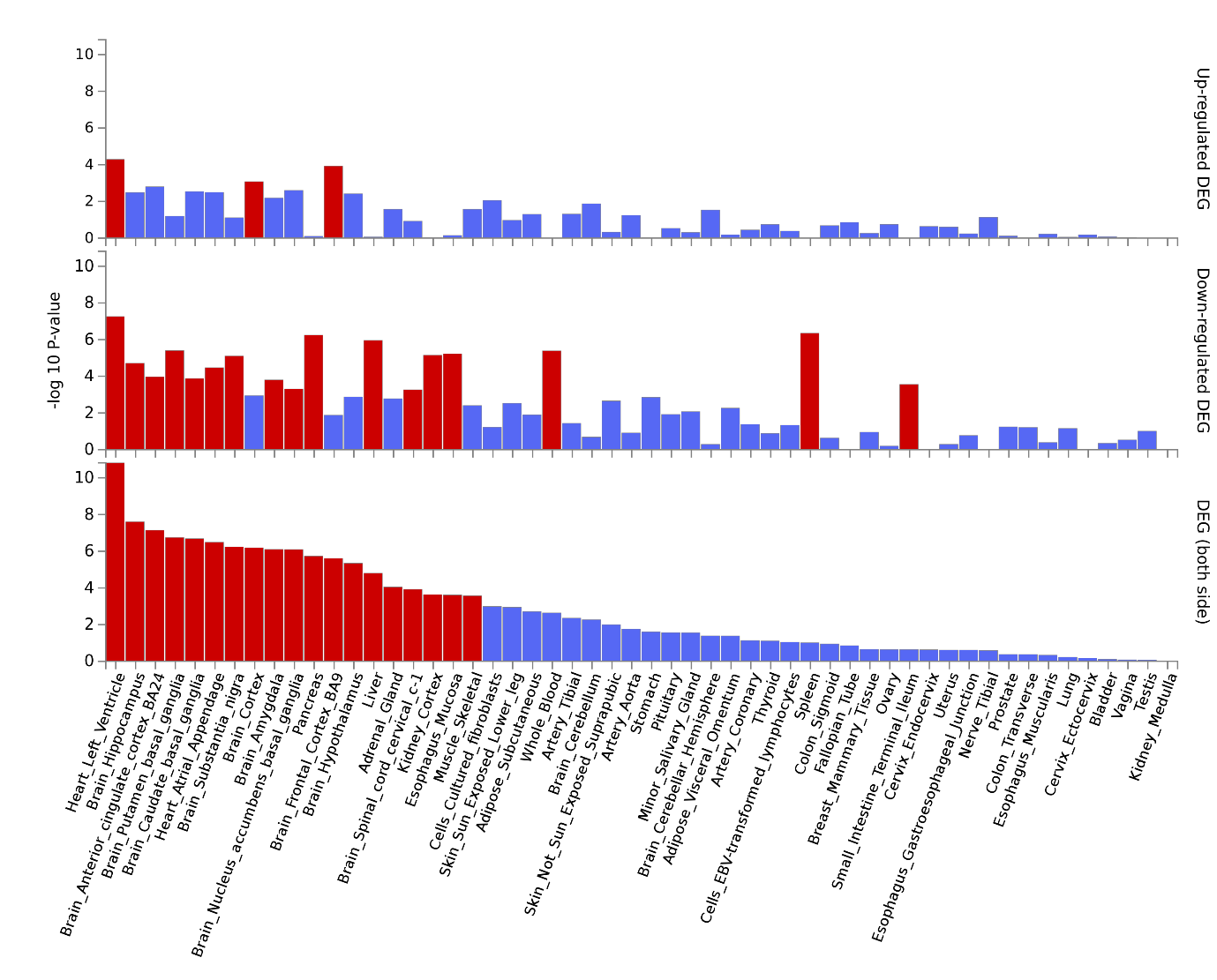

### Supplementary Figure 6 Differentially expressed gene sets across a) 30 general tissue types and b) 53 specific tissue types

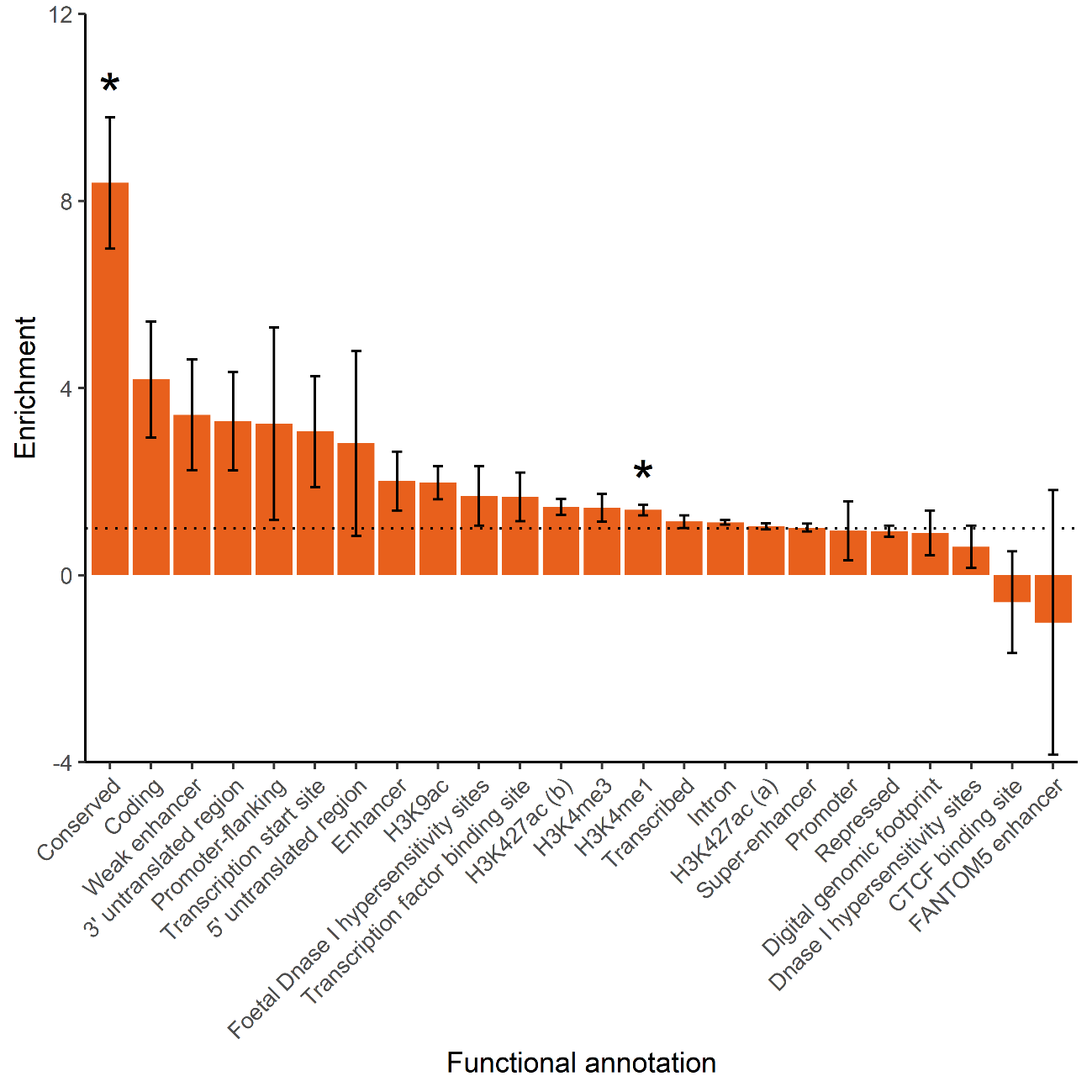

### Supplementary Figure 7 Enrichment estimates for major functional annotations

The 24 major functional annotations were defined by Finucane et al., (2015). Enrichment is the proportion of h^2^/proportion of SNPs. The horizontal dotted line indicates no enrichment (where proportion of h^2^/proportion of SNPs = 1). Error bars represent standard errors of the enrichment estimates. Asterisks indicate enrichment estimates are significant based on a Bonferroni-derived *p* value of < 2.08 x 10^-3^ (for 24 tests).

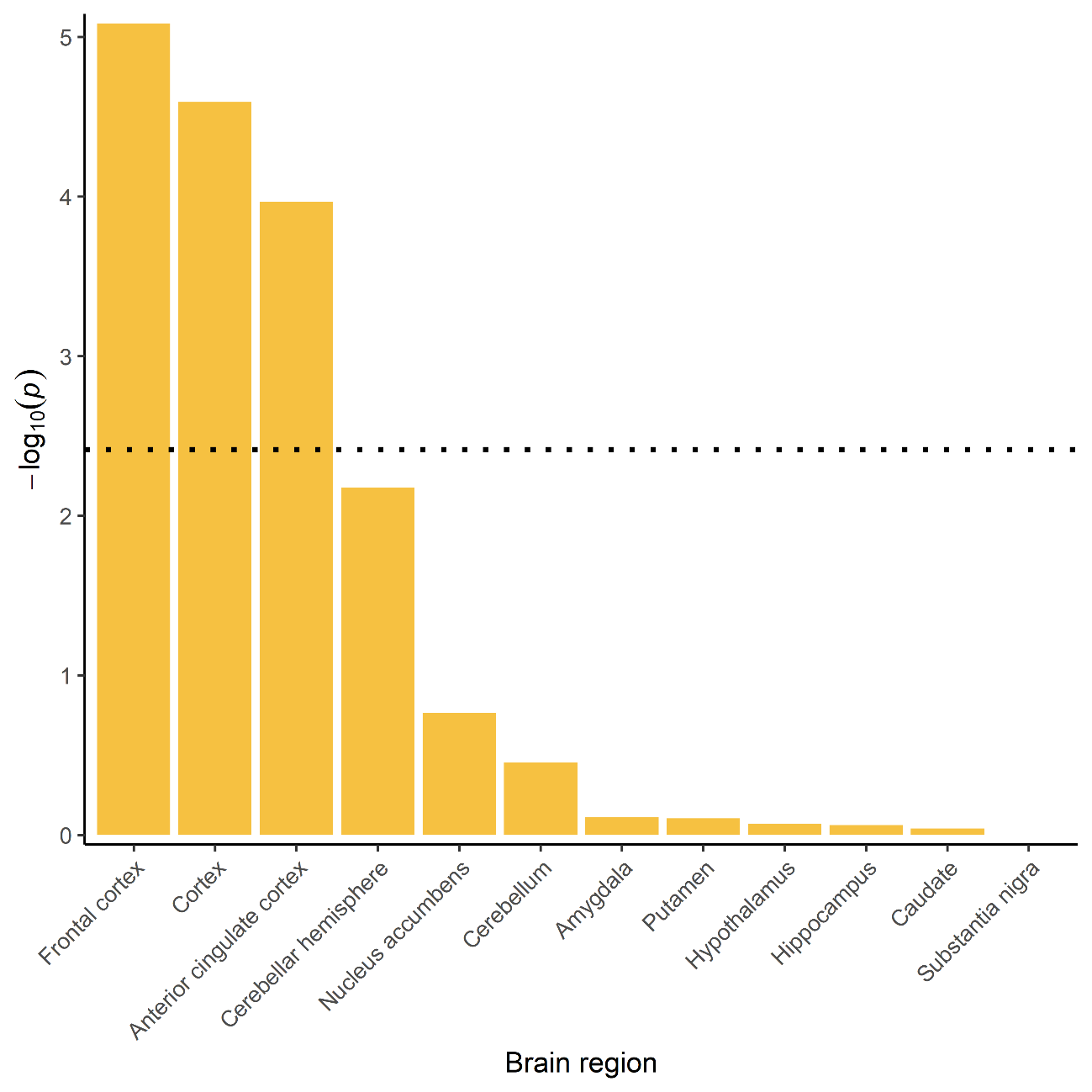

### Supplementary Figure 8 Heritability of dyslexia partitioned by brain tissue gene expression

The -log_10_(*p* value) of the enrichment estimates for heritability of dyslexia for genes expressed in 12 brain regions. The horizontal dotted line indicates significance after Bonferroni correction for 12 tests (*p* < 4.17 x 10^-3^).

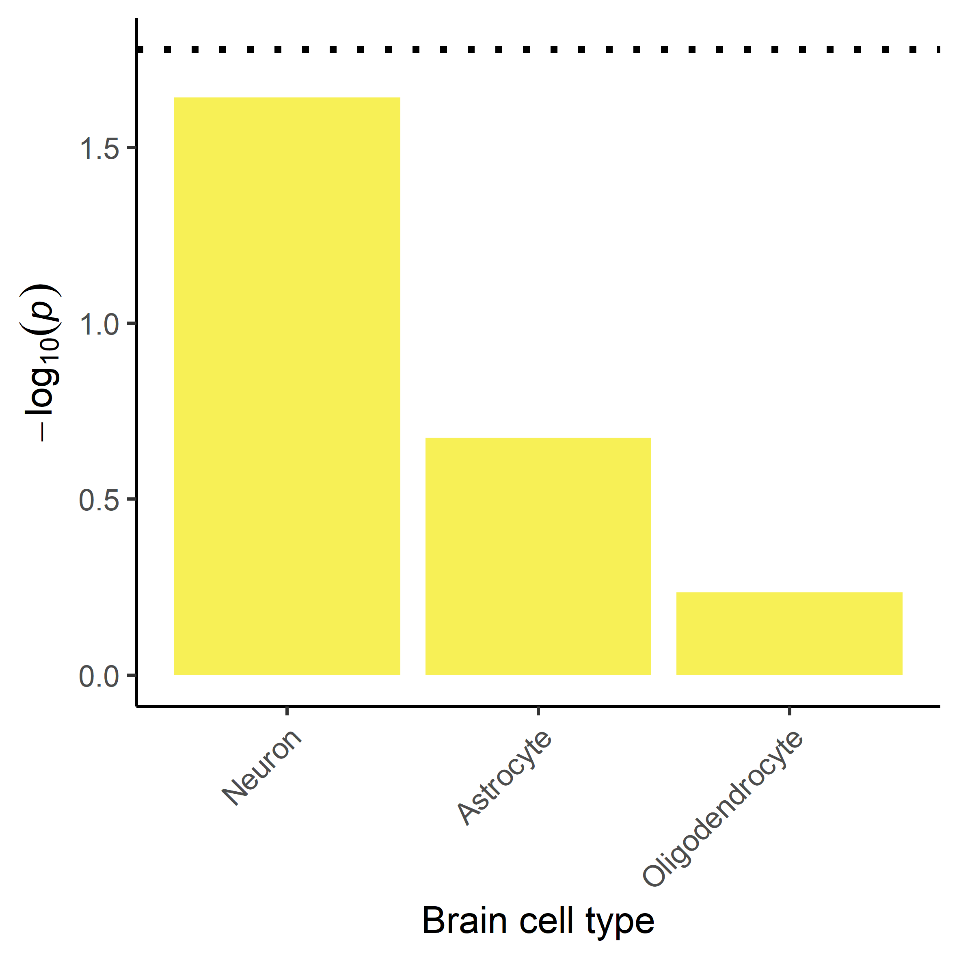

### Supplementary Figure 9 Heritability of dyslexia partitioned by brain cell type

The -log_10_(*p* value) of the enrichment estimates for heritability of dyslexia for brain cell types. The horizontal dotted line indicates significance after Bonferroni correction for three tests (*p* < 1.67 x 10^-2^).

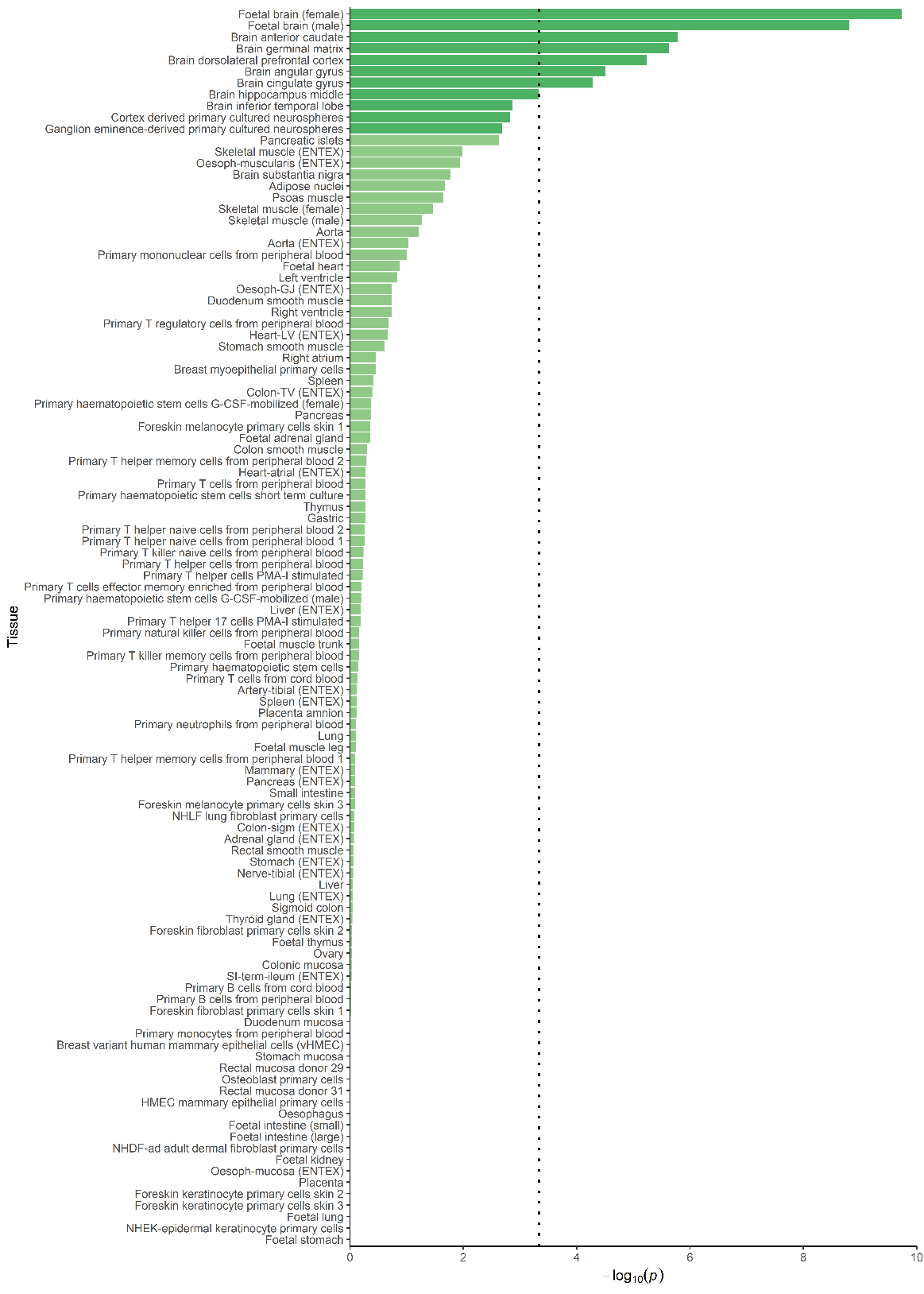

### Supplementary Figure 10 Heritability of dyslexia partitioned by cell-type specific H3K4me1

The -log_10_(*p* value) of the enrichment estimates for heritability of dyslexia for variants located within H3K4me1 peaks of different tissues. Central nervous systems tissues are represented in dark green and other tissues are represented in light green. The vertical dotted line indicates significance after Bonferroni correction for 114 tests (*p* < 4.39 x 10^-4^).

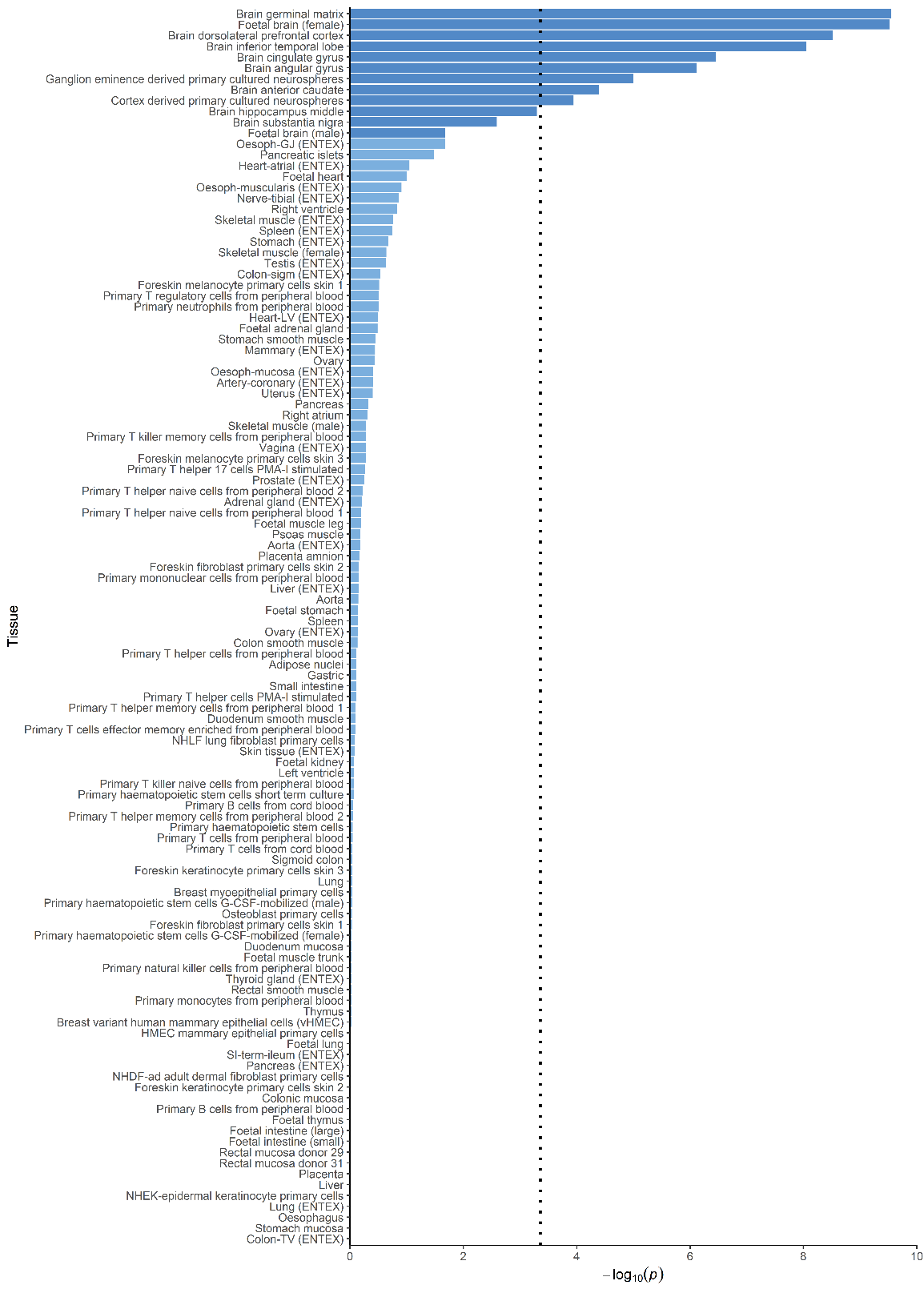

### Supplementary Figure 11 Heritability of dyslexia partitioned by cell-type specific H3K4me3

The -log_10_(*p* value) of the enrichment estimates for heritability of dyslexia for variants located within H3K4me3 peaks of different tissues. Central nervous systems tissues are represented in dark blue and other tissues are represented in light blue. The vertical dotted line indicates significance after Bonferroni correction for 114 tests (*p* < 4.39 x 10^-4^).

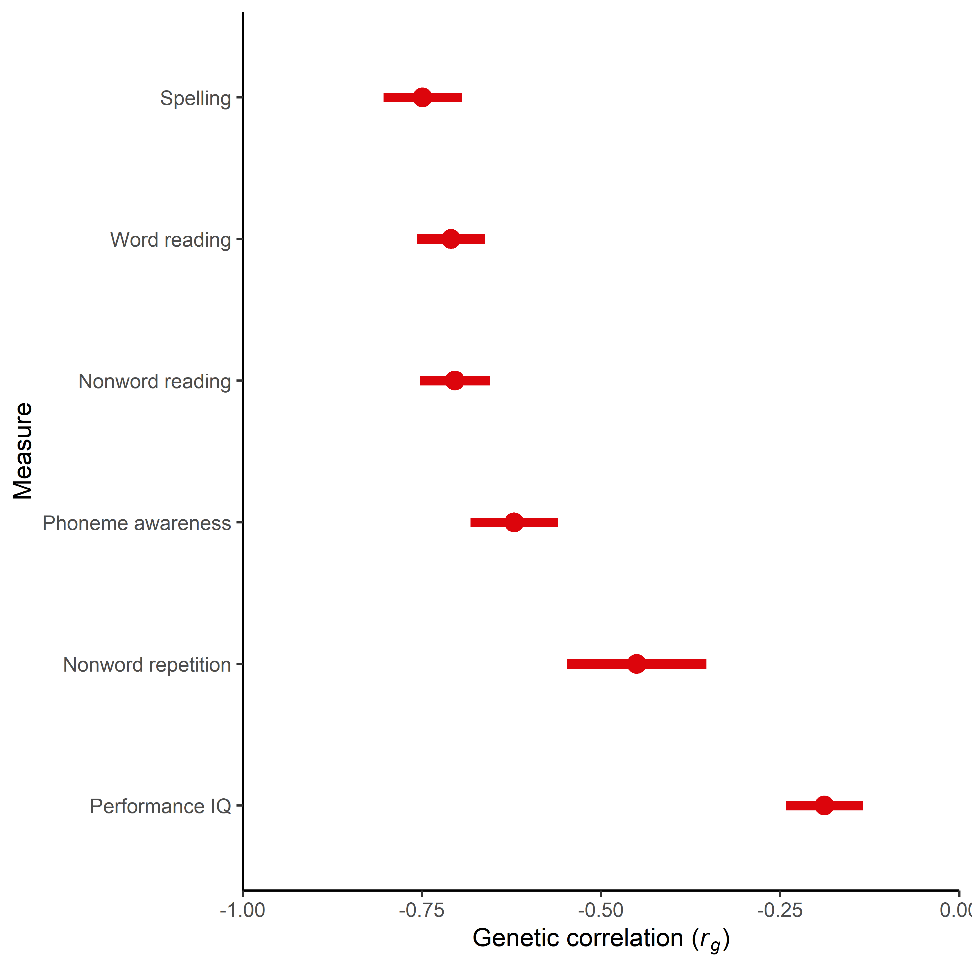

### Supplementary Figure 12 Genetic correlations between dyslexia and measures of reading, language, and nonverbal IQ

Genetic correlations (*r*_g_) between self-reported dyslexia diagnosis from 23andMe and measures of reading, language, and nonverbal IQ in the GenLang consortium. Points represent genetic correlation estimated in LDSC and error bars represent 95% confidence limits.

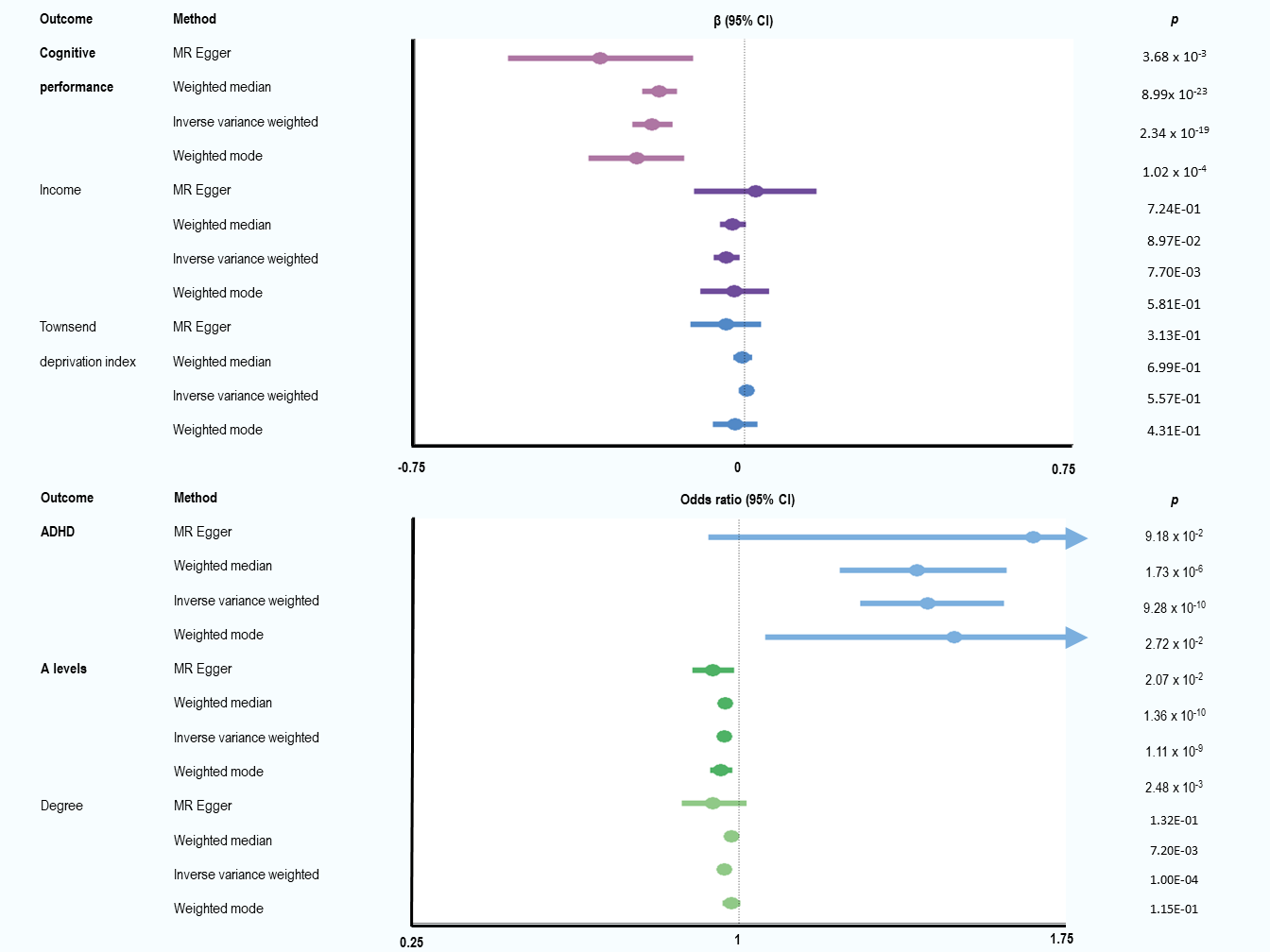

### Supplementary Figure 13 Mendelian randomisation analysis showing the effects of dyslexia on general cognitive ability, ADHD, and educational attainment outcomes

Exposure variants were selected using a significance threshold of *p* < 5 x 10^-8^. The number of variants significantly associated with dyslexia available in the outcome summary statistics was 51 in all cases. The effect of dyslexia on the continuous outcome is given in standardised beta values (top panel) and the effect of dyslexia on the binary outcomes (bottom panel) is given in odds ratios. Error bars represent the 95% confidence intervals of the effect and arrows indicate where confidence intervals extend beyond the axis limit. The vertical dotted line in each panel divides positive from negative effects. Effects are significant where *p* < .05.
