## Supplementary Methods for "Discovery of 42 Genome-Wide Significant Loci Associated with Dyslexia"

Catherine Doust^1^, Pierre Fontanillas^2^, Else Eising^3^, Scott D Gordon^4^, Zhengjun Wang^5^, Gökberk Alagöz^3^, Barbara Molz^3^, 23andMe Research Team^2^, Quantitative Trait Working Group of the GenLang Consortium, Beate St Pourcain^3,6^, Clyde Francks^3,6^, Riccardo E Marioni^7^, Jingjing Zhao^5^, Silvia Paracchini^8^, Joel B Talcott^9^, Anthony P Monaco^10^, John F Stein^11^, Jeffrey R Gruen^12^, Richard K Olson^13^, Erik G Willcutt^13^, John C DeFries^13^, Bruce F Pennington^14^, Shelley D Smith^15^, Margaret J Wright^16^, Nicholas G Martin^4^, Adam Auton^2^, Timothy C Bates^1^, Simon E Fisher^3,6^ & Michelle Luciano^1^

^1^ Department of Psychology, The University of Edinburgh, Edinburgh, EH8 9JZ, UK

^2^ 23andMe, Inc., Sunnyvale, CA, USA

^3^ Language and Genetics Department, Max Planck Institute for Psycholinguistics, 6500 AH, Nijmegen, The Netherlands

^4^ Genetic Epidemiology Laboratory, QIMR Berghofer Medical Research Institute, Brisbane, 4029, Queensland, Australia

^5^ School of Psychology, Shaanxi Normal University and Shaanxi Key Research Center of Child Mental and Behavioral Health, Xi’an, 710062, China

^6^ Donders Institute for Brain, Cognition and Behaviour, 6500 EH, Nijmegen, The Netherlands

^7^ Centre for Genomic & Experimental Medicine, Institute of Genetics & Cancer, University of Edinburgh, Edinburgh, EH4 2XU, UK

^8^ School of Medicine, Medical & Biological Sciences, University of St Andrews, St Andrews, KY16 9TF, UK

^9^ Developmental Cognitive Neuroscience, Aston Brain Centre, Birmingham, B4 7ET, UK

^10^ Office of the President, Tufts University, Medford, Massachusetts, 02155, USA

^11^ Department of Physiology, Anatomy and Genetics, Oxford University, **Oxford, OX1 3PT, UK**

^12^ Departments of Pediatrics and Genetics, Yale Medical School, New Haven, Connecticut, 06520-8081, USA

^13^ Department of Psychology and Neuroscience, University of Colorado, Boulder, Colorado, 80309-0345, USA

^14^ Department of Psychology at the University of Denver, Denver, Colorado, 80208, USA

^15^ Department of Neurological Sciences, College of Medicine, University of Nebraska Medical Center, Omaha, Nebraska, 68198-8440, USA

^16^ Queensland Brain Institute, University of Queensland, Brisbane, 4072, Queensland, Australia

#### **23andMe Genotyping and imputation**

Samples were genotyped on one of five genotyping platforms. The V1 and V2 platforms were variants of the Illumina HumanHap550 + BeadChip, including about 25,000 custom SNPs selected by 23andMe, with a total of about 560,000 SNPs. The V3 platform was based on the Illumina OmniExpress + BeadChip, with custom content to improve the overlap with our V2 array, with a total of ~950,000 SNPs. The V4 platform is a fully custom array, including a lower redundancy subset of V2 and V3 SNPs with additional coverage of lower-frequency coding variation, and ~570,000 SNPs. The v5 platform, in current use, is an Illumina Infinium Global Screening Array (~640,000 SNPs) supplemented with ~50,000 SNPs of custom content. Samples that failed to reach 98.5% call rate were excluded from the study.

Individuals were only included if they had > 97% European ancestry, as determined through an analysis of local ancestry. Briefly, this analysis first partitions phased genomic data into short windows of ~100 SNPs. Within each window, a support vector machine is used to classify individual haplotypes into one of 31 reference populations. The support vector machine classifications are then fed into a hidden Markov model (HMM) that accounts for switch errors and incorrect assignments and gives probabilities for each reference population in each window. Finally, simulated admixed individuals are used to recalibrate the HMM probabilities so that the reported assignments are consistent with the simulated admixture proportions. The reference population data are derived from public data sets (the Human Genome Diversity Project, HapMap and 1000 Genomes) and from 23andMe research participants who have reported having four grandparents from the same country.

A maximal set of unrelated individuals was chosen for each analysis using a segmental identity-by-descent (IBD) estimation algorithm [1]. Individuals were defined as related if they shared more than 700 cM IBD, including regions where the two individuals share either one or both genomic segments identical-by-descent. This level of relatedness (roughly 20% of the genome) corresponds approximately to the minimal expected sharing between first cousins in an outbred population. For the purposes of GWAS, if a case was found to be related to a control, the case was preferentially kept in the sample.

Participant genotype data were imputed against a single unified imputation reference panel, combining the May 2015 release of the 1000 Genomes Phase 3 haplotypes and the UK10K imputation reference panel. Data for each genotyping platform were phased and imputed separately. Variants that were only genotyped on the ‘V1’ platform were flagged due to small sample size, and variants on chrM or chrY, because many of these are not currently called reliably. Using trio data, variants that failed a test for parent–offspring transmission were also flagged; specifically, the child’s allele count was regressed against the mean parental allele count and variants with fitted β < 0.6 and *p* < 10^-20^ for a test of β<1 were flagged. Variants with a Hardy–Weinberg *p* < 10^-20^ in Europeans, or a call rate of < 90%, were also flagged. Genotyped variants were also tested for batch effects and variants with *p* < 10^-50^ by analysis of variance of genotypes against a factor dividing genotyping date into 20 roughly equal-sized buckets were flagged. For imputed GWAS results, variants with average *r*^2^ < 0.5 or minimum *r*^2^ < 0.3 in any imputation batch were flagged, as well as SNPs that had strong evidence of an imputation batch effect, using an analysis of variance of the imputed dosages against a factor representing imputation batch; results with *p* < 10^-50^ were flagged. Each variant flagged by QC on genotyped or imputation data were excluded from the GWAS analysis.

#### **Chinese Reading Study sample**

##### *Participants*

3,127 Grade 3 to Grade 6 primary students aged nine to 14 years were recruited from three cities and four districts in China (Xi’an-YT, Xi’an-CB, Qingyang, and Baotou). In total, 2,476 participants were eligible for subsequent genotyping and association analysis. Ethical approval was obtained for each cohort at the local level and written informed consent was obtained from all the participants’ parents.

##### *Phenotypic measures*

Reading accuracy: A Chinese character recognition test was employed to measure each child’s reading accuracy [2-4]. The test consisted of 150 single Chinese characters selected from China’s Elementary School Textbooks (1996). The average frequency of the characters was 182 per million (ranging from 0 to 2,282), and the reliability of this test was 0.95 [2]. Each child was individually tested and required to read aloud each character at a time.

Reading fluency: A word list reading task [2] was used to measure each child’s reading fluency. In this task, children were asked to name a list of 180 two-character words as rapidly and accurately as possible. All these words were from primary school textbooks and have been learned before Grade 3, such as “我们 (we)” and “太阳 (sun)”. The mean frequency of these words was 212.77 per million [5]. Since words included in this task were all simple, this task was administrated to test children’s reading fluency. The total time for naming the whole word list was recorded as the measurement of reading fluency.

##### *Genotype quality control, imputation, and analysis*

DNA was extracted from saliva samples, and individuals were genotyped using the Illumina Asian screening array (650K) by Beijing Compass Biotechnology. Quality control was performed using standard quality control metrics. Eight samples were excluded as they had sex discrepancies between the records and the genetically inferred data [6, 7]. Next, we removed 53 samples who had unexpected duplicates or probable relatives (PI-HAT > 0.20). Then, SNPs were filtered out if they showed a variant call rate < 0.95, a minor allele frequency (MAF) < 0.01, a missing genotype data (mind) < 0.90, or a Hardy-Weinberg equilibrium (HWE) < 10^-5^ within each dataset.

For imputation, autosomal variants were aligned to the 1000G genomes phase 1v3 reference panel. Imputation was performed using the Michigan imputation Server 4.0 in 5Mb chunks with 500kb buffers, filtering out variants that were monomorphic in the Genome Asia Pilot (GAsP). Chunks with 51% genotyped variants or concordance rate < 0.92 were fused with neighbouring chunks and re-imputed. Finally, imputed variants were filtered out for r^2^ < 0.60, MAF < 0.02, mind < 0.1, HWE < 10^-5^ using Plink (v1.90). After quality control procedures had been performed, 2,415 children with 4,261,603 SNPs were included in the final analysis. Association analyses were performed using PLINK, fitting an additive model to the linear regression model with adjustment for sex, age, and the first two principal components [7].

#### **Biological annotations**

Genome-wide significant variants and the closest gene(s) were annotated using external reference data through FUMA v1.3.6a [8] (unless otherwise specified) and evaluated for functional or regulatory impact. Specifically, we considered the following annotations of SNPs reaching genome-wide significance (*p* < 5 x 10^-8^) (Supplementary Table 10):

- **Gene context:**
  - **Distance:** The distance of the variant to the nearest gene in kb. Variants within the gene body or 1 kb up- or downstream of the transcription start site (TSS) or transcription end site (TES) have a value of zero.
  - **Function:** Whether a variant is intergenic or the functional region in which the variant is located within a gene or RNA locus (e.g., 5’ UTR).
- **Combined Annotation Dependent Depletion (CADD) score:** A score of the deleteriousness of variants computed from 63 integrated annotations [9]. The higher the score, the more deleterious a variant is: 12.37 is the threshold indicated by the study of potentially actionable exonic pathogenic single-nucleotide variants in European- and African ancestry patients [10].
- **RegulomeDB category (RDB):** A variant classification system in which variants are grouped according to evidence of having a functional consequence from Category 6 (minimal evidence) to Category 1a (likely to affect binding and linked to expression of a gene target) [11].
- **Chromatin state:** The minimum and the most common 15-core chromatin state across 127 tissue/cell types predicted by ChromHMM [12] from 15 (quiescent/low) to 1 (active TSS).
- **GWAS Catalog:** SNP-trait associations reported in the NHGRI-EBI Catalog of human GWAS [13], including for each variant: the trait(s), the effect allele(s), the PubMed ID(s), the study title(s) and the study sample size(s) (Supplementary Table 2).

And the following annotations of genes within which genome-wide significant SNPs are located, or where SNPs are intergenic, the closest gene(s) up or downstream (Supplementary Table 12):

- **Probability of Loss-of-function Intolerance (pLI) score:** A score of intolerance to functional mutation from the ExAC database [14] ranging from zero to one. The closer the score is to one, the more intolerant the gene is to loss-of-function mutations. The threshold suggested by Lek et al., [14] for likely disease-causing variants is ≥ 0.9.
- **Non-coding Residual Variation Intolerance Score (ncRVIS):** A score of intolerance to mutation to non-coding variants [15]. Where ncRVIS is zero, the gene has the average number of non-coding variants given its total mutational burden; when ncRVIS is greater than zero, the gene has less non-coding variation than expected; when ncRVIS is less than zero, it has more. The ncRVIS percentile reflects the rank of the gene amongst all genes. The more negative the ncRVIS, or the lower the percentile, the more intolerant to non-coding variation the gene is.
- **Residual Variation Intolerance Score (ncRVIS) percentile:** As for ncRVIS score but the percentile of the average RVIS score for the whole gene sequence.
- **Non-coding Genomic Evolutionary Rate Profiling (ncGERP) score:** Identifies constraint in non-coding regions by quantifying deficits in substitutions [15]. It is calculated by taking the average GERP++ score (see Davydov et al., [16]) across the non-coding sequence. The higher the ncGERP score, the fewer substitutions are present than what would be expected as a result of a neutral rate of evolution, and thus the more conserved are the non-coding regions of the gene. The ncGERP percentile reflects the rank of the gene amongst all genes.
- **Protein-coding Genomic Evolutionary Rate Profiling (pcGERP) percentile:** As for ncGERP score but the percentile of the average GERP score for protein-coding sequence [15].
- **Non-coding Combined Annotation Dependent Depletion (CADD) score:** As for CADD score but the average variant score across the non-coding sequence of the gene [15].
- **Non-coding Genome-Wide Annotation of Variants (ncGWAVA) score:** Predicts the combined functionality of non-coding variants across non-coding sequence [15]. It is the average GWAVA score (see Ritchie et al., [17]) of variants in the non-coding sequence, ranging from zero to one. The closer ncGWAVA to one, the more likely the variants in non-coding regions of the gene are functional.
- **Expression in the brain:** Average log2 expression in transcripts per million (TPM) per tissue type per gene from the GTEx v8 dataset [18] for 12 brain tissues: amygdala, anterior cingulate cortex, caudate basal ganglia, cerebellar hemisphere, cerebellum, cortex, frontal cortex, hippocampus, hypothalamus, nucleus accumbens basal ganglia, putamen basal ganglia, and substantia nigra (Supplementary Table 15).

#### **Partitioned heritability**

*Evolutionary analysis*

Enrichment of heritability was estimated for the following evolutionary annotations (as described in Tilot et al., [19]):

- **Human Gained Enhancers and Promoters:** These regulatory regions were identified based on differential H3K27ac and H3K4me2 patterns in the adult and foetal brain tissues of humans, macaques and mice [19, 20], and shown to be present to a significantly lesser degree in macaques and mice. Thus, these regulatory elements were gained in the last 30 million years of human evolution and may be involved in the emergence of human-specific traits [20, 21].
- **Ancient selective sweep regions:** These regions consist of unusually long regions that reached to fixation in human populations possibly due to an adaptive advantage in the last 250-650 thousand years [22].
- **Neanderthal-introgressed SNPs:** The genomic variants introduced to the human genome by the admixture of *Homo sapiens* and Neanderthal populations around 50,000 years ago [23].
- **Neanderthal Depleted Regions:** Large regions in the human genome that are depleted for Neanderthal ancestry, possibly due to the deleterious effect of the archaic sequences in hybrid individuals [24].

### **References**

1.	Henn, B.M., et al., *Cryptic Distant Relatives Are Common in Both Isolated and Cosmopolitan Genetic Samples.* PLOS ONE, 2012. **7**(4): p. e34267.

2.	Pan, J. and H. Shu, *Rapid Automatized Naming and Its Unique Contribution to Reading: Evidence from Chinese Dyslexia.* Reading development and difficulties in monolingual and bilingual Chinese children, 2014: p. 125-138.

3.	Lei, L., et al., *Developmental trajectories of reading development and impairment from ages 3 to 8 years in Chinese children.* Journal of Child Psychology and Psychiatry, 2011. **52**(2): p. 212-220.

4.	Song, S., et al., *Universal and Specific Predictors of Chinese Children With Dyslexia - Exploring the Cognitive Deficits and Subtypes.* Frontiers in psychology, 2020. **10**: p. 2904-2904.

5.	Wang, H., Chang, B. R., Li, Y. S., Lin, L. H., Liu, J., Sun, Y. L., et al. , *Modern Chinese Frequency Dictionary*. 1986, Beijing: Beijing Language Colleague Press.

6.	Anderson, C.A., et al., *Data quality control in genetic case-control association studies.* Nat Protoc, 2010. **5**(9): p. 1564-73.

7.	Chang, C.C., et al., *Second-generation PLINK: rising to the challenge of larger and richer datasets.* Gigascience, 2015. **4**: p. 7.

8.	Watanabe, K., et al., *Functional mapping and annotation of genetic associations with FUMA.* Nat Commun, 2017. **8**(1): p. 1826.

9.	Kircher, M., et al., *A general framework for estimating the relative pathogenicity of human genetic variants.* Nature genetics, 2014. **46**(3): p. 310-315.

10.	Amendola, L.M., et al., *Actionable exomic incidental findings in 6503 participants: challenges of variant classification.* Genome research, 2015. **25**(3): p. 305-315.

11.	Boyle, A.P., et al., *Annotation of functional variation in personal genomes using RegulomeDB.* Genome Res, 2012. **22**(9): p. 1790-7.

12.	Ernst, J. and M. Kellis, *ChromHMM: automating chromatin-state discovery and characterization.* Nature Methods, 2012. **9**(3): p. 215-216.

13.	Buniello, A., et al., *The NHGRI-EBI GWAS Catalog of published genome-wide association studies, targeted arrays and summary statistics 2019.* Nucleic Acids Res, 2019. **47**(D1): p. D1005-d1012.

14.	Lek, M., et al., *Analysis of protein-coding genetic variation in 60,706 humans.* Nature, 2016. **536**(7616): p. 285-291.

15.	Petrovski, S., et al., *The Intolerance of Regulatory Sequence to Genetic Variation Predicts Gene Dosage Sensitivity.* PLOS Genetics, 2015. **11**(9): p. e1005492.

16.	Davydov, E.V., et al., *Identifying a High Fraction of the Human Genome to be under Selective Constraint Using GERP++.* PLOS Computational Biology, 2010. **6**(12): p. e1001025.

17.	Ritchie, G.R.S., et al., *Functional annotation of noncoding sequence variants.* Nature methods, 2014. **11**(3): p. 294-296.

18.	Aguet, F., et al., *The GTEx Consortium atlas of genetic regulatory effects across human tissues.* bioRxiv, 2019: p. 787903.

19.	Tilot, A.K., et al., *The Evolutionary History of Common Genetic Variants Influencing Human Cortical Surface Area.* Cerebral Cortex, 2020. **31**(4): p. 1873-1887.

20.	Vermunt, M.W., et al., *Epigenomic annotation of gene regulatory alterations during evolution of the primate brain.* Nat Neurosci, 2016. **19**(3): p. 494-503.

21.	Reilly, S.K., et al., *Evolutionary genomics. Evolutionary changes in promoter and enhancer activity during human corticogenesis.* Science, 2015. **347**(6226): p. 1155-9.

22.	Peyrégne, S., et al., *Detecting ancient positive selection in humans using extended lineage sorting.* Genome Res, 2017. **27**(9): p. 1563-1572.

23.	Vernot, B. and J.M. Akey, *Resurrecting surviving Neandertal lineages from modern human genomes.* Science, 2014. **343**(6174): p. 1017-21.

24.	Vernot, B., et al., *Excavating Neandertal and Denisovan DNA from the genomes of Melanesian individuals.* Science, 2016. **352**(6282): p. 235-239.

25.	Bates, T.C., et al., *Dyslexia and DYX1C1: deficits in reading and spelling associated with a missense mutation.* Molecular Psychiatry, 2010. **15**(12): p. 1190-6.

26.	Brkanac, Z., et al., *Evaluation of candidate genes for DYX1 and DYX2 in families with dyslexia.* American Journal of Medical Genetics Part B: Neuropsychiatric Genetics, 2007. **144b**(4): p. 556-60.

27.	Carrion-Castillo, A., et al., *Association analysis of dyslexia candidate genes in a Dutch longitudinal sample.* European Journal of Human Genetics, 2017. **25**(4): p. 452-460.

28.	Chen, Y., et al., *DCDC2 gene polymorphisms are associated with developmental dyslexia in Chinese Uyghur children.* Neural Regeneration Research, 2017. **12**(2): p. 259-266.

29.	Cope, N.A., et al., *No support for association between Dyslexia Susceptibility 1 Candidate 1 and developmental dyslexia.* Molecular Psychiatry, 2005. **10**(3): p. 237-238.

30.	Couto, J.M., et al., *The KIAA0319-like (KIAA0319L) gene on chromosome 1p34 as a candidate for reading disabilities.* Journal of Neurogenetics, 2008. **22**(4): p. 295-313.

31.	Couto, J.M., et al., *Association of reading disabilities with regions marked by acetylated H3 histones in KIAA0319.* American Journal of Medical Genetics Part B: Neuropsychiatric Genetics, 2010. **153b**(2): p. 447-462.

32.	Dahdouh, F., et al., *Further evidence for DYX1C1 as a susceptibility factor for dyslexia.* Psychiatric Genetics, 2009. **19**(2): p. 59-63.

33.	Francks, C., et al., *A 77-kilobase region of chromosome 6p22.2 is associated with dyslexia in families from the United Kingdom and from the United States.* American Journal of Human Genetics, 2004. **75**(6): p. 1046-58.

34.	Harold, D., et al., *Further evidence that the KIAA0319 gene confers susceptibility to developmental dyslexia.* Molecular Psychiatry, 2006. **11**(12): p. 1085-1091.

35.	Kong, R., et al., *Genetic variant in DIP2A gene is associated with developmental dyslexia in Chinese population.* American Journal of Medical Genetics Part B: Neuropsychiatric Genetics, 2016. **171**(2): p. 203-208.

36.	Lind, P.A., et al., *Dyslexia and DCDC2: normal variation in reading and spelling is associated with DCDC2 polymorphisms in an Australian population sample.* European Journal of Human Genetics, 2010. **18**(6): p. 668-73.

37.	Mascheretti, S., et al., *KIAA0319 and ROBO1: evidence on association with reading and pleiotropic effects on language and mathematics abilities in developmental dyslexia.* Journal of Human Genetics, 2014. **59**(4): p. 189-197.

38.	Matsson, H., et al., *Polymorphisms in DCDC2 and S100B associate with developmental dyslexia.* Journal of Human Genetics, 2015. **60**(7): p. 399-401.

39.	Mozzi, A., et al., *A common genetic variant in FOXP2 is associated with language-based learning (dis)abilities: Evidence from two Italian independent samples.* American Journal of Medical Genetics Part B: Neuropsychiatric Genetics, 2017.

40.	Müller, B., et al., *ATP2C2 and DYX1C1 are putative modulators of dyslexia-related MMR.* Brain and Behavior, 2017. **7**(11): p. e00851-e00851.

41.	Müller, B., et al., *Association, characterisation and meta-analysis of SNPs linked to general reading ability in a German dyslexia case-control cohort.* Scientific Reports, 2016. **6**: p. 27901.

42.	Newbury, D.F., et al., *Investigation of dyslexia and SLI risk variants in reading- and language-impaired subjects.* Behavior Genetics, 2011. **41**(1): p. 90-104.

43.	Newbury, D.F., et al., *CMIP and ATP2C2 modulate phonological short-term memory in language impairment.* American Journal of Human Genetics, 2009. **85**(2): p. 264-272.

44.	Paracchini, S., et al., *Analysis of dyslexia candidate genes in the Raine cohort representing the general Australian population.* Genes, Brain and Behavior, 2011. **10**(2): p. 158-65.

45.	Peter, B., et al., *Replication of CNTNAP2 association with nonword repetition and support for FOXP2 association with timed reading and motor activities in a dyslexia family sample.* Journal of Neurodevelopmental Disorders, 2011. **3**(1): p. 39-49.

46.	Poelmans, G., et al., *Identification of novel dyslexia candidate genes through the analysis of a chromosomal deletion.* American Journal of Medical Genetics Part B: Neuropsychiatric Genetics, 2009. **150B**(1): p. 140-7.

47.	Scerri, T.S., et al., *Putative functional alleles of DYX1C1 are not associated with dyslexia susceptibility in a large sample of sibling pairs from the UK.* Journal of Medical Genetics, 2004. **41**(11): p. 853-7.

48.	Scerri, T.S., et al., *DCDC2, KIAA0319 and CMIP are associated with reading-related traits.* Biological Psychiatry, 2011. **70**(3): p. 237-45.

49.	Schumacher, J., et al., *Strong genetic evidence of DCDC2 as a susceptibility gene for dyslexia.* American Journal of Human Genetics, 2006. **78**(1): p. 52-62.

50.	Shao, S., et al., *The Roles of Genes in the Neuronal Migration and Neurite Outgrowth Network in Developmental Dyslexia: Single- and Multiple-Risk Genetic Variants.* Molecular Neurobiology, 2016. **53**(6): p. 3967-3975.

51.	Sun, X., et al., *ROBO1 polymorphisms, callosal connectivity, and reading skills.* Human Brain Mapping, 2017. **38**(5): p. 2616-2626.

52.	Taipale, M., et al., *A candidate gene for developmental dyslexia encodes a nuclear tetratricopeptide repeat domain protein dynamically regulated in brain.* Proceedings of the National Academy of Sciences of the United States of America, 2003. **100**(20): p. 11553-8.

53.	Tolosa, A., et al., *FOXP2 gene and language impairment in schizophrenia: association and epigenetic studies.* BMC Medical Genetics, 2010. **11**: p. 114-114.

54.	Tran, C., et al., *Association of the ROBO1 gene with reading disabilities in a family-based analysis.* Genes, Brain and Behavior, 2014. **13**(4): p. 430-438.

55.	Venkatesh, S.K., et al., *Lack of association between genetic polymorphisms in ROBO1, MRPL19/C2ORF3 and THEM2 with developmental dyslexia.* Gene, 2013. **529**(2): p. 215-9.

56.	Vernes, S.C., et al., *A functional genetic link between distinct developmental language disorders.* New England Journal of Medicine, 2008. **359**(22): p. 2337-2345.

57.	Whitehouse, A.J.O., et al., *CNTNAP2 variants affect early language development in the general population.* Genes, Brain and Behavior, 2011. **10**(4): p. 451-456.

58.	Wigg, K.G., et al., *Support for EKN1 as the susceptibility locus for dyslexia on 15q21.* Molecular Psychiatry, 2004. **9**(12): p. 1111-21.

59.	Gialluisi, A., et al., *Genome-wide association scan identifies new variants associated with a cognitive predictor of dyslexia.* Translational Psychiatry, 2019. **9**(1): p. 77.
