## Supplementary material for "Discovery of 42 Genome-Wide Significant Loci Associated with Dyslexia": GenLang Consortium Author List and Acknowledgements

Supplementary Information

**GenLang Consortium Author List**

Filippo Abbondanza 1

1 School of Medicine, Medical & Biological Sciences, University of St Andrews, St Andrews, Scotland

Andrea G Allegrini 2

2 Social, Genetic and Developmental Psychiatry Centre, Institute of Psychiatry, Psychology and Neuroscience, King’s College London, London, United Kingdom

Till F M Andlauer 3 4

3 Translational Research in Psychiatry, Max Planck Institute of Psychiatry, Munich, Germany

4 Department of Neurology, Klinikum rechts der Isar, School of Medicine, Technical University of Munich, Munich, Germany

Cathy L Barr 5 6 7

5 Division of Experimental and Translational Neuroscience, Krembil Research Institute, University Health Network, Toronto, ON, Canada

6 Program in Neuroscience and Mental Health, Hospital for Sick Children, Toronto, Ontario, Canada

7 Department of Psychiatry, University of Toronto, Toronto, Ontario, Canada

Timothy C Bates 8

8 Psychology, University of Edinburgh, Edinburgh, Scotland

Manon Bernard 9

9 Departments of Physiology and Nutritional Sciences, Hospital for Sick Children, Toronto, Ontario, Canada

Kirsten Blokland 6

Dorret I Boomsma 10 11 12

10 Netherlands Twin Register, Amsterdam, The Netherlands

11 Department of Biological Psychology, Vrije Universiteit Amsterdam, Amsterdam, Nederland, The Netherlands

12 Amsterdam Reproduction and Development (AR&D) Research Institute, Amsterdam UMC, The Netherlands

Manuel Carreiras 13 14 15

13 Basque Center on Cognition, Brain and Language (BCBL), Donostia-San Sebastian, Gipuzkoa, Spain

14 Ikerbasque Basque Foundation for Science, Bilbao, Vizcaya,Spain

15 Lengua Vasca y Comunicación. University of the Basque Country (UPV/EHU), Bilbao, Vizcaya, Spain

Fabiola Ceroni 16 17

16 Department of Pharmacy and Biotechnology, University of Bologna, Bologna, Italy

17 Faculty of Health and Life Sciences, Oxford Brookes University, Oxford, United Kingdom

Philip S Dale 18

18 Department of Speech & Hearing Sciences, University of New Mexico, Albuquerque, New Mexico, USA

Peter F de Jong 19

19 Department of Child Development and Education, University of Amsterdam, Amsterdam, Nederland, The Netherlands

Eveline L de Zeeuw 11

John C DeFries 20 21

20 Institute for Behavioral Genetics, University of Colorado, Boulder Colorado United States

21 Department of Psychology and Neuroscience, University of Colorado, Boulder, Colorado, United States

Else Eising 22

22 Language and Genetics department, Max Planck Institute for Psycholinguistics, Nijmegen, The Netherlands

Yu Feng 23

23 Genetics and Development Division, Krembil Research Institute, University Health Network, Toronto, Ontario, Canada

Simon E Fisher 22 24

24 Donders Institute for Brain, Cognition and Behaviour, Nijmegen, The Netherlands

Marie-Christine J Franken 25

25 Department of Otorhinolaryngology, Erasmus University Medical Centre, Rotterdam, The Netherlands

Clyde Franks 22 24

Alessandro Gialluisi 3 26

26 Department of Epidemiology and Prevention, IRCCS Istituto Neurologico Mediterraneo Neuromed, Pozzilli, Italy

Scott D Gordon 27

27 Genetic Epidemiology Laboratory, QIMR Berghofer Medical Research Institute, Brisbane, Queensland, Australia

Jeffrey R Gruen 28

28 Departments of Pediatrics and Genetics, Yale Medical School, New Haven, Connecticut, USA

Sharon L Guger 29

29 Department of Psychology, Hospital for Sick Children, Toronto, Ontario, Canada

Marianna E Hayiou-Thomas 30

30 Department of Psychology, University of York, York, United Kingdom

Juan Hernández-Cabrera 31

31 Departamento de Psicología Clínica Psicobiología y Metodología, La Laguna, Santa Cruz de Tenerife, Spain

Jouke- Jan Hottenga 11

Charles Hulme 32

32 Department of Education, University of Oxford, Oxford, Oxfordshire, United Kingdom

Philip R Jansen 33 34

33 Department of Child and Adolescent Psychiatry/Psychology, Erasmus University Medical Center, Rotterdam, The Netherlands

34 Department of Complex Trait Genetics, Center for Neurogenomics and Cognitive Research, Amsterdam Neuroscience, VU University, Amsterdam, The Netherlands

Elizabeth N Kerr 28 35 36

35 Department of Neurology, Hospital for Sick Children, Toronto, Ontario, Canada

36 Department of Paediatrics, The University of Toronto, Toronto, Ontario, Canada

Tanner Koomar 37

37 Department of Psychiatry, University of Iowa, Iowa City, Iowa, USA

Gabriel T Leonard 38

38 Cognitive Neuroscience Neurology and Neurosurgery, Montreal, Quebec, Canada

Zhijie Liao 39

39 Department of Psychology, University of Toronto, Toronto, Ontario, Canada

Maureen W Lovett 6 36

Michelle Luciano 8

Nicholas G Martin 27

Angela Martinelli 1

Jacob J Michaelson 37

Nazanin Mirza-Schreiber 40

40 Institute of Neurogenomics, Helmholtz Zentrum München, Munich, Germany

Kristina Moll 41

41 Department of Child and Adolescent Psychiatry, Psychosomatics, and Psychotherapy, LMU University Hospital Munich, Munich, Germany

Anthony P Monaco 42

42 Office of the President, Tufts University, Medford, MA 02155, USA

Angela T Morgan 43 44 45

43 Speech and Language, Murdoch Children's Research Institute, Melbourne, Victoria, Australia

44 Department of Audiology and Speech Pathology, University of Melbourne, Melbourne, Victoria, Australia

45 Speech Pathology Department, Royal Children's Hospital, Melbourne, Victoria, Australia

Bertram Müller-Myhsok 3 46

46 Department of Health Science, University of Liverpool, Liverpool, United Kingdom

Dianne F Newbury 17

Markus M Nöthen 47

47 Institute of Human Genetics, University Hospital of Bonn, Bonn, Germany

Richard K Olson 20

Silvia Paracchini	1

Tomas Paus 7 48 49

48 Departments of Psychiatry and Neuroscience and Centre Hospitalier Universitaire Sainte Justine, University of Montreal, Montreal, Quebec, Canada

49 Department of Psychology, University of Toronto, Toronto, Ontario, Canada

Zdenka Pausova 9, 50

50 Hospital for Sick Children, Toronto, Ontario, Canada

Craig E Pennell 51 52 53

51 School of Medicine and Public Health, University of Newcastle, Newcastle New South Wales Australia

52 Mothers and Babies Research Centre, Hunter Medical Research Institute, Newcastle, New South Wales, Australia

53 Maternity and Gynaecology, John Hunter Hospital, Newcastle, New South Wales, Australia

Bruce F Pennington 54

54 Department of Psychology at the University of Denver, Denver, Colorado, USA

Robert J Plomin 2

Kaitlyn M Price 6 23 55

55 Department of Physiology, University of Toronto, Toronto, Ontario, Canada

Sheena Reilly 42 56

56 Menzies Health Institute Queensland, Griffith University, Gold Coast, Queensland, Australia

Louis Richer 57

57 Department of Health Sciences, Université du Québec à Chicoutimi, Chicoutimi, Québec, Canada

Kaili Rimfeld 2

Gerd Schulte-Körne 41

Chin Yang Shapland 58 59

58 MRC Integrative Epidemiology Unit, University of Bristol, Bristol, UK

59 Population Health Sciences, University of Bristol, Bristol, UK

Nuala H Simpson 60

60 Department of Experimental Psychology, University of Oxford, Oxford, UK

Shelley D Smith 61

61 Department of Neurological Sciences, College of Medicine, University of Nebraska Medical Center, Omaha, Nebraska, USA

Margaret J Snowling 60 62

62 St John’s College, University of Oxford, Oxford, UK

Beate St Pourcain 22 24 58

John F Stein 63

63 Department of Physiology, Anatomy and Genetics, Oxford University, **Oxford OX1 3PT, UK**

Lisa J Strug 64 65

64 Departments of Statistical Sciences and Computer Science and Division of Biostatistics, The University of Toronto, Toronto, Ontario, Canada

65 Program in Genetics and Genome Biology and The Centre for Applied Genomics, The Hospital For Sick Children, Toronto, Ontario, Canada

Joel B Talcott 66

66 Developmental Cognitive Neuroscience, Aston Brain Centre, Birmingham, B4 7ET, UK

Henning Tiemeier 32 67

67 Harvard, T.H. Chan School of Public Health, Boston, MA, USA

James B Tomblin 68

68 Communication Sciences and Disorders, University of Iowa, Iowa City, Iowa, United States

Dongnhu T Truong 28

Elsje van Bergen 11 69

69 Research Institute LEARN!, Vrije Universiteit Amsterdam, Amsterdam, The Netherlands

Marc MP van de Schroeff 70 71

70 Department of Otolaryngology, Head and Neck Surgery, Erasmus MC, Rotterdam, Netherlands

71 Generation R Study Group, Erasmus MC, Rotterdam, Netherlands

Marjolein Van Donkelaar 22

Ellen Verhoef 22

Carol A Wang 51 52

Kate E Watkins 60

Andrew JO Whitehouse 72

72 Telethon Kids Institute, The University of Western Australia, Perth, Western Australia, Australia

Karen G Wigg 23

Erik G Willcutt 20

Margaret Wilkinson 6

Margaret J Wright 73

73 Queensland Brain Institute, University of Queensland, Brisbane, Australia

Gu Zhu 27

**GenLang Quantitative Trait Consortium Acknowledgements**

Adolescent Brain Cognitive Development (ABCD) Study

Data used in the preparation of this article were obtained from the Adolescent Brain Cognitive Development (ABCD) Study (<https://abcdstudy.org>), held in the NIMH Data Archive (NDA). This is a multisite, longitudinal study designed to recruit more than 10,000 children age 9-10 and follow them over 10 years into early adulthood. The ABCD Study® is supported by the National Institutes of Health and additional federal partners under award numbers U01DA041048, U01DA050989, U01DA051016, U01DA041022, U01DA051018, U01DA051037, U01DA050987, U01DA041174, U01DA041106, U01DA041117, U01DA041028, U01DA041134, U01DA050988, U01DA051039, U01DA041156, U01DA041025, U01DA041120, U01DA051038, U01DA041148, U01DA041093, U01DA041089, U24DA041123, U24DA041147. A full list of supporters is available at <https://abcdstudy.org/federal-partners.html>. A listing of participating sites and a complete listing of the study investigators can be found at <https://abcdstudy.org/consortium_members/>. ABCD consortium investigators designed and implemented the study and/or provided data but did not necessarily participate in the analysis or writing of this report. This manuscript reflects the views of the authors and may not reflect the opinions or views of the NIH or ABCD consortium investigators.

Aston

Data collection for the Aston Cohort has been supported by funding from the European Union Horizon 2020 Programme (641652), The Waterloo Foundation (797/17290), and with the invaluable support of the participants and their families.

Avon Longitudinal Study of Parents and their Children (ALSPAC)

We are extremely grateful to all the families who took part in this study, the midwives for their help in recruiting them, and the whole ALSPAC team, which includes interviewers, computer and laboratory technicians, clerical workers, research scientists, volunteers, managers, receptionists and nurses. GWAS data was generated by Sample Logistics and Genotyping Facilities at Wellcome Sanger Institute and LabCorp (Laboratory Corporation of America) using support from 23andMe. The UK Medical Research Council and Wellcome (Grant ref: 217065/Z/19/Z) and the University of Bristol provide core support for ALSPAC. This publication is the work of the authors and they will serve as guarantors for the contents of this paper. A comprehensive list of grants funding is available on the ALSPAC website (<http://www.bristol.ac.uk/alspac/external/documents/grant-acknowledgements.pdf>)

The Basque Center on Cognition, Brain and Language (BCBL)

We thank the study participants and all staff who supported the project.

Brisbane Adolescent Twins Study (BATS)

We thank the twins and their families for participating in the study; the staff working on the project, including study coordinators, research nurses, DNA sample processing and preparation technicians; information technologists; and colleagues who assisted in the quality control and preparation of the imputed GWAS data. The research was supported by the Australian Research Council (A7960034, A79906588, A79801419, DP0212016 and DP0343921), with genotyping funded by the National Health and Medical Research Council (Medical Bioinformatics Genomics Proteomics Program, 389891).

Colorado Learning Disabilities Research Center (CLDRC)

We thank the participants and their families and acknowledge financial support from a NICHD grant P50 HD 27802.

The Early Language in Victoria Study (ELVS)

Thank the families for participating and the study staff involved in project management, recruitment and data collection. Funding by the National Health and Medical Research Council grant number 436958 is gratefully appreciated.

Familial Influences on Literacy Abilities (FIOLA)

The FIOLA Project was developed by E. van Bergen, T. van Zuijen, and P.F. de Jong, and is supported by the University of Amsterdam, the MPI Nijmegen, and by fellowships awarded to Elsje van Bergen (NWO’s Rubicon 446-12-005 and VENI 451-15-017). We are grateful to all participating families and the NEMO Science Museum.

Generation R

The Generation R Study is conducted by the Erasmus Medical Center in close collaboration with School of Law and Faculty of Social Sciences of the Erasmus University Rotterdam, the Municipal Health Service Rotterdam area, the Rotterdam Homecare Foundation, and the Stichting Trombosedienst & Artsenlaboratorium Rijnmond (STAR[1]MDC), Rotterdam. We gratefully acknowledge the contribution of children and parents, general practitioners, hospitals, midwives and pharmacies in Rotterdam for their participation in the Generation R Study.

The Genes, Reading and Dyslexia (GRaD) Study

We thank the study participants and all staff who supported the project. The Genes, Reading and Dyslexia (GRaD) Study was funded by the generous support of the Manton Foundation , the National Institutes of Health (Grant ref: P50-HD027802; K99-HD094902), and the Lambert Family.

Iowa

We are grateful for the contributions of the participants of the Iowa language cohort and their families. JJM, JBT, and TK were supported by NIH grant DC014489.

Neurodys

A.G. and T.F.M.A. were supported by the Munich Cluster for Systems Neurology (SyNergy). S.P. is a Royal Society University Research fellow. B.M.M., C.F., B.S.P. and S.E.F. are supported by the Max Planck Society. A.W., B.M. and H.K. were funded by the Fraunhofer Society and the Max Planck Society within the ‘Pakt für Forschung und Innovation’.  F.R. is supported by Agence Nationale de la Recherche (ANR-06-NEURO-019-01, ANR-17-EURE-0017 IEC, ANR-10-IDEX-0001-02 PSL, ANR-11-BSV4-014-01), European Commission (LSHM-CT-2005-018696).

We would also like to acknowledge our project partners Catherine Billard, Caroline Bogliotti, Vanessa Bongiovanni, Laure Bricout, Camille Chabernaud, Isabelle Comte-Gervais, Florence Delteil-Pinton, Florence George, Christophe-Loïc Gérard, Marie Lageat, Marie-France Leheuzey, Marie-Thérèse Lenormand, Marion Liébert, Emilie Longeras, Emilie Racaud, Isabelle Soares-Boucaud, Sylviane Valdois, Nadège Villiermet, and Johannes Ziegler.

Pediatric Imaging, Neurocognition and Genetics Study (PING)

Data collection and sharing for this project was funded by the Pediatric Imaging, Neurocognition and Genetics Study (PING) (National Institutes of Health Grant RC2DA029475). PING is funded by the National Institute on Drug Abuse and the Eunice Kennedy Shriver National Institute of Child Health & Human Development. PING data are disseminated by the PING Coordinating Center at the Center for Human Development, University of California, San Diego.

The Philadelphia Neurodevelopmental Cohort (PNC)

Support for the collection of the data sets was provided by grant RC2MH089983 awarded to R Gur and RC2MH089924 awarded to H Hakonarson. Subjects were recruited through the Center for Applied Genomics at The Children’s Hospital in Philadelphia.

Raine Study

The authors are grateful to the Raine Study participants and their families, and to the Raine Study team for cohort coordination and data collection. The Raine Study was supported by the National Health and Medical Research Council of Australia [grant numbers 572613, 403981, 1059711], the Canadian Institutes of Health Research [grant number MOP-82893], and WA Health, Government of Western Australia (WADOH) [Future Health WA G06302]. Funding was also generously provided by Safe Work Australia. The authors gratefully acknowledge the NHMRC for their long-term funding to the study over the last 30 years and also the following institutes for providing funding for Core Management of the Raine Study: The University of Western Australia (UWA), Curtin University, Women and Infants Research Foundation, Telethon Kids Institute, Edith Cowan University, Murdoch University, The University of Notre Dame Australia and The Raine Medical Research Foundation. This work was supported by resources provided by the Pawsey Supercomputing Centre with funding from the Australian Government and Government of Western Australia.

Saguenay Youth Study (SYS)

We thank the study participants and their families, and all staff who have supported the study. The Canadian Institutes of Health Research (ZP, TP), Heart and Stroke Foundation of Quebec (ZP), and the Canadian Foundation for Innovation (ZP) support the Saguenay Youth Study.

SLI Consortium

SLI Consortium members are as follows: Wellcome Trust Centre for Human Genetics, Oxford: D. F. Newbury, N. H. Simpson, F. Ceroni, A. P. Monaco; Max Planck Institute for Psycholinguistics, Nijmegen: S. E. Fisher, C. Francks; Newcomen Centre, Evelina Children’s Hospital, St Thomas’ Hospital, London: G. Baird, V. Slonims; Child and Adolescent Psychiatry Department and Medical Research Council Centre for Social, Developmental, and Genetic Psychiatry, Institute of Psychiatry, London: P. F. Bolton; Medical Research Council Centre for Social, Developmental, and Genetic Psychiatry Institute of Psychiatry, London: E. Simonoff; Salvesen Mindroom Centre, Child Life & Health, School of Clinical Sciences, University of Edinburgh: A. O’Hare; Cell Biology & Genetics Research Centre, St. George’s University of London: J. Nasir; Queen’s Medical Research Institute, University of Edinburgh: J. Seckl; Department of Speech and Language Therapy, Royal Hospital for Sick Children, Edinburgh: H. Cowie; Speech and Hearing Sciences, Queen Margaret University: A. Clark, J. Watson; Department of Educational and Professional Studies, University of Strathclyde: W. Cohen; Department of Child Health, the University of Aberdeen: A. Everitt, E. R. Hennessy, D. Shaw, P. J. Helms; Audiology and Deafness, School of Psychological Sciences, University of Manchester: Z. Simkin, G. Conti-Ramsden; Department of Experimental Psychology, University of Oxford: D. V. M. Bishop; Biostatistics Department, Institute of Psychiatry, London: A. Pickles.

The Twins Early Development Study (TEDS)

We gratefully acknowledge the on-going contribution of the participants in the Twins Early Development Study (TEDS) and their families. TEDS is supported by a programme Grant to R.P. from the UK Medical Research Council (Grant Nos. MR/V012878/1 and previously MR/M021475/1), with additional support from the US National Institutes of Health (Grant No. AG046938). The research leading to these results has also received funding from the European Research Council under the European Union's Seventh Framework Programme (FP7/2007-2013)/ grant agreement n 602768. K.R. is supported by a Sir Henry Wellcome Postdoctoral Fellowship.

Toronto

We thank the psychologists, coordinators, psychometrists and volunteers that assisted with this project over the years and the participants in this study. Support for this project was provided by grants from the Canadian Institutes of Health Research (MOP-133440). K.P. was supported by the Hospital for Sick Children Research Training Program (Restracomp).

UK Dyslexia (UKdys)

Thank the families for participating. Cohort recruitment and data collection was supported by Wellcome Trust (076566/Z/05/Z); (075491/Z/04) and The Waterloo Foundation [grants to JBT and SP; 797–1720]. Genotype data were generated and analyses funded by the EU [Neurodys, 018696] and the Royal Society [UF100463 grant to SP].

York

The York cohort was funded by Wellcome Trust Programme Grant 082036/B/07/Z. We would like to thank the team who collected the data and the families who participated.
